# Biobank-scale methods and projections for sparse polygenic prediction from machine learning

**DOI:** 10.1101/2023.03.06.23286870

**Authors:** Timothy G. Raben, Louis Lello, Erik Widen, Stephen D.H. Hsu

## Abstract

In this paper we characterize the performance of linear models trained via widely-used *sparse* machine learning algorithms. We build polygenic scores and examine performance as a function of training set size, genetic ancestral background, and training method. We show that predictor performance is most strongly dependent on size of training data, with smaller gains from algorithmic improvements. We find that LASSO generally performs as well as the best methods, judged by a variety of metrics. We also investigate performance characteristics of predictors trained on one genetic ancestry group when applied to another. Using LASSO, we develop a novel method for projecting AUC and Correlation as a function of data size (i.e., for new biobanks) and characterize the asymptotic limit of performance. Additionally, for LASSO (compressed sensing) we show that performance metrics and predictor sparsity are in agreement with theoretical predictions from the Donoho-Tanner phase transition. Specifically, a predictor trained in the Taiwan Precision Medicine Initiative for asthma can achieve an AUC of 0.63_(0.02)_ and for height a correlation of 0.648_(0.009)_ for a Taiwanese population. This is above the measured values of 0.61_(0.01)_ and 0.631_(0.008)_, respectively, for UK Biobank trained predictors applied to a European population.

## 1 Introduction

Given the complexity of the human genome, large datasets are required to detect associations between specific genetic variations and their effect on phenotypes. These large datasets provide the statistical power necessary to overcome false signals (fluctuations) resulting from examination of millions of genetic variants at a time. With the advent of very large biobanks [1–3], which collect millions of individual genotypes and associated phenotypes, it has become possible to probe the genetic architectures of important disease risks and other complex traits.

The analysis of large genotype and phenotype datasets has led to the development of polygenic scores (PGS). A PGS is simply a score built from numerically weighting the state of a persons genome. In most work to date, and in this paper, we are interested in linear PGS built from single nucleotide polymorphisms (SNPs), i.e., 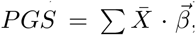, for genotype matrix X̄ and SNP weights 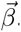. SNP weights are typically obtained through a machine learning algorithm on genotype/phenotype pairs and can be as simple as single marker regression (e.g., Genome-Wide-Association-Studies or GWAS).

The vast majority of available GWAS and biobank data is from individuals of European ancestry. For example, the UK Biobank [3] (UKB) is *>* 90% self-reported white. As a consequence, current PGS perform better for descendants of Europeans. There are a number of new biobank-scale efforts, focusing on non-European populations, which will ameliorate this situation; e.g., the Taiwan Precision Medicine Initiative [4] (TPMI). Other novel projects have focused on gathering samples with ancestral equity in mind, e.g., All of Us [5] (AoU). However, until more diverse data become available, it is necessary to adapt the European results for other ancestral populations in order for PGS to have utility for the largest number of individuals possible – i.e., in applications such as disease risk estimates, potential clinical interventions, etc.

As genotype databases become larger and as sequencing technology incorporates more SNPs (i.e., increasing the number of SNPs through imputation, larger arrays, and whole genome sequencing or WGS), novel difficulties arise in PGS construction. First, larger samples and features require greater computational power (we comment on the computational requirements for the results in this paper in the Supplementary Information). Second, and as mentioned above, the application of PGS has been largely restricted to those of European ancestry [6–17]. In order to “transport” PGS to other ancestry groups, many techniques have been proposed, using features that are most important in different groups and adjusting their specific weights [14, 18–22] (e.g., by using minor allele frequency differences or functional information). The complexity of this analysis clearly scales with the number of relevant features. Third, future benefits of PGS [7, 10, 16, 17,23–43] rely on genotyping future participants. If this genotyping can be restricted to a small number of SNPs (e.g. as opposed to more costly WGS) it can be more cost effective to implement. Fourth, most PGS development approaches and methods use linear models. Further challenges include non-linear SNP effects (e.g., which are responsible for the difference between narrow and broad sense heritability [44–50]), the relationship between tagged vs causal SNPs, and incorporating genome-environment interactions. Addressing these challenges again scales with the number of relevant features.

In this paper we focus on the performance, detailed application, and future power of *sparse* algorithms, i.e. algorithms that perform feature selection, for the 11 traits listed in **Table 1**. Performing feature selection can help ameliorate some of the issues raised above. Sparse algorithms have previously been shown to be comparable to non-sparse methods in terms of standard metrics (e.g., area under receiver operator curve, correlation, *r*^2^, etc.) [10, 51–53]. We focus on 11 phenotypes that have been previously been shown to cover a wide range of sparsities (see **Table 1**): asthma, atrial fibrillation, breast cancer, coronary artery disease (CAD), hypertension, type 1 and type 2 diabetes (T1D, T2D), body mass index (BMI), direct bilirubin, height, and lipoprotein A.

**Table 1:** The 11 phenotypes studied in this work and their relative sparsity. As described in section 3.3 predictor sparsity can be defined in a variety of ways. Here we color-code the various traits according to the order of magnitude of SNP sparsity. Numbers in parentheses are the approximate number of SNPs used in a LASSO trained predictor using the maximum amount of data from the UKB. This definition of sparsity is consistent with the number of SNPs with non-zero weights found in previous publications[10, 11].

| | high sparsity<br>$\mathcal{O}(10^2)$SNPs | moderate sparsity<br>$\mathcal{O}(10^3)$SNPs | low sparsity<br>$\mathcal{O}(10^4)$SNPs |
| --- | --- | --- | --- |
| Case/Control | breast cancer ($\sim 500$)<br>type 1 diabetes ($\sim 50$) | asthma ($\sim 3.5k$)<br>type 2 diabetes ($\sim 4k$)<br>atrial fibrillation ($\sim 3k$) | hypertension ($\sim 10k$)<br>CAD ($\sim 7.5k$) |
| Continuous | lipoprotein A ($\sim 170$) | direct bilirubin ($\sim 3k$) | height ($\sim 22.5k$)<br>BMI ($\sim 24.5k$) |

As mentioned, there are many efforts to modify PGS trained in primarily European ancestries to improve performance in non-European ancestries. As we discuss, these improvements, while important, are unlikely to close the gap completely. It seems necessary to create large data cohorts in each ancestry group. This is difficult and expensive. Hence it is valuable to understand in advance what the resulting benefits will be for polygenic prediction. In this work we present a novel method for projecting and predicting the results from sparse algorithms in a novel biobank.

The main results of this paper can be summarized as follows

1. Widely-used sparse methods perform comparably, with a simple LASSO-based approach regularly achieving the best results.
2. Increased biobank/database size and access to new datasets will lead to large gains, especially for the performance of PGS in diverse ancestries.
3. We develop a novel method which predicts correlation and AUC for continuous and case/control phenotypes (respectively), with uncertainty bands, for biobank-sized datasets.
4. We explore details of “phase change” behavior of compressed sensing/LASSO with increasing data size, and the corresponding the SNP content of resulting predictors.

## 2 Results

Here we present the main results of this project. Additional details concerning specific methods are found in section 3 and in the Supplementary Information. The main results are obtained using UKB genotype-phenotype data. Training with PRScs at times uses linkage disequilibrium (LD) information from the 1,000 Genomes Project[1] (1kg). Projections are given for de novo training in TPMI and AoU. We refer to ancestry groupings European (EUR), South Asian (SAS), East Asian (EAS), African (AFR), and American (AMR). These labeling conventions come from [54].

### 2.1 Comparison of Sparse predictors

We compare the performance of several sparse methods: LASSO, Elastic Net, L1-penalized Logistic regression (for case-control conditions), and PRScs with LD matrix information from either UKB or 1KG. It is important to note that the results presented here for AUC and correlation are for *purely genetic* PGS. In brief, phenotypes are first regressed on covariates (such as age, sex, and the first 20 genetic principal components) - this allows them to explain as much of the variance as possible so that we can conservatively estimate purely genetic effects. Only then are SNP predictors are trained (using genotype-phenotype data or summary statistics in the case of PRScs) on residual phenotypes. Further details about the training and evaluation of PGS can be found in section 3.

In **Figure 1** we see the comparison results for asthma and height. Similar plots for the other phenotypes can be found in appendix D. Ancestry groups SAS, AFR, EAS, and EUR result from UKB definitions of self-reported ancestry (although training, as described in section 3 involves a principal component, or PC, adjustment). AMR refers to an American–like group constructed via principal component clustering detailed in appendix A.1 and similar to that found in [55]. Sib refers to a set of white siblings (i.e. every member of the set has at least one sibling also in the set) where the ancestry is self-reported, but the sibling status is determined by a genetic analysis as detailed in appendix A.2. This sib-set attempts to partially control for environmental effects as described in [56, 57]. It also allows for performing sibling selection experiments as described in appendix A.2. All results reflect training on a EUR population and then applied to a set of siblings or a different ancestry group, not used in training. The bands for TMPI/AoU are based on projections for future biobanks - this is described in detail in section 2.2.

**Figure 1:**
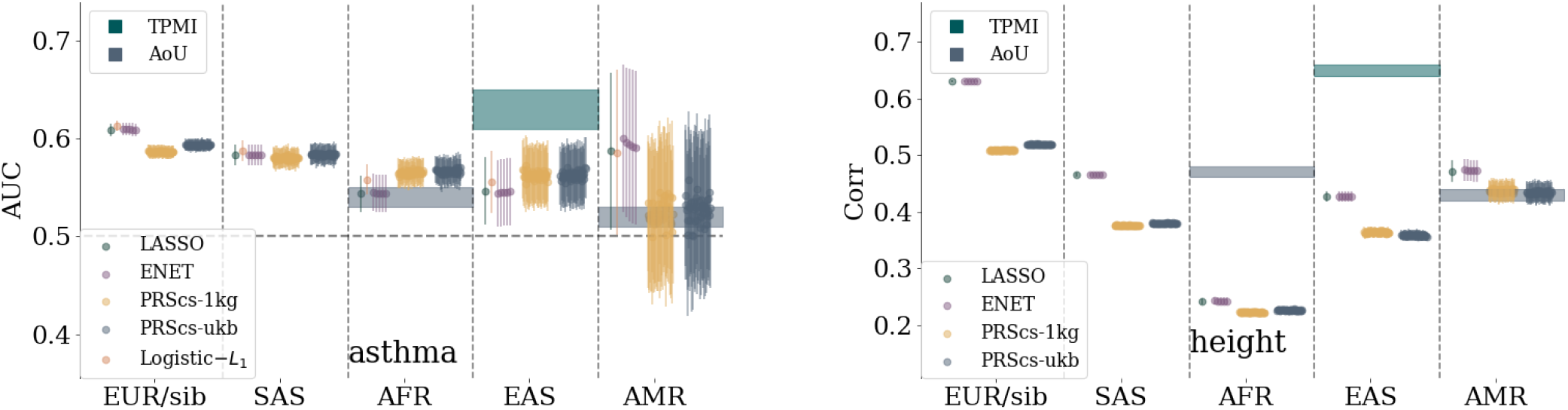
Comparison of sparse methods for asthma and height predictors with a comparison to prediction bands for more diverse biobanks. On the left, asthma predictors trained on a UKB white population. Predictors are built with LASSO, *L*_1_-penalized Logistic regression, Elastic Nets, and PRScs with UKB and 1,000 Genomes LD matrices. The specific parameters for the Elastic nets and PRScs are described in section 3. Similar results for the other phenotypes can be found in appendix D.

Uncertainty (error bars) depend on cross-validation (i.e. multiple training sets), finite size effects from computing AUC/correlation, and from sample sizes. Details about uncertainty calculations are given in the Supplementary Information. For case-control conditions this can lead to error bars that are the same size as the central values. However, for continuous phenotypes with much larger sample sizes this is not the case. For example, compare the AMR group on both plots in **Figure 1**. On the left, the AUC error bar overlaps 0.5 (i.e. consistent with no signal), while on the right, the correlation error bar is relatively small.

Comparing performance across ancestry groups, we see a well known fall-off behavior which is observed when predictors trained in one population are applied to another (e.g., see [52, 55]). The amount of fall-off is phenotype specific and ranges from complete fall-off, e.g. diabetes fall-off from sib to AFR, to negligible fall-off, e.g. breast cancer AUC from sibs to EAS. The relative order of fall-off is also phenotype specific. For most traits, the (EUR-ancestry) sibling set shows the least reduction – then either SAS, EAS, and AMR, – and finally AFR. However, there are clear exceptions like the direct bilirubin correlation which goes from largest metric to smallest: Sib, SAS, AFR, EAS, AMR. There have been recent arguments that PGS fall-off is roughly linear as a function of local genetic distance [52]. We should note that this claim is not necessarily in conflict with the results we present here. First, there are many exceptions to this general linear behavior, e.g., Figure 5 in [52]. Second, the claimed linear fall-off is a function of Euclidean distance in PC space of training population. For genetically distant groups, the axes of variation will be different: any measure of PC distance is therefore a *local* measure. In other words, only ancestry groups that are near-enough to the original population where PCs where computed can be considered well-ordered in terms of genetic difference. These effects can also be exacerbated by the fact that these are all *sparse* predictors and, after projecting onto PC space, the order of Euclidean genetic-distance may change.

The performance of PGS can always be confounded due to environmental factors, interactions between genes and the environment, and non-linear genetic effects (e.g., epistasis). To attempt to guard against some of these effects we can perform sibling tests similar to those described in [56, 57]. Genetic siblings can be assumed to have, on average, a more similar environmental background than unrelated individuals. However, there can be a competing effect from the enhancement of signals from effects like genetic nurture [58]. For case control conditions we can create *affected sibling pairs* (ASPs) where one person is a case and one is a control. Then we can ask what fraction of the time does the higher PGS correspond to the case vs control. We can also condition this question on the PGS difference being larger than some cut off (e.g., 1.5, 2, or 2.5 standard deviations). For continuous phenotypes we can simply compare the fraction of the time where the person with the higher PGS also has the higher phenotype value. We can again condition this question for siblings whose phenotypes are separated by a cut off (e.g., 0.5, 1, or 1.5 standard deviations). These selection rates for asthma and BMI are shown in **Figure 2** as an example; the results for all traits considered in this work can be found in the Supplementary Information. Again, we see that while several methods are extremely competitive, LASSO is regularly among the best performing methods. For larger and larger cut-offs, the selection rate improves, but the associated uncertainty also increases largely because of decreasing sample sizes. Interestingly, while PRScs performed similarly to other methods in terms of AUC and correlation, it routinely under performs methods training directly on genetic data in sibling selection tests.

**Figure 2:**
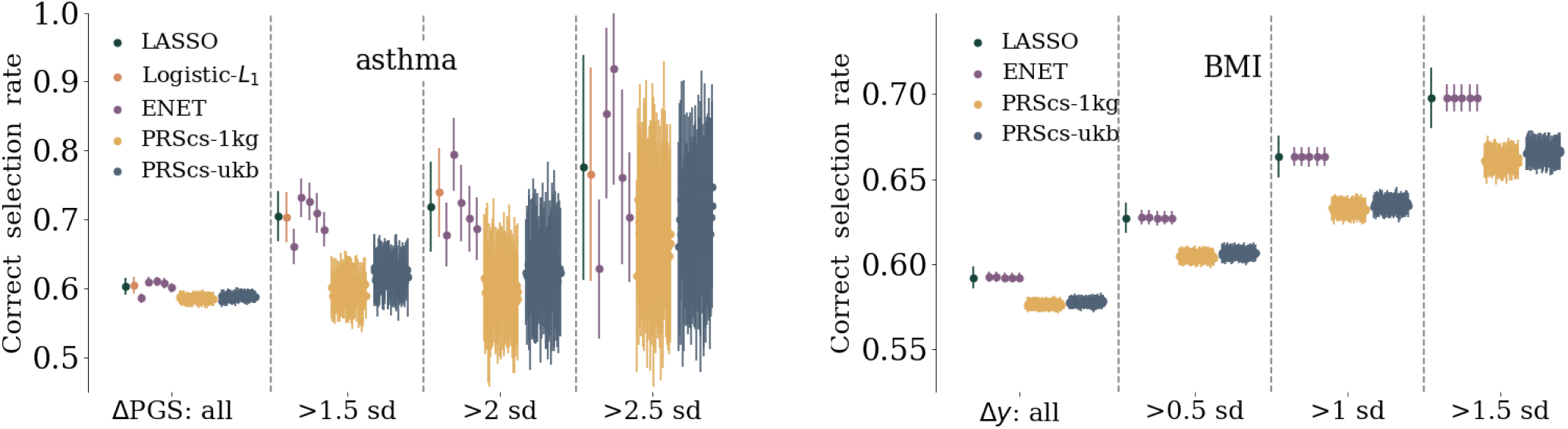
Left: affected sibling pair (ASP) selection rate for asthma. Pairs of siblings, where one person is a case and the other a control, are used and the rate corresponds to the number of times the case sibling has the higher PGS. The rate of correct selection, and uncertainty, increases if the siblings are also separated by at least 1.5, 2, or 2.5 standard deviations in PGS. Right: rank order selection rate for BMI respectively. The rate corresponds to frequency of the sibling with the larger BMI also having the larger PGS. Again the selection rate, and uncertainty (due to reduced statistics), increase if the sibling BMI is required to differ by at least 0.5, 1, or 1.5 standard deviations. Similar results for the other phenotypes are found in the Supplementary Information.

PGS for case-control conditions can be converted to more clinically interpretable metrics. In **Figure 3** we see an example of an *inclusive* odds ratio (OR) for asthma. It is inclusive in the sense that the OR corresponds to the ratio of the cases to controls (normalized by the ratio of the total cases to controls) *at a specific PGS value or above*. As before uncertainties are conservative and include contributions from multiple cross-validation folds and finite size effects. Similar plots for the other case control conditions can be found in the Supplementary Information. Because LASSO routinely performed among the best predictors in terms of AUC and correlation we only present OR plots for this method. Analogous plots for the other methods can be generated similarly, although for all 7 case-control traits it leads to 357 plots which can be difficult to interpret. An initial interpretation of these results is that at extreme values of PGS there are large increases in OR: for asthma, within ancestry testing (i.e. the sibling group) leads to 2 *<* OR *<* 2.5 at large PGS. When testing on other ancestries, an optimistic interpretation is that asthma OR ranges from 1.25-3.5 at large PGS. While this is encouraging, we also urge caution. The extremes of the PGS distributions are the regions where model assumptions are most likely to break down, e.g., the linearity of SNP effects. Additionally, the sample sizes in these regions are smallest which leads to large uncertainties and difficulty interpreting the results. In addition, odds ratios are difficult to model in the presence of non-Gaussian distributions. The inclusive odds ratio can be written as a ratio of cumulative distribution functions of cases and controls. Similarly the PGS percentile can be written as an integral over the sum of the probability distribution functions for cases and controls.

**Figure 3:**
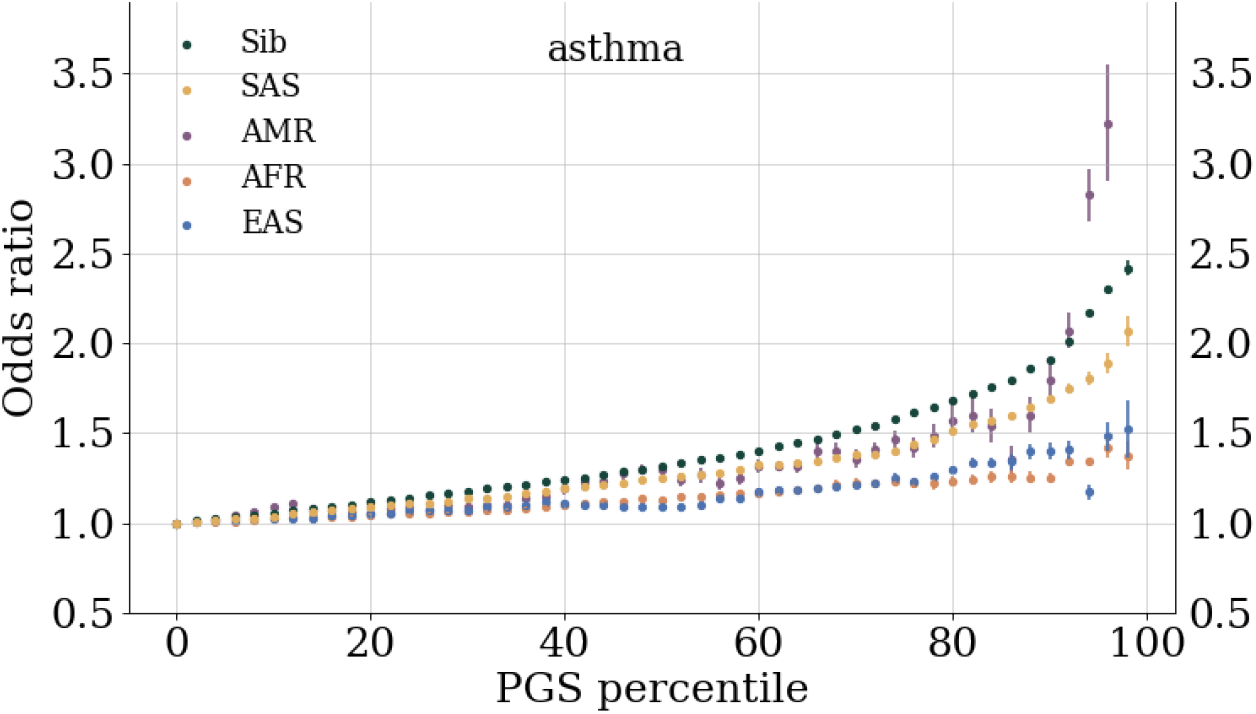
Inclusive odds ratio (OR) for asthma. The inclusive OR is the ratio of all cases to controls *at a given PGS or above* normalized to the ratio of the total number of cases to controls. At the highest PGS bins, data is omitted if there are no cases or controls. Similar plots for the other phenotypes and details about how uncertainties are computed are all located in the Supplementary Information.

### 2.2 Biobank Projections

This section details how training at various sizes in current biobanks can be used to model the growth dependence on training size. For most clinically relevant metrics, e.g., correlation and AUC below, this growth can be modeled with uncertainty. Using relatively simple parametric functions, we find that we can model the growth of these metrics. The results can be used to guide future studies and genotype-phenotype database construction by identifying where large gains can be made.

For sparse methods like LASSO, it has been previously shown that using self-reported ancestry performs similarly to principal component based clustering when based on AUC and Correlation metrics [10]. Nonetheless, after sorting on self-reported ancestry, we perform an additional regression on the top 20 PCs to adjust for any remaining population stratification.

To be conservative in the projections for PRS performance in other biobanks, several modeling assumptions are made. First, to be conservative we assume that these biobanks will have population prevalence rates for diseases even though some biobanks over-recruit cases to enrich their datasets. Additionally, the actual incidence rate for disease conditions fluctuates over time. For the conditions considered in this paper we try to consider the most recent surveys of all ages. Finally, when there are various estimates for disease prevalence within ancestry sub-groups, we choose a conservative (i.e. low) prevalence as the representative for the overall ancestry prevalence. Specific details about within ancestry prevalences are given in the Supplementary Information.

In **Figure 4** we see projection bands for asthma and BMI. The bands grow from no signal (0.5 AUC and 0 correlation respectively) to asymptotic values. The various colored prediction bands correspond to the Monte Carlo (MC) confidence intervals for the various fit functions as described in section 3. The asymptotic predictions for each trait can be averaged, incorporating the confidence intervals, and the results are given in **Table 2**. Further projection plots for the remaining traits can be found in the Supplementary Information. The fraction of phenotypic variance captured by SNPs (i.e., the linear, narrow sense heritability) of traits is traditionally estimated via means such as Restricted Maximum Likelihood estimates (REML) and using Linkage Disequilibrium Score Regression (LDSR). For continuous traits, the correlation of the residual phenotype with the PGS can be related to the linear SNP heritability *explained by the predictor* by simply squaring the correlation. In **Table 2** this can be seen in the final column. The heritability explained by the predictor is a *lower bound* on the REML heritability in that there may not be enough data to saturate the REML estimates. In **Figure 4** we can see the heritability estimates from GCTA (using REML) and LDSR converted to a correlation scale. For traits like BMI and height it appears that, eventually, sparse predictors will capture all the linear SNP heritability. For much sparser conditions, e.g. Lipoprotein A presented in the Supplementary Information, it appears that sparse methods are out performing traditional measures of heritability.

**Figure 4:**
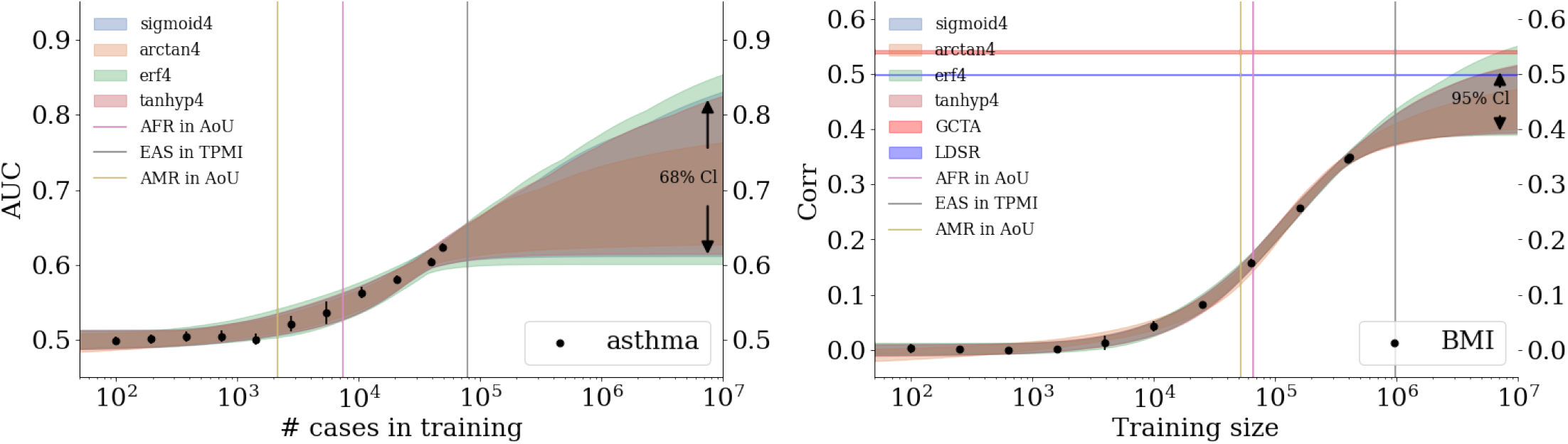
Growth of AUC (left: asthma) and correlation (right: BMI) as a function of training size in the UKB. Colored, curved bands come from fitting data with various 4 paramter functions. Width of the band corresponds to a confidence interval on the predictions: on the left 2 standard deviations or ∼68% and on the right 4 standard deviations or ∼95%. Vertical bars represent projections for de novo training in other biobanks using literature prevalences, summarized in the Supplementary Information. On the right, horizontal lines indicate the correlation predicted from GCTA and LDSR.

**Figure 5:**
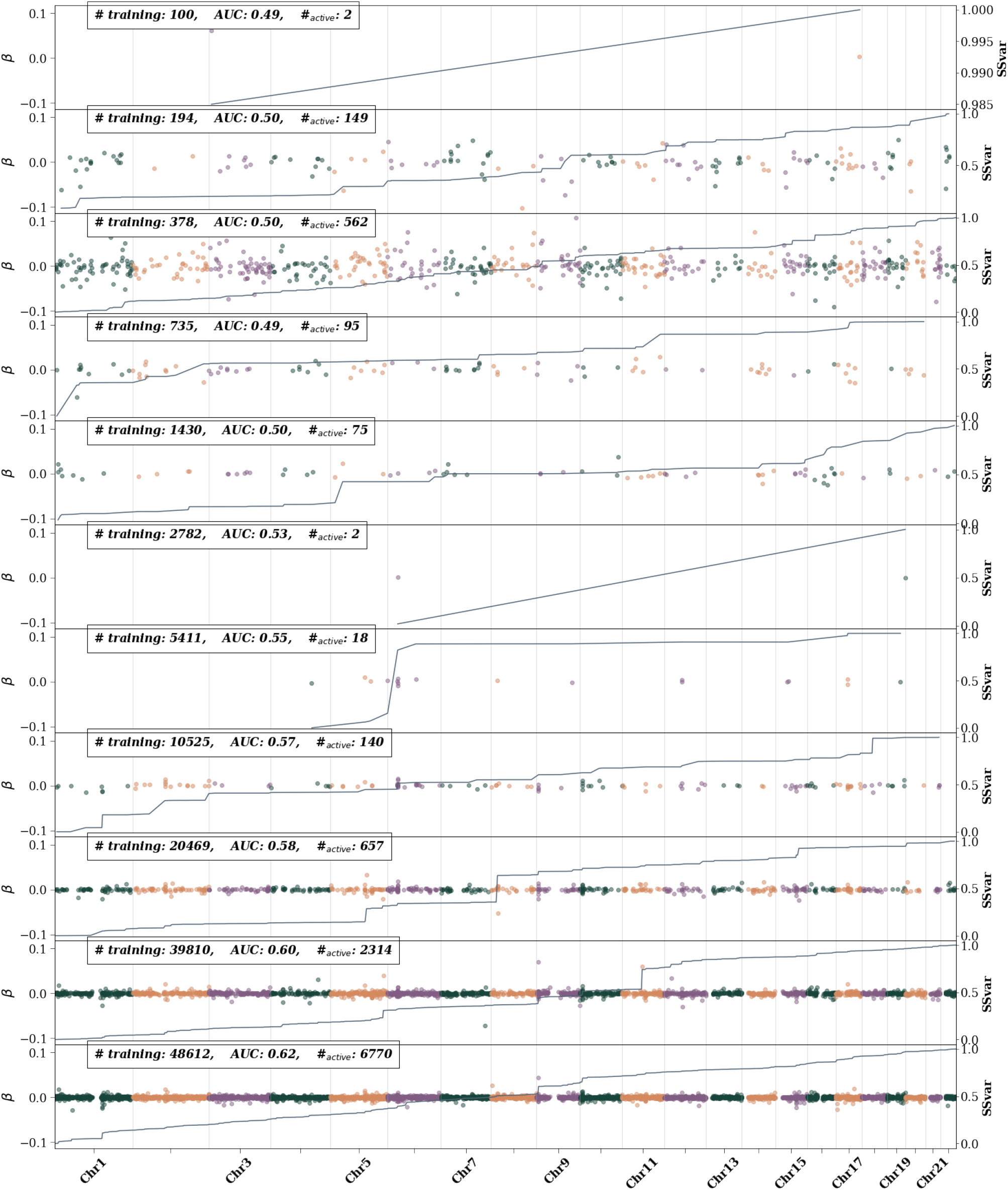
Asthma active SNPs – i.e., SNPs with non-zero *β* weights– as training size is increased. The left axis shows the *β* value and is represented by colored dots. Different colors are used to differentiate chromosomes. The right axis represents the single SNP variance (SSV) normalized to the total SSV. The solid line showes the cumulative SSV. The “training” label represents the number of cases used in training. The first 10 (from the top) training sizes use equal number of cases and controls. The final training size uses all possible remaining controls.

**Table 2:** Asymptotic projections for AUC and and correlation for case-control and continuous traits respectively. Type 1/2 diabetes (T1/2D), coronary artery disease (CAD), and body mass index (BMI) are all abbreviated to save space. “Gain” represents the additional gain over the best result from training reported here. For most case control conditions, except T1D, there are large gains that can be found from increased training sizes. For continuous phenotypes, BMI can benefit from training on larger data sizes. Correlations are also translated to the asymptotic projection of linear, narrow-sense SNP heritability explaind by the predictor. Colors correspond the relative sparsity of the predictor as mentioned in Table 1.

| trait | $AUC_{(std)}^\infty$ | $gain_{(std)}$ | trait | $corr_{(std)}^\infty$ | $gain_{(std)}$ | linear $h_{(std)}^{2\infty}$ |
| --- | --- | --- | --- | --- | --- | --- |
| asthma | 0.71<sub>(0.03)</sub> | 70<sub>(5)</sub>% | direct bilirubin | 0.53<sub>(0.02)</sub> | 5<sub>(4)</sub>% | 0.28<sub>(0.02)</sub> |
| atrial fibrillation | 0.82<sub>(0.06)</sub> | 180<sub>(10)</sub>% | BMI | 0.47<sub>(0.02)</sub> | 34<sub>(6)</sub>% | 0.22<sub>(0.02)</sub> |
| T2D | 0.83<sub>(0.06)</sub> | 150<sub>(10)</sub>% | height | 0.690<sub>(0.005)</sub> | 10<sub>(1)</sub>% | 0.476<sub>(0.007)</sub> |
| T1D | 0.668<sub>(0.006)</sub> | -0.01<sub>(0.03)</sub>% | lipoprotein A | 0.779<sub>(0.009)</sub> | 3<sub>(1)</sub>% | 0.61<sub>(0.01)</sub> |
| CAD | 0.82<sub>(0.06)</sub> | 170<sub>(10)</sub>% |  |  |  |  |
| hypertension | 0.635<sub>(0.005)</sub> | 31<sub>(1)</sub>% |  |  |  |  |
| breast cancer | 0.75<sub>(0.06)</sub> | 110<sub>(50)</sub>% |  |  |  |  |

### 2.3 Sparse Output Interpretation

An advantage of sparsity is that, because there are fewer features, it is relatively easier to categorize features compared to non-sparse methods. Here we identify important features for predictors for each trait. Because a simple LASSO routinely performed as one of the best predictors in terms of AUC and correlation in section 2.1 we focus here on interpreting the LASSO outputs.

In **Figure 5** we see an example of the SNP content for an asthma predictor as it is trained with larger and larger training sizes. The LASSO weights *β*, single-SNP-variance (SSV), and training sizes are all further described in section 3. A notable feature from this figure is that, as training size is increased, the LASSO algorithm first adds more SNPs (becomes less sparse) while barely increasing the AUC. Eventually the algorithm gets rid of most of these SNPs, becomes much more sparse, and then quickly increase AUC as SNPs are then added again. This is an example of the algorithm “searching” for seemingly important features. Once some of the important features are identified, the algorithm finds more and more features that are important as evidenced by the rising AUC. Note that, as described in section 3, LASSO does not try to directly optimize AUC. This sparsity behavior (rising and falling before eventually finding important features that greatly increase AUC/correlation) appears in the rest of the predictors seen in the Supplementary Information and appears to be related to the Donoho-Tanner phase transition (see [59] for an explanation of this transition in the context of genomics). This behavior can be seen in other features of each predictor – e.g., the SSV – as displayed in Supplementary Information.

At the largest training sizes – i.e., after the phase transition – we also look at how the SNP content varies across CV folds. As detailed in section 3, for case control phenotypes the largest training size includes as many controls as possible while all previous training sizes contain an equal number of cases and controls. In **Figure 6** and **Figure 7** we see examples of the SNP content for these two training sizes for asthma. Analogous plots for the other phenotypes can be found in the Supplementary Information. For the largest training sizes we can also average over the CV folds to find the fraction of SSV per chromosome. An example can be seen in **Figure 8** where the error bars describe the variation over folds (the plots for other phenotypes are in the Supplementary Information). There are several important takeaways from these results. The main metric for case-control phenotypes is much more affected by the number of cases than controls. Specifically, increasing the number of cases used in training increases the AUC, but increasing the number of controls in training, even by a factor of 2 or more, either does not change the AUC or leads to a very minor change. However, when looking at the single SNP variance, the number of both cases and controls do have appreciable effects. Take for example asthma, seen in **Figure 6** for 5 cross-validation folds with the maximal number of cases *and* controls, and seen in **Figure 7** for 5 cross-validation folds with near maximal number of cases *and an equal number* of controls. If we look for common features among the folds, in **Figure 6** we see a modest jump in SSV at specific locations on chromosome 1, 2, 5, 6, 9, 10, 11, and 17. Additionally the increase in SSV on chromosome 17 is much larger on a single fold. In contrast, **Figure 7** shows impact regions at similar regions, but the fraction of SSV at the beginning of chromosome 9 is much larger and the effect on chromosome 17 seem to be more smoothed out. This is also reflected in **Figure 8** where the SSV fraction is averaged per chromosome and contrasts these two cases.

**Figure 6:**
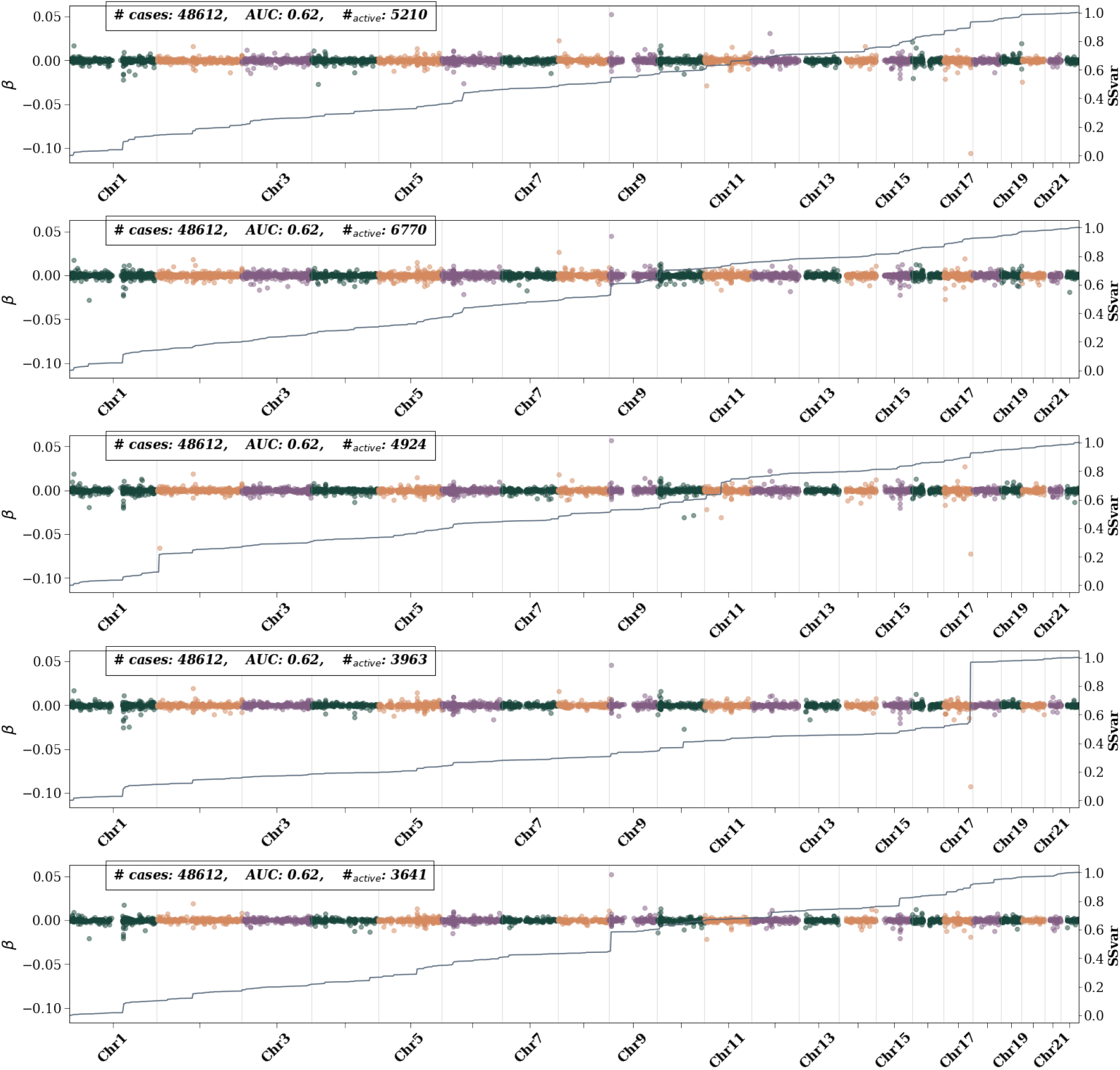
Asthma active SNPs – i.e., SNPs with non-zero *β* weights– for 5 CV folds at maximum training size. Left axis shows the *β* value and is represented by colored dots. Different colors are used to differentiate chromosomes. The right axis represents the single SNP variance (SSV) normalized to the total SSV. The “training” label represents the number of cases used in training. All possible controls were used in each fold. While features generally appear consistent across folds, i.e., the presence of a bump in the SSV line, the size of the bump varies.

**Figure 7:**
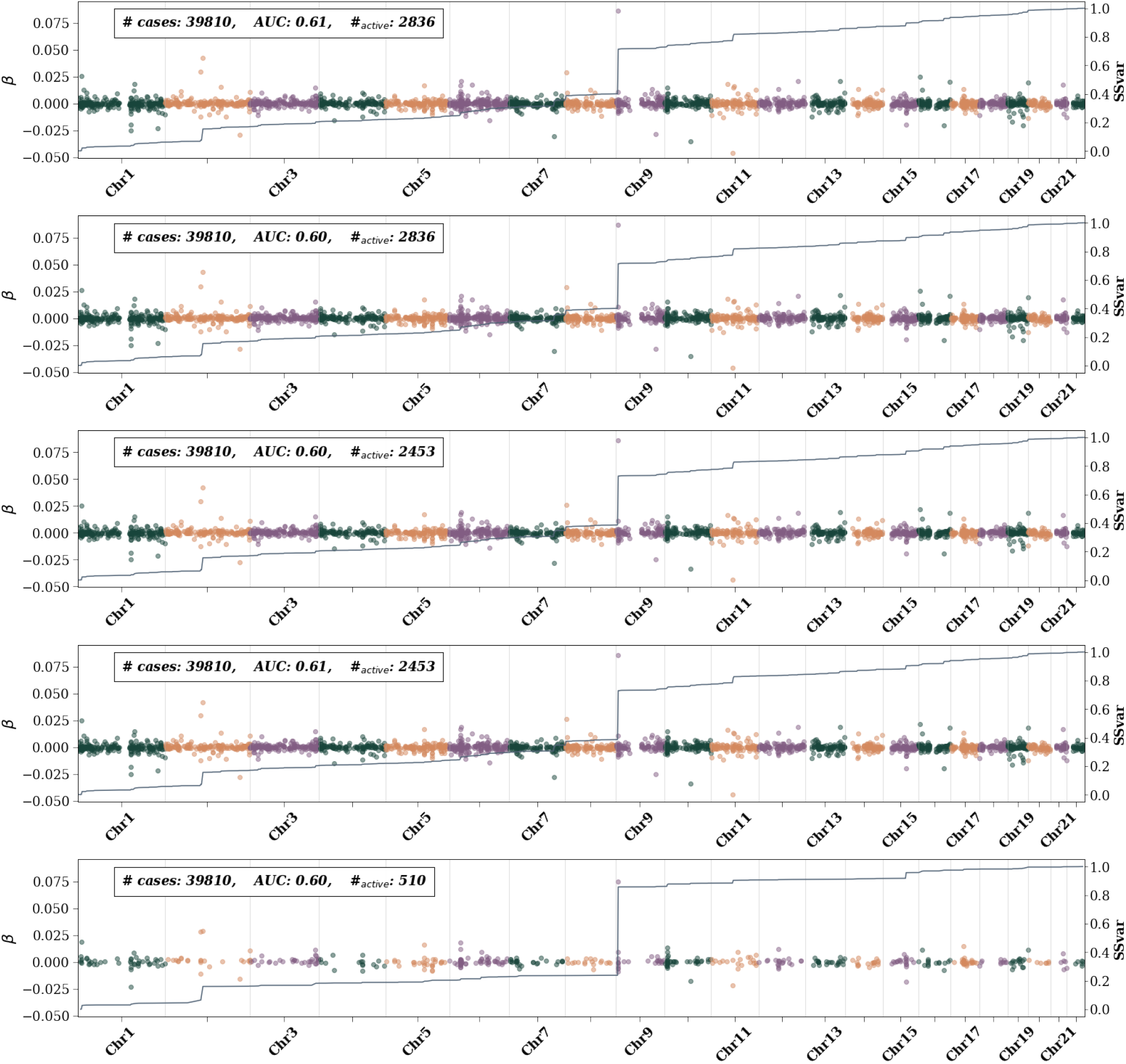
Asthma active SNPs – i.e., SNPs with non-zero *β* weights– for 5 CV folds at near-maximum training size, but with equal cases and controls. Left axis shows the *β* value and is represented by colored dots. Different colors are used to differentiate chromosomes. The right axis represents the single SNP variance (SSV) normalized to the total SSV. The “training” label represents the number of cases used in training (an equal number of controls also used). Compared to maximal training in Figure 6, the features here are much more consistent across folds.

**Figure 8:**
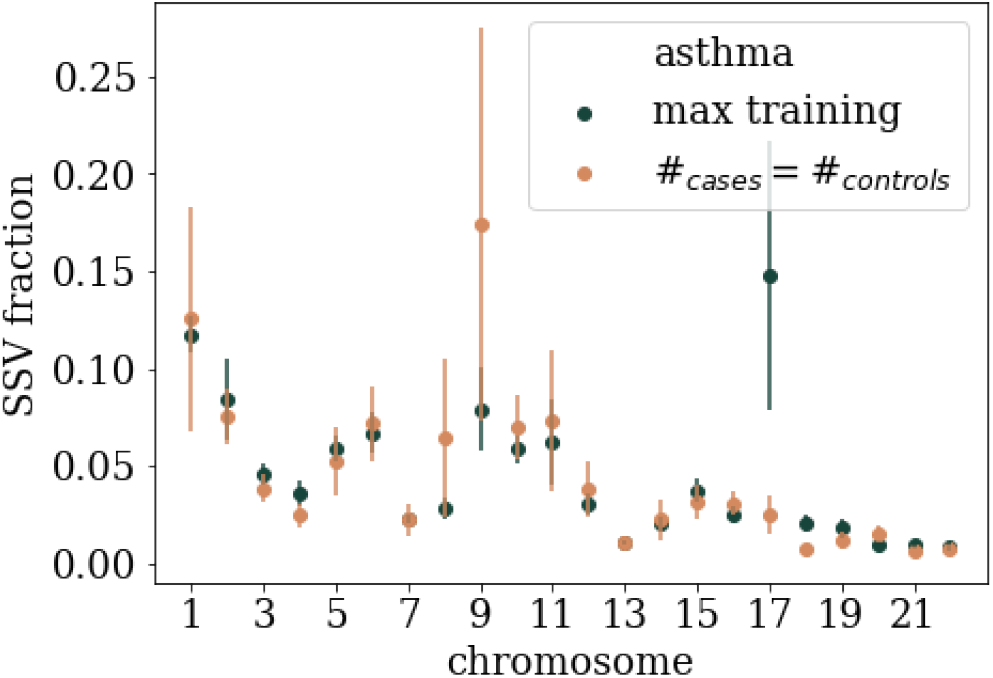
Average SSV per chromosome for asthma. Uncertainty comes from averaging over 5 fold cross validation. Max training refers to using the maximum number of cases and all possible controls. #*_cases_* = #*_controls_* uses near maximal number of cases and an equal number of controls. Both types of training find generally similar SSV distribution, but max training finds a much larger signal and chromosome 17 and a smaller signal on chromosome 9.

To interpret these plots we have collected and identified every SNP that accounts for at least 1% of total SSV. These tables are found in the Supplementary Information. Here we review the most important SNPs and compare them to known results, i.e. we identify genes, associated with the particular phenotypes, that are within at least 2 million base pairs of a SNP accounting for *>* 1% SSV. We choose a 2 million base pair distance to be a conservative measure of possible long-range LD[60]. All predictors, except those for Lipoprotein A, include SNPs that account for at least 1% of total SSV but are not near any known gene that associates with the phenotype. Additionally, we list the p-value associated with a GWAS on the raw (case control) or adjusted (continuous) phenotype to highlight that many of these important LASSO SNPs would be missed by a traditional GWAS approach.

SNPs which we identify that are located near known associated genes for several phenotypes are listed here:

- Asthma: SNPs around FLG, IL1RL1, TSLP, IL33, SMAD3, HLA-DQ, RORA, CLEC16A, and SERPIN7 which have been previously been identified by GWAS [61–65]
- Atrial fibrillation: at least 164 SNPs have been identified in GWAS studies [66–69], and while none of these exact SNPs appear in the 58 SNPs identified using LASSO, many of the LASSO SNPs are located around the genes KCNN3, PMVK, LMNA, KIFAP3, PRRX1, SCN5A, SCN10A, PITX2, FAM13B, WNT8A, CAV1, SH3PXD2A, HCN4, ZFHX3, RPL, and FBXO32 which are associated with some of the gwas SNPs
- Breast cancer: none of the SNPs identified here are near the genes identified in [70]
- Type 2 Diabetes: we find relevant SNPs in the associated genes GCKR, TCF7L2, and SLC12A1[71]
- Type 1 Diabetes: we find SNPs near HLA-A, TRIM26, MICA, HLA-DRB1, and LAT[72]
- Coronary Artery Disease: we find SNPs near the associated genes PCSK9, PLPP3, IL6R, MIA3, VAMP5, ZEB2, SLC22A4/A5, SLC22A3, LPAL2, LPA, PLG, and CETP[73, 74]
- Hypertension: we find important contributions near the ULK4, NR3C2, PRRC2A, and NOS3 genes[75]
- Direct Bilirubin: we find results near the UGT1A1, SLCO1B3, and SLCO1B1 associated genes [76, 77]
- Body Mass Index: we find SNPs in genes TMEM18 and the well studied FTO - both appear with more than 1% SSV for BMI [78]
- Height: Liu et. al. [79] categorized over 400 genes associated with height that were later reanalyzed by Yengo et. al [80]. Of these, SNPs near ORC1, COL11A2, FANCE, BRAF, ACAN, ANKRD11, CDK10, CDT1, FANCA, GALNS, and RPL13 all appear in our analysis
- Lipoprotein A: all SNPs appear in or near the genes LPA, LPAL2, and SLC22A3 which are all known to be associated with Lipoprotein A levels [81, 82] and additionally contribute to coronary artery diseases [83].

There are several interesting aspects of examining SNPs in this manner. First we note that we are interested in *common* variants and exclude SNPs with a minor allele frequency below 0.01 to avoid any spurious associations. Because of this, rare variants can’t appear in our analysis, even if they are known to be associated with a phenotype. An example of this can be seen in the case of breast cancer where the BRCA mutations aren’t included in our analysis. Interestingly there are SNPs that have previously been identified via GWAS, that are available on our array, but are *not* selected by LASSO. An example would be rs116716490 which is part of the ZBTB10 gene, previously associated with asthma via GWAS [63], but not selected by LASSO.

A much more coarse grained interpretation of the impactful regions can be found in **Figure 8** and in the Supplementary Information. Here, we examine–for each trait–the fraction of SSV that resides on each chromosome. For case-control phenotypes, we also compare the result for max possible training (i.e., the max possible cases *and* controls), and training with the largest possible equal number of cases and controls. We highlight some of the notable results. For atrial fibrillation, max training and equal case control training both find a large fraction of SSV on chromosome 4, but the signal is much larger for equal cases and controls. On chromosome 15 there is a large signal for max training, but not for equal cases and controls. For type 2 diabetes a large fraction of SSV is on chromosome 10, but the signal is largest for equal cases and controls. For type 1 diabetes. equal cases and controls find a strong signal on chromosome 6 while max training also finds signals on chromosomes 1, 11, 15, and 17. For CAD, the equal case control training finds the largest signal in chromosome 6 while max training finds similarly large signals in chromosomes 1, 2, 3, 6, and 12. Hypertension shows varying signals all throughout the chromosome with the most precise signal coming from chromosome 1. Breast cancer has the most diverse differences between the two types of training with large signals for equal case control training on chromosomes 10 and 16, and large signals on 7, 11, 16, and 19 for max training. For continuous phenotypes we only have one measure of SSV per chromosome. For direct bilirubin the largest contributions are on chromosomes 1 and 2. For BMI the largest signals are on chromosomes 1, 2, and 3. For height there is strong signal throughout most of the genome. Finally for Lipoportein A, the signal seems to be concentrated on chromosome 6.

### 2.4 Sparsity and Heritability

We can see examples of all the definitions of sparsity in **Figure 9** where the definitions themselves are explaind in section 3.3. The traits are roughly grouped according to their heritability estimate using GCTA [84]. Using all metrics together, the scatter of data can be regressed linearly in *log*_10_ scales in both training size and sparsity. We find *log*_10_(*s*) = 0.7_(0.1)_*log*_10_(*N*)+0.1_(0.5)_ (the black line in **Figure 9**) with an *r*^2^ = 0.43. This weakly implies *s*∼*N* ^0.7^ (or conversely *N*∼*s*^1.4^). In [59] it was shown, using simulated data, that for *h*^2^ = 0.5 (where *h*^2^ represents the narrow-sense heritability) the compressed sensing phase transition occurs at *N≈*30*s*. The result reported here is consistent with this previous prediction, but tighter error bars are required to completely determine the coefficient and its dependence on *h*^2^.

**Figure 9:**
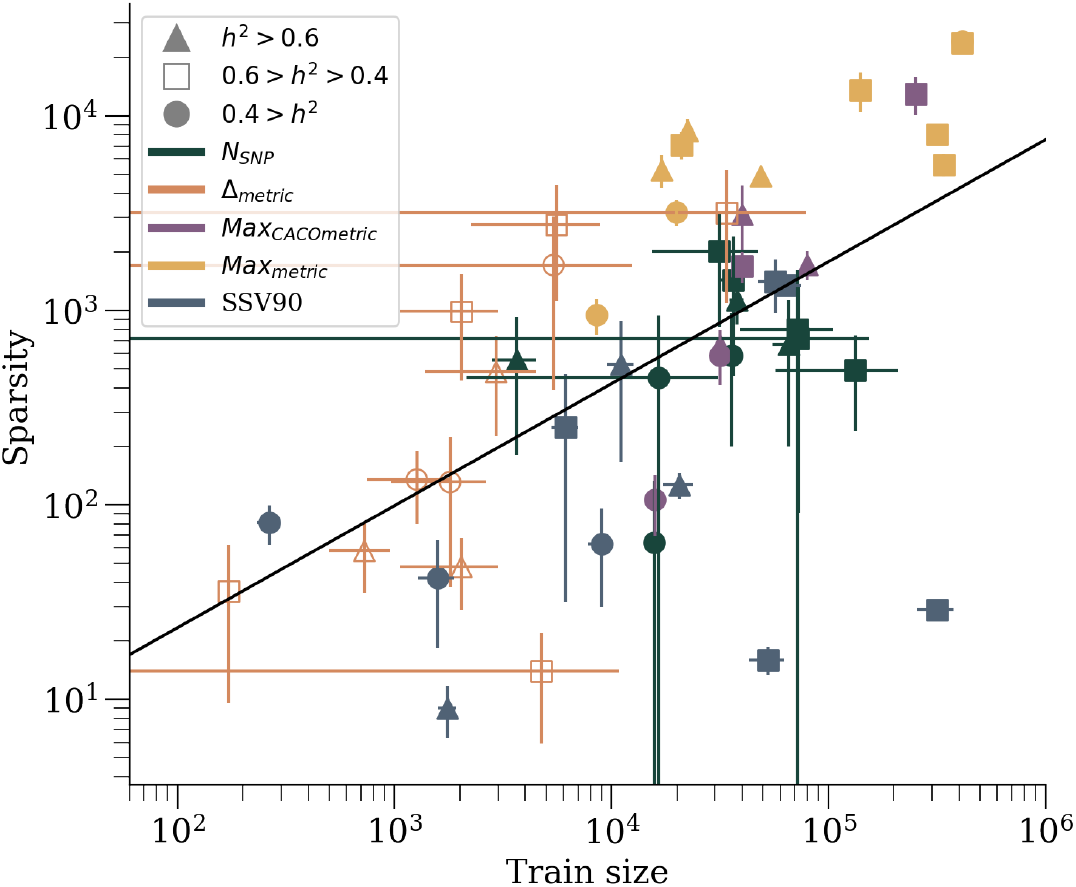
Sparsity measurements, as a function of training size, for all 11 traits. Different markers correspond to different (arbitrary) estimated heritability groupings. Different colors correspond to different versions of sparsity. Heritablity here for case-control phenotypes is *broad sense* heritability reported from twin/family study literature, whereas GCTA was used to estimate heritability for continuous phenotypes. Low heritability traits (circles) include: atrial fibrillation, breast cancer, and BMI[85, 86]. Medium heritability traits (squares) include: CAD, hypertension, direct bilirubin, height, and lipoprotein A[87, 88]. High heritability traits (triangles) include: asthma and type 1/2 diabetes[89–91].

Finally we can see estimates of heritability using a variety of metrics in **Figure 10** and in the Supplementary Information. In **Figure 10** we see examples of these heritability estimates, from a LASSO training, as a function of training size. (Additional phenotypes are in the Supplementary Information). Additionally, for the largest training size, we also record these heritability estimates for the other training methods. As a function of training size we generally see the phenotype dependent phase transition behavior noticed before. After a critical amount of training data is used, the error bars for all methods greatly decrease. Identifying this transition behavior is important as it indicates looking at central values *below* the critical training amount gives a deceptively high estimate. For most phenotypes–regardless of training size, metric, or method–the estimated heritability is below what can be estimated from GCTA. However, there are a few exceptions: for asthma and type 1 diabetes, at the largest training size PRS-cs using the ▴ metric outperforms GCTA; for height the metric generally, and PRS-cs at largest training, outperforms GCTA; and for Lipoprotein A, every metric and method *except* PRS-cs results in a higher value than GCTA.

**Figure 10:**
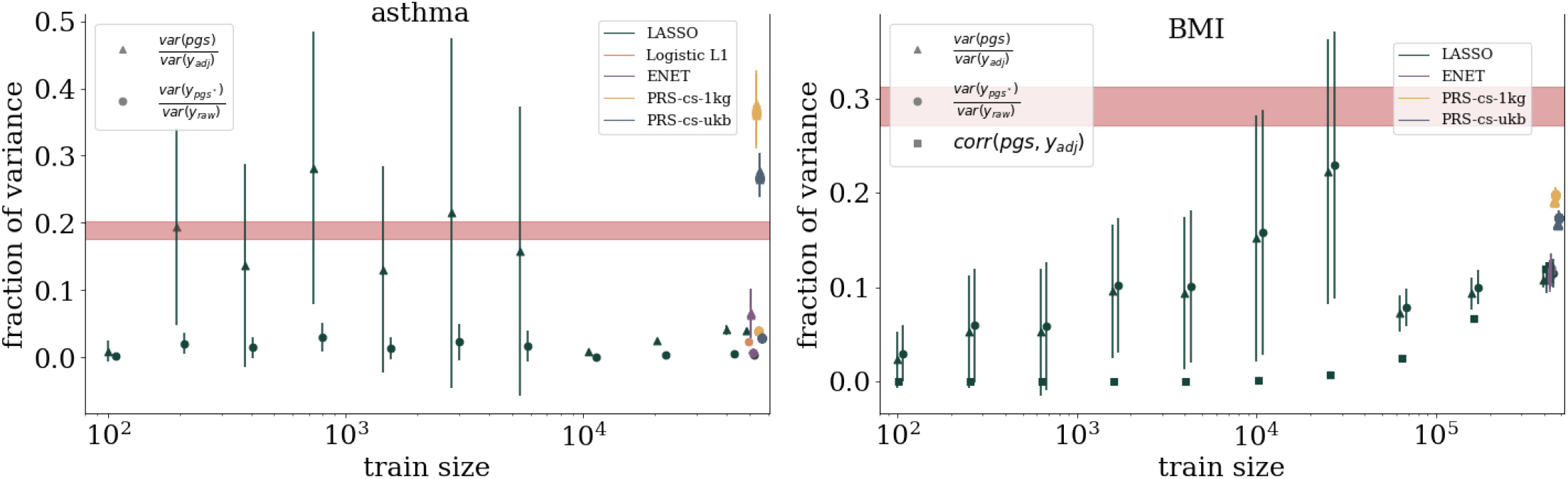
Estimates of the fraction of variance explained from *purely genetic* contributions for asthma and BMI. There are various ways to estimate the variance explained as explained in appendix G. Similar plots for the other traits are found in the Supplementary Information.

## 3 Methods

### 3.1 Predictor Training

We start with a very brief description of the general training pipeline for generating predictors: **step (1)** initial populations are separated into groups; **step (2)** quality control (QC) for both phenotypes and genotypes; **step (3)** phenotypes are regressed on covariates and adjusted phenotypes are built; **step (4)** SNP set is filtered down to a computationally manageable size; **step (5)** machine learning on adjusted phenotype and genotypes using cross-validation; **step (6)** predictors are tested on withheld groups.

As an initial grouping of UKB data, we separate participants using self-reported ancestry (a principal component adjustment is done below). We use participants reporting ancestry as White (EUR), South Asian (SAS), East Asian (EAS), and Black (AFR). Additionally we construct an American ancestry (AMR) set done using the approach from [55] and described in the Supplementary Information (there is a small 60 person subset of the 322 AMR labeled participants who also self identify as white and could appear in the training. This set is so small that its effects are assumed to be negligible). From the EUR population we identify genetic siblings as described in A.2 and remove them to use as a final testing set. The remaining EUR population is used for training.

Step 2 involves performing quality control on both the genotypes and phenotypes. Full phenotype definitions, including UKB codes, are given in Supplementary Information. For phenotype quality control we exclude any missing values or negative values (usually used as a placeholder or indicator in the UKB). For continuous phenotypes we average over all recorded measurements (many participants are measured on repeated visits, but there is not a consistent number of visits per participant). Case control conditions are defined with a logic ’or’, i.e., if the participant is recorded as a case for *any* relevant code, then they are counted as a case. The UKB array contains 805,426 SNPs. We run QC using PLINK to filter out (remove) variants (SNPs) with more than 3% missing values, samples (participants) with more than 3% missing values, and variants with minor allele frequency less than 0.001 (i.e. 0.1%). After QC this leaves 663,533 SNPs and 487,048 participants. (Exact number of participants can slightly vary due to participant withdrawal from the UKB program.) Finally we reduce our SNP set down to only the autosome as this allows us to roughly double our training base, i.e. use all sexes in training (except for sex specific phenotypes).

Step 3 can be briefly described as sex-specific z-scoring and covariate adjustment. The z-scoring is only done on continuous phenotypes while covariate adjustment is done for all phenotypes. We z-score to improve the efficiency of the machine learning algorithms (e.g., using normalized data lowers the risk of large numbers appearing in a gradient descent algorithm). If we assume that we are looking for common genetic factors that are independent of sex then we can z-score *each sex individually* and roughly double our training data. The ultimate aim is to identify genetic variants that we are most confident are related to a phenotype. To do this we assume that common covariates and population stratification *have a maximal effect*. That is, we regress covariates on the raw (or z-scored) phenotype and then adjust (i.e., create a residual phenotype) phenotypes for the contribution explained by these covariates. Common covariates included are: age, sex (except for sex specific traits like breast cancer), and the top 20 principal components as computed by the UKB. Even though we do a sex specific z-scoring, at this stage we still assume sex can have an impact. That is, for phenotype, 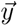, and covariates, H̄, we regress: 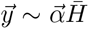. Then we can construct an *adjusted phenotype*, 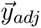, that just includes the residual signal: 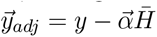.

Unfortunately, before we employ a machine learning algorithm, we have to reduce down the ~ 600*k* SNPs to a computationally manageable number. The exact computational details will depend on the machine being used to run the analysis and whether or not computational cost saving measures can be used (e.g., parallelization). For our analysis, as shown in the Supplementary Information, the time to run lasso scales as a function of training data, *N*, roughly as ∼ *N* ^1.35^. Additionally, the memory and CPU usage, while growing more slowly than exponential, grows quickly. For step 4, we choose to subset our SNP set by selecting the top 50k SNPs (10k for penalized logistic regression which is more computationally intensive) via GWAS with the training set.

Next, in step 5 we run machine learning algorithms. For LASSO and Elastic Net we use the Scikit-learn[92] lasso path linear_model.lasso_path and enet path linear_model.enet_path algorithm. The Elastic Net algorithm minimizes the objective function,

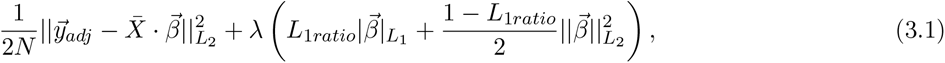

where X̄, the normalized genotype matrix, 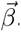 the regression weights, *λ* the hyperparameter, *L*_1*ratio*_ relatively weights the *L*_1_ and *L*_2_ penalties. As *L*_1*ratio*_*→*1 this becomes LASSO and *L*_1*ratio*_→0 it becomes ridge regression. For all phenotypes we run for 5 different ratio weights: *L*_1*ratio*_∈.1*, .*3*, .*5*, .*7*, .*9 . For *L*_1_ penalized logistic regression we use the Scikit-learn function linear_model.LogisticRegression with an L1 penalization and parallelize it using the Multiprocessing package Pool function. For all methods of (penalized) regression we use five-fold cross validation and use a 2,500 sample validation set (withheld from training) for hyperparemeter selection. For case-control phenotypes, the validation set is equal number of cases and controls. Finally, we also use PRS-cs[93] which is a Bayesian shrinkage prior which runs directly on summary statistics (i.e., GWAS output) and an LD matrix. We use three fold cross validation and run with the global shrinkage/sparseness parameter, *φ* ∈ {10^−1^, 10^−3^, 10^−5^, 10^−7^, 10^−9^} and the local scale parameters {*a, b*} ∈ {1*/*2, 1, 3*/*2}.

In the final step 6 we apply the predictors to all testing sets: the EUR genetic siblings set, an SAS set, an EAS set, an AFR set, and an AMR set. Case control phenotypes are evaluated by computing the Area Under the receiver operator Curve (AUC) which compares the true positive rate to the false positive rate, i.e. how well the predictor is calling cases and controls. Within this work we generally report the AUC *only using the genetic weights* to emphasize how well cases and controls can be identified using only genetic information. For case control phenotypes we can also compute an odds ratio, as a function of polygenic score, as seen in **Figure 3**. This is an *inclusive* odds ratio: at each score, we compute the ratio of the number of cases to controls (normalized to the total cases and controls) of all individuals with that score or a higher score. I.e., the 80th percentile represents all individuals with a score ≥ 80%. For continuous phenotypes we simply report the correlation between the polygenic score and the adjusted phenotype, 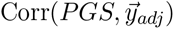, and again this estimates the *purely genetic* contribution. For both types of phenotypes there are sibling specific tests that we perform. These were developed in [57, 94]. Siblings generally share a similar environment growing up which helps to control for some external factors. For case control phenotypes we can consider Affected Sibling Pairs (ASPs), which are a pair of siblings where one person is a case and the other a control. We can then look at the correct selection rate, that is the fraction of the time the person with the larger polygenic score corresponds to the case. This computation can be done again while requiring that the sibling pair’s scores are at least 1.5, 2, or 2.5 standard deviations different. For continuous phenotypes we show a similar selection rate for the amount of time the person with the larger polygenic score has the larger phenotype. Again this rate can be recomputed with the requirement that the phenotype difference between the siblings is at least 0.5, 1, or 1.5 standard deviations.

In addition to standard metrics like AUC and correlation, we are often interested in the amount of variance described by a particular polygenic predictor. For a particular polygenic score, the variance is described by

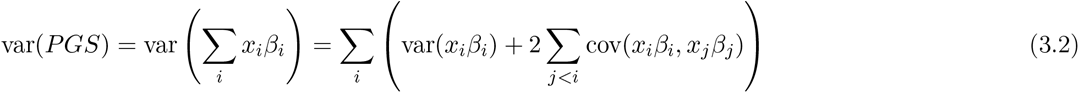

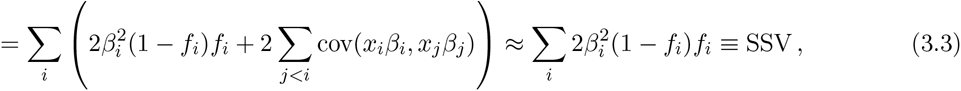

here the approximation defines the Single SNP Variance (SSV). This approximation assumes that the covariance between SNPs is small. For most sparse methods, this covariance is minimized to enforce sparsity. For example, in the objective function eq. (3.1), adding extra SNPs with non-zero weights amounts to decreasing the first term and increasing the second term. For an added SNP that is highly correlated with another SNP with non-zero weight, the decrease in the first term is likely smaller than the increase in the second. The accuracy of this approximation is demonstrated in Supplementary Information.

### 3.2 Metric Projection

We can model improvement in predictor performance metrics as a function of training data size. In **Figure 4** we see examples of this for asthma and BMI. We use four functions, which have left and right asymptotes, to model this growth: sigmoid, inverse tangent, error function, and hyperbolic tangent. The hyperbolic tangent can be written as a rescaled version of a sigmoid function. We use that as a cross-check, i.e. we make sure both functions give the same results, to avoid fitting routines getting stuck in a local minimum. For all functions, four parameters are used to fit the performance, e.g., erf4(*x*; *a, b, c, d*) = *a* + *b* erf(*c*(*x* + *d*)). Care is taken to incorporate uncertainty from cross-validation and finite data sizes. More details about the functions, uncertainty calculations, and fitting results are given in the Supplementary Information. The non-linear fit of each functional form involves computing a Hessian matrix as a function of the parameters *a, b, c, d* . The inverse of this matrix is the correlation matrix for the model’s fit parameters. Using these empirical correlations we build a MC method that relies on a Cholesky decomposition. For a given correlation matrix *<u>C</u>* of fit parameters, we can generate a correlated set of MC parameters, 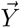, from random numbers, 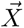, via a Cholesky matrix, *<u>L</u>*:

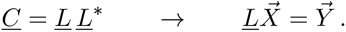

Subject to physical constraints – 0.5 *<* AUC *<* 1 and 0 *<* Corr *<* 1 (positive correlation is a convention choice) – we can then build MC bands as seen in **Figure 4**. For both types of phenotypes, these projection bands are informative: they indicate **(1)** the asymptotic (i.e. best possible) metric obtained from infinite data, as in **Table 2 (2)** which traits are nearing that asymptotic maximum or which are still improving substantially with more training data, and **(3)** what sample size other biobanks will need to obtain meaningful results. These projection bands can be compared to the application of European trained predictors applied to other ancestry groups. In **Figure 1** we see that predictors built using data from TPMI and AoU will greatly surpass the results of UKB trained predictors when applied to distant ancestry groups.

### 3.3 Sparsity

The sparsity associated with a phenotype can be defined in *several* ways and it is not *a priori* clear which definition is most appropriate. The definitions used here are:

1. The number of SNPs with non-zero weights using all possible cases and controls. This quantity can appear to continue to grow without reaching an asymptote even though traditional metrics (e.g. AUC or correlation) appear to asymptote. However, this definition is obviously impacted by training sample size and difficult to compare between datasets/biobanks. Assuming infinite data generated by a linear model plus noise, compressed sensing guarantees complete signal recovery and the sparsity will reach an asymptotic value corresponding to the underlying model that generated the data [59]. Additionally, for algorithms like LASSO there is debate as whether to use the maximal metric for hyperparameter selection or to step one standard deviation “back” (i.e. towards fewer features) in hyperparameter space to avoid over-fitting when applying to outside true testing sets. (see [10] and references therein).
2. We can use the previous definition but for the training case with the maximum number of equal cases and controls. The same caveats from definition 1. apply.
3. Because we expect traits not to be 100% heritable, we expect metrics to asymptotically approach a limit value. We can look at relative increase in these metrics to try to identify the point of largest growth (or above some cut), i.e. the inflection point. We can do this by looking at a *relative metric*, i.e. (*y_i_*_+1_ − *y_i_*)*/y_i_*. We can look at the sparsity of the predictors at this point as a second version of sparsity.
4. Instead of looking at the metrics, we can instead look at the relative increase in the number of features (SNPs) at each training size and look for the relative maximum increase (or increase above some cut) in features to define a sparsity value.

Comparisons for these definitions can be seen in **Figure 9**.

### 3.4 Fraction of Variance Explained

Heritability, *H*^2^, is traditionally defined as the proportion of phenotypic variance explained by genetic factors. All the work discussed here involves linear genomic prediction, i.e. that 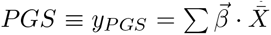. The proportion of variance explained by a linear model is generally referred to as the narrow-sense or linear heritability, *h^2^_linear_*. There are several ways we can estimate the fraction of variance explained via the results of a linear predictor. First, we note that our linear predictors are trained after sex specific z-scoring. Because of this, the overall scale of *y_P GS_* (i.e. the variance) is expected to be of the order of the adjusted phenotype, *not* the original raw phenotype. Therefore, we can consider the ratio of the variance of the PGS to the adjusted phenotype (▴ denotes *var*(*pgs*)*/var*(*y_adj_*)) or we can undo the sex specific z-scoring only on the PGS and compute the ratio of variance in this rescaled PGS to the raw phenotype (● denotes *var*(*y_p_*^∗^*_gs_*)*/var*(*y_raw_*)). Finally, for continuous traits we can simply look at the correlation between pgs and adjusted phenotype as in [10, 51, 56].

In both section 2.2 and in section 2.3 we refer to the heritability estimated via REML and LDSR. The LDSR results are reported from [95, 96]. The REML results are produced using the GCTA software[84, 97]. All GCTA computations used 350,000 SNPs and the following number of samples: asthma – 20,469; atrial fibrillation – 9,027; type 2 diabetes – 11,077; type 1 diabetes –1,755; CAD – 11,077; hypertension 11,659; breast – ; direct bilirubin – 21,544; BMI – 25,118; height – 21,544; lipoprotein A – 21,544.

## 4 Discussion

Sparse methods optimize prediction while activating as few features (SNPs) as possible during training. In contrast, non-sparse methods construct predictors in which potentially every SNP in the genome has non-zero (but possibly very small) effect size. Because sparse predictors perform about as well as predictors constructed using non-sparse methods [98–100], it is reasonable to conclude that actual genetic architectures are themselves sparse. For all known complex traits and disease risks, only a small fraction of common SNPs are required to build a predictor which performs nearly, or equally, as well as any non-sparse predictor.

In this paper we analyzed many aspects of sparse predictors. Within the class of algorithms that produce spare predictors we find that LASSO, also known as compressed sensing, is competitive with other more complex techniques. In fact, methodological improvements in predictor training, although important, generally produce improvements which are relatively small (in rough terms, of order 5 or 10 percent), whereas increases in data size are likely to produce much larger gains. Additionally, we demonstrate the performance characteristics promised from the compressed sensing literature. Specifically, we demonstrate phase transition behavior in performance using actual genotype/phenotype pairs – above a certain data threshold we recover the SNPs which provide the strongest contribution to performance metrics.

We develop a methodology for projecting the performance of LASSO on larger datasets. This method can be applied to anticipated future biobanks and to analysis which is forthcoming on existing biobanks. Specifically, we project that a predictor trained in the Taiwan Precision Medicine Initiative for asthma can achieve an AUC of 0.63_(0.02)_ and for height a correlation of 0.648_(0.009)_ for a Taiwanese population. For comparison, the measured values are 0.61_(0.01)_ and 0.631_(0.008)_, respectively, for UK Biobank trained predictors applied to a European population. We also show that, in terms of AUC, atrial fibrillation, type 2 diabetes, CAD, and breast cancer will more than double their signal if trained on larger datasets.

One of the main challenges of polygenic prediction is that most of the available data comes from studies in which most of the participants are individuals of European ancestry. Thus the current predictors perform poorly when applied to other ancestry groups (e.g., East Asians or Africans). The hope of developing methods which “transport” a predictor trained on one ancestry to other ancestral groups remains, but despite ongoing research efforts the goal remains elusive. In the absence of such a breakthrough, much larger cohorts of non-Europeans are required to produce predictors of comparable quality. The new methods developed here allow us to predict, e.g., how well new predictors trained on biobank-scale data from the Taiwan Precision Medicine Initiative (which is planned to surpass 1 million genotypes) and the US All of Us project. In the former case, we predict that new predictors will exceed AUC / correlation metrics for current best-in-class predictors for European ancestry.

## Data Availability

All data produced in the present study are either, contained in the manuscript, available at https://github.com/MSU-Hsu-Lab/biobank-scale-methods-paper-2023/, or available upon reasonable request to the authors.

## Acknowledgements

Computational resources provided by the Michigan State University High-Performance Computing Center. The authors acknowledge acquisition of data sets via UK Biobank Main Application 15326.

## Data Use and Institutional Review

Access to the UK Biobank resource is available via application (http://www.ukbiobank.ac.uk). UKB data was collected under policies conforming with national and local requirements and subject to privacy rights. (https://www.ukbiobank.ac.uk/privacy-policy) All data handled by researchers in this work was de-identified. MSU researchers working on this project do not have access to any identifiable data and are covered under MSU IRB [STUDY00006493].

## Public Data

Raw data is not available for direct sharing, but can be obtained via application to the UK biobank (https://www.ukbiobank.ac.uk/enable-your-research/apply-for-access). Predictors (i.e. SNPs and weights) and some code used to produce results can be found at https://github.com/MSU-Hsu-Lab/biobank-scale-methods.

## A Data

### A.1 Populations

The UK Biobank provides a self-reported ethnic background field (field 21000) on which to filter into various superpopulations – European, American, South Asian, East Asian and African. Self-reported Europeans are identified by the codes 1001, 1002, 1003 or 1 (White, British, Irish, Any other white background). Self-reported South Asians are identified by the codes 3, 3001, 3002, 3003, 3004 (Asian or Asian British, Indian, Pakistani, Bangladeshi, Any other Asian background). Self-reported East Asians are identified by the code 5 (Chinese). Self-reported Africans are identified by the codes 4, 4001, 4002, 4003, 4004 (Black or Black British, Caribbean, African, Any other Black background). Additionally code 6 is “Other ethnic group” and codes 2001-2004 are mixed background.

In addition to self-report, genetic ancestry is computed with ADMIXTURE Version 1.3.0 [101] and the 1000 genomes phase 3 as the reference panel. First, 1000 genomes was filtered down to SNPs which overlap the UK Biobank and then further sampled to 23,326 SNPs for computational ease. With this SNP subset, Admixture was run unsupervised on the 1000 genomes with 5 populations. The output P files (the allele frequencies of the inferred ancestral populations) were then applied via projection mode to the UK Biobank in batches - resulting in each individual with a percentage in each of the 5 super-populations (given by the ancestry fractions Q file).

The 5 different components were verified to correspond to the different super-populations in 1000 genomes. A set of American ancestry individuals was selected by keeping all individuals who had at least 35% on the American component of the analysis. This results in 322 individuals who correspond to the following report codes: 6:206, 1003:56, 2004:33, 1001:3, 4001/2003/2002/2001/2/1:1. These 322 individuals are considered the American super-population test group and are withheld from the other self-reported test sets.

### A.2 Siblings

The UK Biobank estimated kinship coefficients for individuals using *KING* as described in [102]. The UKB records related pairs of degree 3 or closer and provide the results as a single pairwise kinship table which is provided with the UK Biobank data. The set of siblings used in this work is identified by filtering this pairwise list on kinship coefficient and IBS0 in a manner similar done by the UKB (see the supplement of [102]). Specifically, to be included as a sibling pair, we keep all pairs with kinship coefficient larger than 0.176 and IBS0 larger than 0.0012. This procedure results in 22,667 sibling pairs, in agreement with [102].

### A.3 Phenotypes

The phenotype definitions used here involve various UKB-fields to define case and control status. The definitions are inclusive in that if any of the indicated data fields report a diagnoses or self report then the individual is counted as a case. For continuous phenotypes, the average of all measurements was used. The definitions used the ICD9, ICD10 and OPCS4 codes in UKB-fields 41271, 41270, 41272; self-reported non-cancer codes from UKB-field 20002 and cancer codes in UKB-field 20001. Additionally, some diseases were specifically included in the intake questionnaire or otherwise used other UKB-fields, which also are listed below.

There might be some quantitative performance gains that could come from the inclusion of more fields/codes for the phenotypes. Performance gains might also come from an analysis of related information like medication, lifestyle choices, and family history.

Training and evaluation of the predictors used a UKB download date of April 2021. The following disease definitions were used:

**Asthma** non-cancer codes: 1111; ICD9: 49300, 49309, 49310, 49319, 49390, 49399; ICD10: J450,J451,J458

**Atrial fibrillation** non-cancer codes: 1471, 1483; ICD9: 4273; ICD10: I480-I484, I489

**Breast cancer** cancer codes: 1002; ICD9: 174, 1749; ICD10: C50, C500-C506, C508, C509; field ID 40001: C50, C500-C506, C508, C509; field ID40002: C50, C500-C506, C508, C509; field ID 40006: C50, C500-C506, C508, C509; field ID 40013: 174, 1749

**Coronary artery disease** non-cancer codes: 1075; ICD9: 410, 4109, 412, 4129; ICD10: I21, I210-I214, I219, I21X, I22, I220, I221, I228, I229, I23, I230-I236, I238, I241, I252; OPCS4: K401-K404, K411-K414, K451-K455, K491, K492, K498, K499, K502, K751-K754, K758, K759

**Diabetes type I** non-cancer codes: 1222; ICD10: E100-E109, 0240

**Diabetes type II** non-cancer codes: 1223; ICD9: 25000, 25002, 25010, 25012, 25020, 25022, 25030, 25032, 25040, 25042, 25050, 25052, 25060, 25062, 25070, 25072, 25080, 25082, 25090, 25092; ICD10: E11,E110-E119

**Hypertension** non-cancer codes: 1065,1072,1073; ICD9: 4010,4011,4019,4050,4051,4059,4160,6420,6423,6429; ICD10: I10

Body Mass Index field ID: 21001

Direct Bilirubin field ID: 30660

**Height** field ID: 50

**Lipoprotein A** field ID: 30790

### A.4 Disease Prevalence

For asthma, recent surveys have found a prevalence of 10.9% in African Americans [103] (consistent with statistics from a decade earlier [104]). Recent studies have found the prevalence of asthma greatly increasing in Taiwan over the past several decades [105] (with rates in children reaching as high as ~ 20%), however we use the more conservatively reported number of 7.9% reported in [106]. The prevalence of asthma in Hispanic communities varies enormously up to ∼ 30% [107], but we use the conservative figure of 4.9% for Mexicans given in [107].

For atrial fibrillation a prevalence of 0.35% has been found for African Americans [108]. In Taiwan, a sex specific prevalence of 14% for men and 7% for women was found [109]. Because TPMI recruitment is still ongoing, this was averaged to 10.5%. For Hispanics and Latinos in the USA, after doing a weighted average over prevalences in sub-ancestry groups, an overall prevalence of just 1% was found[110].

Breast cancer prevalence in Hispanics fluctuates slightly within ancestry subgroups in the USA, but was recently observed to be 1% [111]. For African Americans the prevalence is much higher at 11.5% [112]. In Taiwan the prevalence is only .83% [113].

CAD as defined in A.3 is a complex phenotype made up of various self report, ICD, and OPCS codes. While this definition is widely used, e.g. in [29], it is not necessarily consistent with other definitions of CAD reported in survey literature. Because of this we use the reported prevalence in UKB, 5%, as the same prevalence in other biobanks. This is consistent with reports of CAD prevalence in Taiwan (4% sex averaged) [114], African Americans (5.4%) [115], and Hispanics in the USA (5.1%) [115].

It has been shown that there are differences in the rate and outcomes of hypertension in African Americans and caucasians in the USA [116]. Here it was found that the sex averaged rate of hypertension in African Americans is 42%. In Taiwan, the prevalence of hypertension is 26.1% according to a 2017 national survey [117]. Among Hispanics in the USA, the prevalence varies based on ancestry, but affects roughly 30% of the population [118].

The prevalence of type 1 diabetes has been found to be 0.57% among African Americans [119], 0.18% in American Hispanics [120], and as little as 0.01-0.05% in Taiwan [121, 122]. In contrast, type 2 diabetes is much more prevalent, being counted in roughly 13% of African Americans [123], 9.5% of American Hispanics [123], and 8.3% of Taiwanese [124].

For continuous traits—BMI, Direct Bilirubin, height, and Lipoprotein A—we assumed the same reporting rate as the UKB. For all 4 of these traits, measurements were recorded for 97% of UKB participants. This same rate was assumed for the other biobanks.

## B Uncertainty Analysis

Performance metrics—AUC, correlation, etc.—are often reported with a single measure of uncertainty, e.g. a standard error. In this work, the uncertainty of the metric estimate is a *key* ingredient to model predictive behavior. To this end, in this section we detail exactly how we characterize uncertainty.

### B.1 Standard Errors

Standard errors (SE) characterize the precision with which a statistic has been measured. SEs generally tend toward 0 as the amount of data increases. The exact form of the SEs depend on how the underlying statistic is modeled. In this work we use the following common definitions *uniformly minimum variance unbiased* (UMVU) estimators for a distribution with mean (*µ*), variance (*V*), and standard deviation (*σ*):

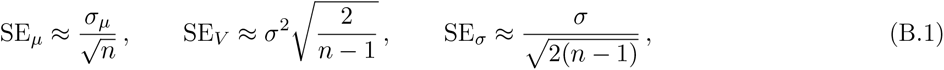

where *n* is the number of samples used to estimate the statistic. Similarly, for computing the correlation (*ρ*) between two sets of length *n* data, we can estimate the SE as:

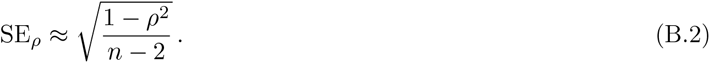

These SEs are all defined up to order O(1*/n*).

For computing AUCs we are not aware of a canonical *analytic* approximation of the SE. However, we can numerically approximate the uncertainty via Monte Carlo, i.e. we assume cases and controls are Gaussian distributed and randomly sample. As can be seen in 11 this was done for 20 different values of AUCs and 3 different ratios of cases to controls.

The AUC uncertainty can be fit with a simple polynomial. We find very little dependence on the number of controls, but instead find, after averaging over all 60 draws,

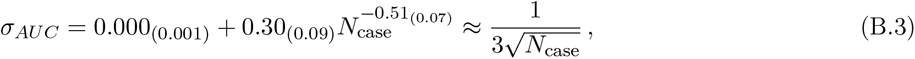

where the right hand side is the approximation we use in the analysis.

**Figure 11:**
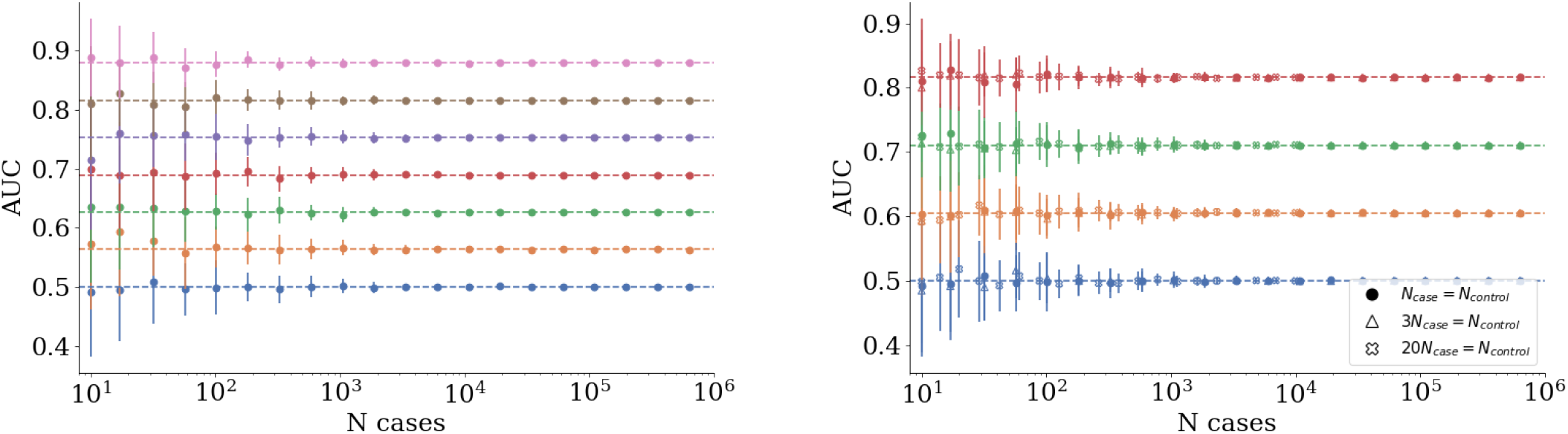
MC AUC error using simulated data. Left, equal cases and controls at different theoretical AUC values. Right, different ratios of cases of controls shows a very weak dependence on the number of controls.

### B.2 Data Uncertainty

Because understanding the uncertainty on our datapoints is critical to the analysis, especially the projection method, we give some examples here of how error bars/uncertainty was computed.

**Odds ratio** : the results presented for the LASSO algorithm involve 5-fold cross validation (CV). For each fold, an inclusive odds ratio (OR) is computed. At a given PGS value we can count how many cases have that PGS value or greater, *n*, how many controls have that PGS value or greater, *N*, and the total number of cases, *n*_0_, and total number of controls, *N*_0_. A standard error of the OR can be computed by looking at the log(OR) (for noisy data, this approach can lead to nonphysical OR bounds as the OR is not a symmetric distribution) and we find

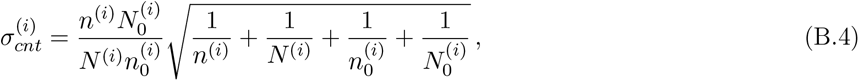

where we have added the superscript 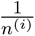 to indicate this contribution comes from the i-th CV fold. We use the label “*cnt*” to indicate that this quantity depends on the counts of cases and controls themselves. To compute the OR we take the mean over the folds, *µ*. This average itself has a standard deviation, *σ_µ_*. The total uncertainty associated with *µ* then has a contribution directly from the distribution of CV values and from the counts themselves,

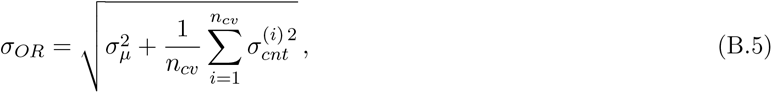

where *n_cv_* is the number of CV folds. The first term comes from the distribution of OR values over CV folds and the second term is an average uncertainty from the counts of cases and controls

**AUC** : computing the uncertainty of an AUC measurement is similar to that for the OR: there is one piece coming from averaging over the CV folds and a second piece that comes from the size of the data sample used. Using eq. (B.3), and the fact that each validation set is the same size, we have

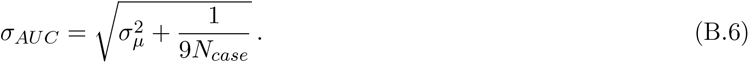

**Correlation** : the uncertainty is analogous to that computed for AUC but now using eq. (B.2),

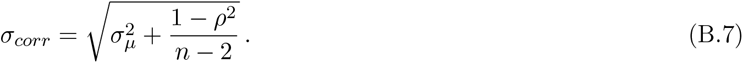

In all these examples, the second contribution is always dependent on the sample size used to compute the quantity. In practice, the size of this contribution compared to the size of the first contribution can be used as an indicator of whether enough data was used for validation/testing. In the results shown in this work the second term is consistently the smaller contribution indicating that the distribution over folds is the limiting factor.

## C Projection functions

The value of any metric will depend on the number of samples (cases and controls, or total number for continuous phenotype) used in training *and* validation/testing. It is not *a priori* known what functional form various metrics should take as a function of training samples. From a biological perspective it is true whatever function used should be bounded from above and below, i.e., there should be a lower limit to reflect that linear SNP genetics is playing no role and an upper limit to reflect that you have completely captured linear genetic effects.

Here we collect a group of bounded functions that are used in the main analysis

**Table 3:**
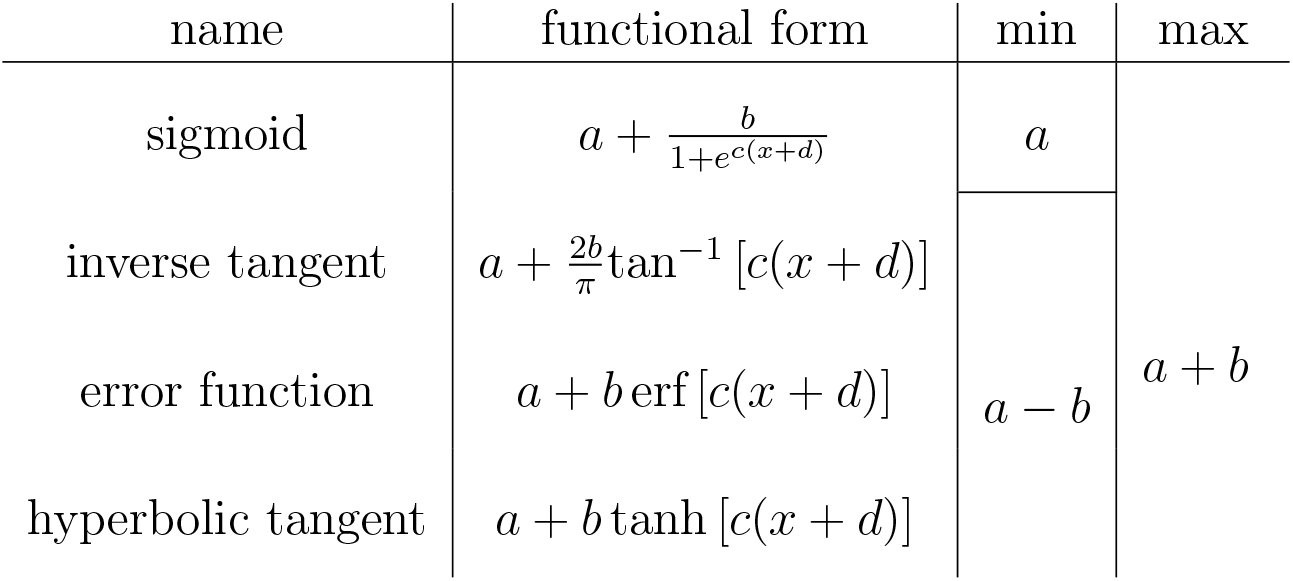
Various bounded functions used for fitting. Here, *x* is the log_10_ of the number of cases included in training. Note that all functions are centered at *x* = −*d* which implies that for *x <* −*d* there is faster than linear growth in AUC.

**Table 4:** Asymptotic central values, and uncertainty, from fitted curves for asthma.

| func | Asymp Mean | Asymp Stand Dev | SEM | 99% CI of Mean |
| --- | --- | --- | --- | --- |
| sigmoid4 | 0.7066 | 0.0510 | 0.0007 | 0.0019 |
| arctan4 | 0.7024 | 0.0347 | 0.0005 | 0.0013 |
| erf4 | 0.7260 | 0.0680 | 0.0010 | 0.0030 |
| tanhyp4 | 0.7062 | 0.0495 | 0.0007 | 0.0018 |

**Table 5:** Fit results for asthma. Note, for a fit *p_value_* → 1 indicates a poor fit and *p_value_* → 0 indicates possible over-fitting.

| func | $\chi^2/\text{dof}$ | p-value |
| --- | --- | --- |
| sigmoid4 | 0.72 | 0.65 |
| arctan4 | 0.95 | 0.46 |
| erf4 | 0.66 | 0.71 |
| tanhyp4 | 0.72 | 0.65 |

**Table 6:** Various functional predictions, and uncertainties, for asthma.

| | sigmoid | $\sigma$ | arctan | $\sigma$ | erf | $\sigma$ | tanh | $\sigma$ | pred | $\sigma$ |
| --- | --- | --- | --- | --- | --- | --- | --- | --- | --- | --- |
| SIB in UKB | 0.61 | 0.01 | 0.61 | 0.01 | 0.61 | 0.02 | 0.61 | 0.02 | 0.612 | 0.009 |
| AFR in AoU | 0.54 | 0.02 | 0.54 | 0.01 | 0.55 | 0.02 | 0.54 | 0.02 | 0.540 | 0.010 |
| EAS in TPMI | 0.63 | 0.02 | 0.63 | 0.02 | 0.63 | 0.03 | 0.63 | 0.02 | 0.630 | 0.020 |
| AMR in AoU | 0.52 | 0.01 | 0.52 | 0.01 | 0.52 | 0.02 | 0.52 | 0.01 | 0.520 | 0.010 |

**Table 7:** Asymptotic central values, and uncertainty, from fitted curves for atrial fibrillation

| func | Asymp Mean | Asymp Stand Dev | SEM | 99% CI of Mean |
| --- | --- | --- | --- | --- |
| sigmoid4 | 0.832 | 0.099 | 0.005 | 0.013 |
| arctan4 | 0.815 | 0.105 | 0.004 | 0.011 |
| erf4 | 0.821 | 0.104 | 0.004 | 0.011 |
| tanhyp4 | 0.820 | 0.102 | 0.004 | 0.011 |

**Table 8:** Fit results for atrial fibrillation. Note, for a fit p_value_→1 indicates a poor fit and p_value_→0 indicates possible over-fitting.

| func | $\chi^2/\text{dof}$ | p-value |
| --- | --- | --- |
| sigmoid4 | 0.71 | 0.66 |
| arctan4 | 0.59 | 0.76 |
| erf4 | 0.78 | 0.60 |
| tanhyp4 | 0.73 | 0.65 |

**Table 9:** Various functional predictions, and uncertainties, for atrial fibrillation.

| | sigmoid | $\sigma$ | arctan | $\sigma$ | erf | $\sigma$ | tanh | $\sigma$ | pred | $\sigma$ |
| --- | --- | --- | --- | --- | --- | --- | --- | --- | --- | --- |
| SIB in UKB | 0.62 | 0.05 | 0.62 | 0.07 | 0.61 | 0.03 | 0.61 | 0.04 | 0.61 | 0.03 |
| AFR in AoU | 0.51 | 0.02 | 0.52 | 0.03 | 0.51 | 0.01 | 0.51 | 0.02 | 0.51 | 0.01 |
| EAS in TPMI | 0.58 | 0.03 | 0.58 | 0.04 | 0.58 | 0.02 | 0.58 | 0.02 | 0.58 | 0.02 |
| AMR in AoU | 0.52 | 0.02 | 0.52 | 0.03 | 0.51 | 0.01 | 0.51 | 0.01 | 0.52 | 0.01 |

**Table 10:** Asymptotic central values, and uncertainty, from fitted curves for type 2 diabetes.

| func | Asymp Mean | Asymp Stand Dev | SEM | 99% CI of Mean |
| --- | --- | --- | --- | --- |
| sigmoid4 | 0.831 | 0.099 | 0.005 | 0.012 |
| arctan4 | 0.832 | 0.101 | 0.004 | 0.010 |
| erf4 | 0.828 | 0.095 | 0.003 | 0.009 |
| tanhyp4 | 0.823 | 0.102 | 0.004 | 0.009 |

**Table 11:** Fit results for type 2 diabetes. Note, for a fit *p_value_*→1 indicates a poor fit and *p_value_*→0 indicates possible over-fitting.

| func | $\chi^2/\text{dof}$ | p-value |
| --- | --- | --- |
| sigmoid4 | 2.16 | 0.03 |
| arctan4 | 2.03 | 0.05 |
| erf4 | 2.23 | 0.03 |
| tanhyp4 | 2.18 | 0.03 |

**Table 12:** Various functional predictions, and uncertainties, for type 2 diabetes.

| | sigmoid | $\sigma$ | arctan | $\sigma$ | erf | $\sigma$ | tanh | $\sigma$ | pred | $\sigma$ |
| --- | --- | --- | --- | --- | --- | --- | --- | --- | --- | --- |
| SIB in UKB | 0.62 | 0.04 | 0.62 | 0.05 | 0.62 | 0.03 | 0.62 | 0.04 | 0.62 | 0.02 |
| AFR in AoU | 0.57 | 0.02 | 0.57 | 0.03 | 0.57 | 0.02 | 0.57 | 0.02 | 0.57 | 0.01 |
| EAS in TPMI | 0.68 | 0.08 | 0.70 | 0.10 | 0.69 | 0.07 | 0.68 | 0.07 | 0.69 | 0.06 |
| AMR in AoU | 0.56 | 0.02 | 0.57 | 0.03 | 0.57 | 0.02 | 0.57 | 0.02 | 0.57 | 0.01 |

**Table 13:** Asymptotic central values, and uncertainty, from fitted curves for type 1 diabetes.

| func | Asymp Mean | Asymp Stand Dev | SEM | 99% CI of Mean |
| --- | --- | --- | --- | --- |
| sigmoid4 | 0.6640 | 0.0074 | 0.0001 | 0.0003 |
| arctan4 | 0.6755 | 0.0134 | 0.0002 | 0.0005 |
| erf4 | 0.6634 | 0.0073 | 0.0001 | 0.0003 |
| tanhyp4 | 0.6637 | 0.0076 | 0.0001 | 0.0003 |

**Table 14:** Fit results for type 1 diabetes. Note, for a fit p_value_→1 indicates a poor fit and p_value_→0 indicates possible over-fitting.

| func | $\chi^2/\text{dof}$ | p-value |
| --- | --- | --- |
| sigmoid4 | 0.18 | 0.99 |
| arctan4 | 0.12 | 1.00 |
| erf4 | 0.21 | 0.98 |
| tanhyp4 | 0.18 | 0.99 |

**Table 15:**
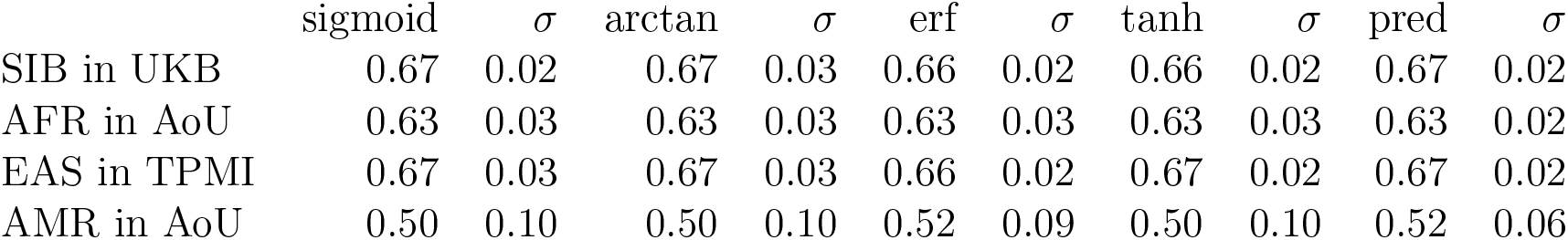
Various functional predictions, and uncertainties, for type 1 diabetes.

**Table 16:**
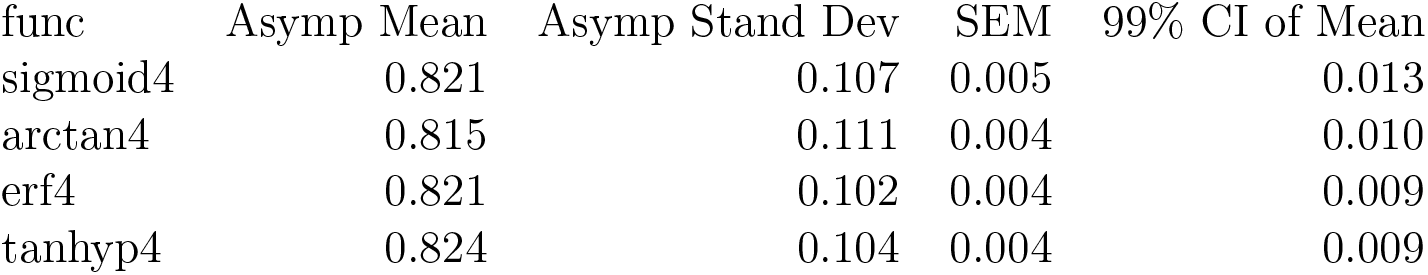
Asymptotic central values, and uncertainty, from fitted curves for CAD.

**Table 17:** Fit results for CAD. Note, for a fit *p_value_* → 1 indicates a poor fit and *p_value_* → 0 indicates possible over-fitting.

| func | $\chi^2/\text{dof}$ | p-value |
| --- | --- | --- |
| sigmoid4 | 1.65 | 0.12 |
| arctan4 | 1.70 | 0.10 |
| erf4 | 1.63 | 0.12 |
| tanhyp4 | 1.65 | 0.12 |

**Table 18:** Various functional predictions, and uncertainties, for CAD.

| | sigmoid | $\sigma$ | arctan | $\sigma$ | erf | $\sigma$ | tanh | $\sigma$ | pred | $\sigma$ |
| --- | --- | --- | --- | --- | --- | --- | --- | --- | --- | --- |
| SIB in UKB | 0.61 | 0.05 | 0.61 | 0.06 | 0.61 | 0.03 | 0.61 | 0.04 | 0.61 | 0.03 |
| AFR in AoU | 0.53 | 0.02 | 0.54 | 0.03 | 0.54 | 0.02 | 0.54 | 0.02 | 0.54 | 0.01 |
| EAS in TPMI | 0.68 | 0.09 | 0.70 | 0.10 | 0.65 | 0.06 | 0.67 | 0.08 | 0.67 | 0.05 |
| AMR in AoU | 0.53 | 0.02 | 0.54 | 0.03 | 0.53 | 0.02 | 0.53 | 0.02 | 0.53 | 0.01 |

**Table 19:** Asymptotic central values, and uncertainty, from fitted curves for hypertension.

| func | Asymp Mean | Asymp Stand Dev | SEM | 99% CI of Mean |
| --- | --- | --- | --- | --- |
| sigmoid4 | 0.6317 | 0.0093 | 0.0001 | 0.0003 |
| arctan4 | 0.6434 | 0.0084 | 0.0001 | 0.0003 |
| erf4 | 0.6299 | 0.0102 | 0.0001 | 0.0004 |
| tanhyp4 | 0.6321 | 0.0095 | 0.0001 | 0.0003 |

**Table 20:** Fit results for hypertension. Note, for a fit p_value_→1 indicates a poor fit and p_value_→0 indicates possible over-fitting.

| func | $\chi^2/\text{dof}$ | p-value |
| --- | --- | --- |
| sigmoid4 | 2.34 | 0.02 |
| arctan4 | 2.30 | 0.02 |
| erf4 | 2.47 | 0.02 |
| tanhyp4 | 2.34 | 0.02 |

**Table 21:** Various functional predictions, and uncertainties, for hypertension.

| | sigmoid | $\sigma$ | arctan | $\sigma$ | erf | $\sigma$ | tanh | $\sigma$ | pred | $\sigma$ |
| --- | --- | --- | --- | --- | --- | --- | --- | --- | --- | --- |
| SIB in UKB | 0.61 | 0.01 | 0.610 | 0.010 | 0.62 | 0.01 | 0.62 | 0.01 | 0.615 | 0.006 |
| AFR in AoU | 0.55 | 0.01 | 0.550 | 0.010 | 0.55 | 0.01 | 0.55 | 0.01 | 0.551 | 0.006 |
| EAS in TPMI | 0.55 | 0.01 | 0.540 | 0.010 | 0.55 | 0.01 | 0.55 | 0.01 | 0.546 | 0.006 |
| AMR in AoU | 0.53 | 0.01 | 0.528 | 0.009 | 0.53 | 0.01 | 0.53 | 0.01 | 0.528 | 0.005 |

**Table 22:**
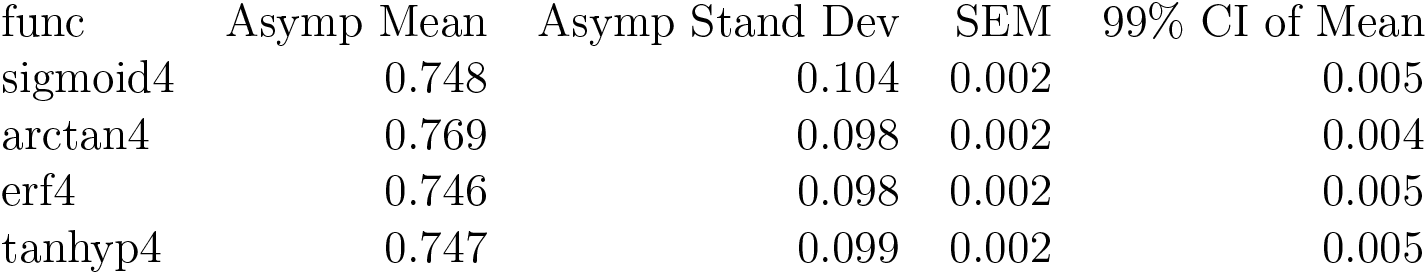
Asymptotic central values, and uncertainty, from fitted curves for breast cancer.

| func | Asymp Mean | Asymp Stand Dev | SEM | 99% CI of Mean |
| --- | --- | --- | --- | --- |
| sigmoid4 | 0.748 | 0.104 | 0.002 | 0.005 |
| arctan4 | 0.769 | 0.098 | 0.002 | 0.004 |
| erf4 | 0.746 | 0.098 | 0.002 | 0.005 |
| tanhyp4 | 0.747 | 0.099 | 0.002 | 0.005 |

**Table 23:** Fit results for breast cancer. Note, for a fit *p_value_*→1 indicates a poor fit and *p_value_*→0 indicates possible over-fitting.

| func | $\chi^2/\text{dof}$ | p-value |
| --- | --- | --- |
| sigmoid4 | 0.84 | 0.56 |
| arctan4 | 0.89 | 0.51 |
| erf4 | 0.82 | 0.57 |
| tanhyp4 | 0.84 | 0.56 |

**Table 24:** Various functional predictions, and uncertainties, for breast cancer.

| | sigmoid | $\sigma$ | arctan | $\sigma$ | erf | $\sigma$ | tanh | $\sigma$ | pred | $\sigma$ |
| --- | --- | --- | --- | --- | --- | --- | --- | --- | --- | --- |
| EUR/SIB in UKB | 0.58 | 0.03 | 0.58 | 0.02 | 0.58 | 0.02 | 0.58 | 0.03 | 0.58 | 0.01 |
| AFR in AoU | 0.57 | 0.03 | 0.57 | 0.02 | 0.57 | 0.02 | 0.57 | 0.03 | 0.57 | 0.01 |
| EAS in TPMI | 0.57 | 0.03 | 0.58 | 0.02 | 0.57 | 0.02 | 0.58 | 0.03 | 0.57 | 0.01 |
| AMR in AoU | 0.52 | 0.02 | 0.52 | 0.02 | 0.52 | 0.02 | 0.52 | 0.02 | 0.52 | 0.01 |

**Table 25:** Asymptotic central values, and uncertainty, from fitted curves for direct bilirubin.

| func | Asymp Mean | Asymp Stand Dev | SEM | 99% CI of Mean |
| --- | --- | --- | --- | --- |
| sigmoid4 | 0.5226 | 0.0274 | 0.0004 | 0.0010 |
| arctan4 | 0.5249 | 0.0240 | 0.0003 | 0.0009 |
| erf4 | 0.5279 | 0.0324 | 0.0005 | 0.0013 |
| tanhyp4 | 0.5218 | 0.0270 | 0.0004 | 0.0010 |

**Table 26:** Fit results for direct bilirubin. Note, for a fit p_value_→1 indicates a poor fit and p_value_→0 indicates possible over-fitting.

| func | $\chi^2/\text{dof}$ | p-value |
| --- | --- | --- |
| sigmoid4 | 0.64 | 0.72 |
| arctan4 | 0.63 | 0.73 |
| erf4 | 0.65 | 0.72 |
| tanhyp4 | 0.64 | 0.72 |

**Table 27:** Comparison of known correlation and heritability methods to the asymptotic prediction for direct bilirubin.

| | Corr | $\sigma$ | heritability | $\sigma$ |
| --- | --- | --- | --- | --- |
| GCTA | 0.660 | 0.080 | 0.40 | 0.40 |
| LDSR | 0.711 | 0.006 | 0.51 | 0.02 |
| Asymp | 0.520 | 0.010 | 0.27 | 0.01 |

**Table 28:** Various functional predictions, and uncertainties, for direct bilirubin.

| | sigmoid | $\sigma$ | arctan | $\sigma$ | erf | $\sigma$ | tanh | $\sigma$ | pred | $\sigma$ |
| --- | --- | --- | --- | --- | --- | --- | --- | --- | --- | --- |
| SIB in UKB | 0.50 | 0.04 | 0.49 | 0.03 | 0.52 | 0.06 | 0.50 | 0.04 | 0.50 | 0.02 |
| AFR in AoU | 0.51 | 0.05 | 0.50 | 0.03 | 0.53 | 0.07 | 0.51 | 0.05 | 0.51 | 0.03 |
| EAS in TPMI | 0.50 | 0.04 | 0.49 | 0.03 | 0.51 | 0.05 | 0.50 | 0.04 | 0.50 | 0.02 |
| AMR in AoU | 0.51 | 0.05 | 0.50 | 0.03 | 0.53 | 0.07 | 0.51 | 0.05 | 0.51 | 0.03 |

**Table 29:**
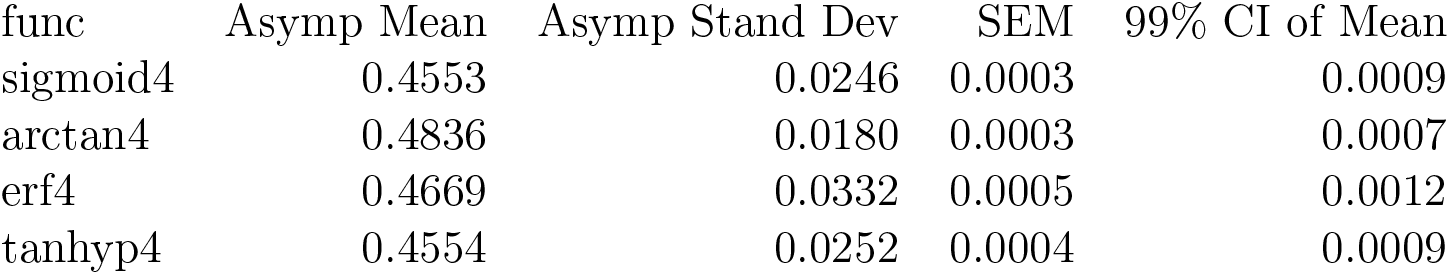
Asymptotic central values, and uncertainty, from fitted curves for BMI.

**Table 30:** Fit results for BMI. Note, for a fit p_value_ → 1 indicates a poor fit and p_value_ → 0 indicates possible over-fitting.

| func | $\chi^2/\text{dof}$ | p-value |
| --- | --- | --- |
| sigmoid4 | 0.20 | 0.98 |
| arctan4 | 1.11 | 0.35 |
| erf4 | 0.18 | 0.99 |
| tanhyp4 | 0.20 | 0.98 |

**Table 31:**
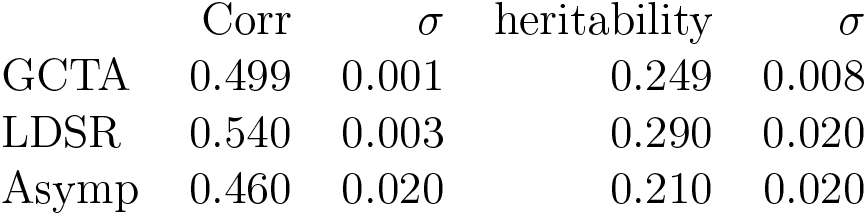
Comparison of known correlation and heritability methods to the asymptotic prediction for BMI.

| | Corr | $\sigma$ | heritability | $\sigma$ |
| --- | --- | --- | --- | --- |
| GCTA | 0.499 | 0.001 | 0.249 | 0.008 |
| LDSR | 0.540 | 0.003 | 0.290 | 0.020 |
| Asymp | 0.460 | 0.020 | 0.210 | 0.020 |

**Table 32:** Various functional predictions, and uncertainties, for BMI.

| | sigmoid | $\sigma$ | arctan | $\sigma$ | erf | $\sigma$ | tanh | $\sigma$ | pred | $\sigma$ |
| --- | --- | --- | --- | --- | --- | --- | --- | --- | --- | --- |
| SIB in UKB | 0.36 | 0.01 | 0.36 | 0.01 | 0.36 | 0.01 | 0.36 | 0.01 | 0.36 | 0.01 |
| AFR in AoU | 0.16 | 0.01 | 0.16 | 0.02 | 0.17 | 0.01 | 0.16 | 0.01 | 0.16 | 0.01 |
| EAS in TPMI | 0.40 | 0.03 | 0.39 | 0.02 | 0.40 | 0.03 | 0.40 | 0.03 | 0.40 | 0.02 |
| AMR in AoU | 0.14 | 0.01 | 0.13 | 0.01 | 0.14 | 0.01 | 0.14 | 0.01 | 0.14 | 0.01 |

**Table 33:** Asymptotic central values, and uncertainty, from fitted curves for height.

| func | Asymp Mean | Asymp Stand Dev | SEM | 99% CI of Mean |
| --- | --- | --- | --- | --- |
| sigmoid4 | 0.66702 | 0.00618 | 0.00009 | 0.00022 |
| arctan4 | 0.75880 | 0.00990 | 0.00010 | 0.00040 |
| erf4 | 0.65413 | 0.00550 | 0.00008 | 0.00020 |
| tanhyp4 | 0.66715 | 0.00609 | 0.00009 | 0.00022 |

**Table 34:** Fit results for height. Note, for a fit p_value_ → 1 indicates a poor fit and p_value_ → 0 indicates possible over-fitting.

| func | $\chi^2/\text{dof}$ | p-value |
| --- | --- | --- |
| sigmoid4 | 1.48 | 0.1700 |
| arctan4 | 4.43 | 0.0001 |
| erf4 | 0.99 | 0.4400 |
| tanhyp4 | 1.48 | 0.1700 |

**Table 35:** Comparison of known correlation and heritability methods to the asymptotic prediction for height.

| | Corr | $\sigma$ | heritability | $\sigma$ |
| --- | --- | --- | --- | --- |
| GCTA | 0.697 | 0.005 | 0.49 | 0.02 |
| LDSR | 0.717 | 0.006 | 0.51 | 0.02 |
| Asymp | 0.663 | 0.003 | 0.439 | 0.005 |

**Table 36:** Various functional predictions, and uncertainties, for height.

| | sigmoid | $\sigma$ | arctan | $\sigma$ | erf | $\sigma$ | tanh | $\sigma$ | pred | $\sigma$ |
| --- | --- | --- | --- | --- | --- | --- | --- | --- | --- | --- |
| SIB in UKB | 0.631 | 0.008 | 0.63 | 0.008 | 0.631 | 0.008 | 0.631 | 0.008 | 0.631 | 0.006 |
| AFR in AoU | 0.470 | 0.010 | 0.48 | 0.010 | 0.470 | 0.010 | 0.470 | 0.010 | 0.470 | 0.008 |
| EAS in TPMI | 0.650 | 0.010 | 0.65 | 0.010 | 0.650 | 0.010 | 0.650 | 0.010 | 0.648 | 0.009 |
| AMR in AoU | 0.430 | 0.010 | 0.44 | 0.010 | 0.430 | 0.010 | 0.430 | 0.010 | 0.434 | 0.009 |

**Table 37:** Asymptotic central values, and uncertainty, from fitted curves for lipoprotein A.

| func | Asymp Mean | Asymp Stand Dev | SEM | 99% CI of Mean |
| --- | --- | --- | --- | --- |
| sigmoid4 | 0.7721 | 0.0100 | 0.0001 | 0.0004 |
| arctan4 | 0.7992 | 0.0192 | 0.0003 | 0.0007 |
| erf4 | 0.7690 | 0.0082 | 0.0001 | 0.0003 |
| tanhyp4 | 0.7722 | 0.0101 | 0.0001 | 0.0004 |

**Table 38:** Fit results for lipoprotein A. Note, for a fit p_value_→1 indicates a poor fit and p_value_→0 indicates possible over-fitting.

| func | $\chi^2/\text{dof}$ | p-value |
| --- | --- | --- |
| sigmoid4 | 0.13 | 1.0 |
| arctan4 | 0.12 | 1.0 |
| erf4 | 0.14 | 1.0 |
| tanhyp4 | 0.13 | 1.0 |

**Table 39:** Comparison of known correlation and heritability methods to the asymptotic prediction for lipoprotein A.

| | Corr | $\sigma$ | heritability | $\sigma$ |
| --- | --- | --- | --- | --- |
| GCTA | 0.356 | 0.006 | 0.13 | 0.09 |
| LDSR | 0.670 | 0.005 | 0.45 | 0.02 |
| Asymp | 0.779 | 0.007 | 0.61 | 0.01 |

**Table 40:** Various functional predictions, and uncertainties, for lipoprotein A.

| | sigmoid | $\sigma$ | arctan | $\sigma$ | erf | $\sigma$ | tanh | $\sigma$ | pred | $\sigma$ |
| --- | --- | --- | --- | --- | --- | --- | --- | --- | --- | --- |
| SIB in UKB | 0.73 | 0.03 | 0.75 | 0.02 | 0.68 | 0.09 | 0.73 | 0.04 | 0.72 | 0.03 |
| AFR in AoU | 0.72 | 0.04 | 0.74 | 0.02 | 0.67 | 0.09 | 0.72 | 0.04 | 0.71 | 0.03 |
| EAS in TPMI | 0.74 | 0.03 | 0.75 | 0.02 | 0.68 | 0.09 | 0.74 | 0.04 | 0.73 | 0.03 |
| AMR in AoU | 0.72 | 0.04 | 0.73 | 0.02 | 0.67 | 0.09 | 0.72 | 0.04 | 0.71 | 0.03 |

**Figure 12:**
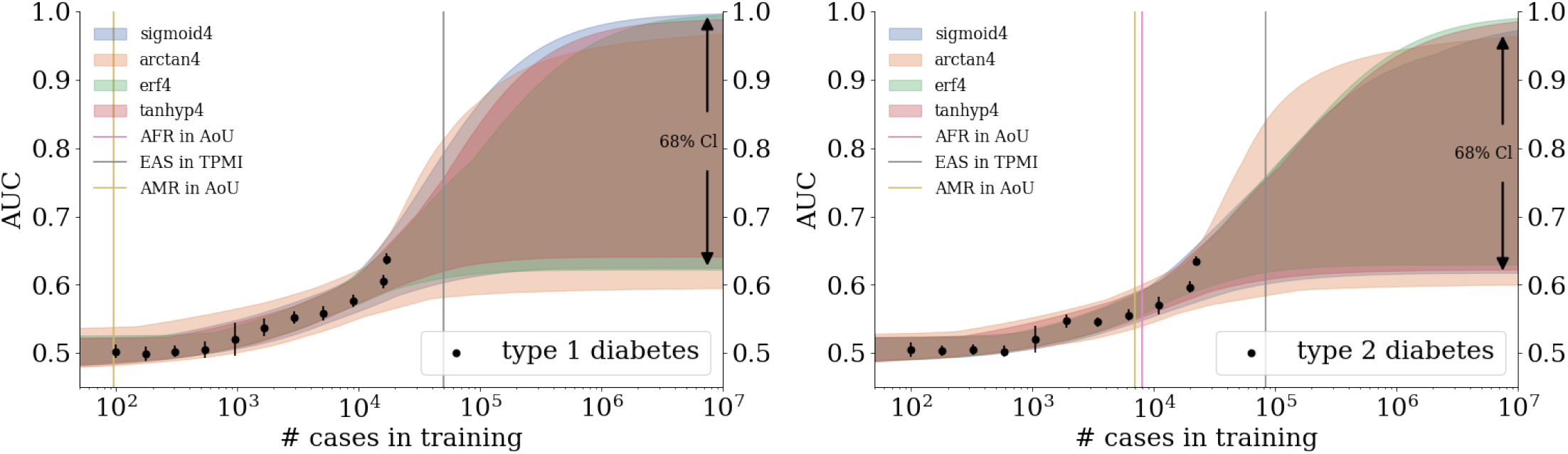
Growth of AUC as a function of training size in the UKB for diabetes type 1 and diabetes type 2. Colored, curved bands come from fitting data with various 4 parameter functions. Width of the bands corresponds to a ∼68%, or 2 standard deviations, confidence interval on the predictions. Vertical bars represent projections for de novo training in other biobanks using literature prevalences.

**Figure 13:**
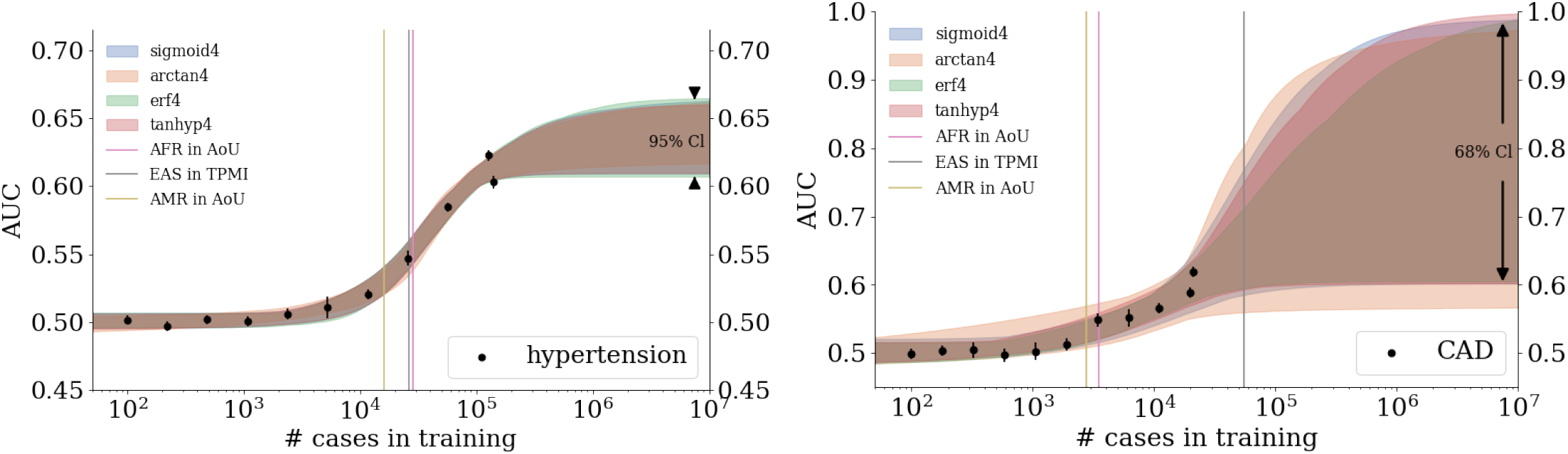
Growth of AUC as a function of training size in the UKB for hypertension and coronary artery disease. Colored, curved bands come from fitting data with various 4 parameter functions. Width of the band corresponds to a confidence interval on the predictions: on the right 2 standard deviations or ∼68% and on the left 4 standard deviations or ∼95%. Vertical bars represent projections for de novo training in other biobanks using literature prevalences.

**Figure 14:**
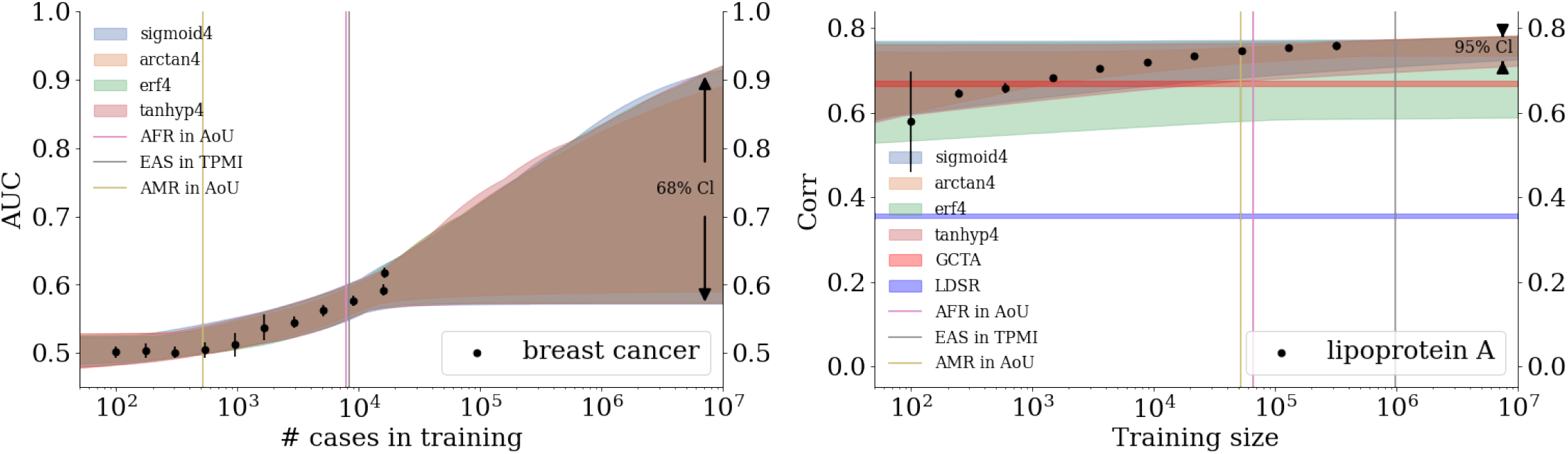
Growth of AUC (left: breast cancer) and correlation (right: lipoprotein A) as a function of training size in the UKB. Colored, curved bands come from fitting data with various 4 parameter functions. Width of the band corresponds to a confidence interval on the predictions: on the left 2 standard deviations or ∼68% and on the right 4 standard deviations or ∼95%. Vertical bars represent projections for de novo training in other biobanks using literature prevalences. On the right, horizontal lines indicate the correlation predicted from GCTA and LDSR.

**Figure 15:**
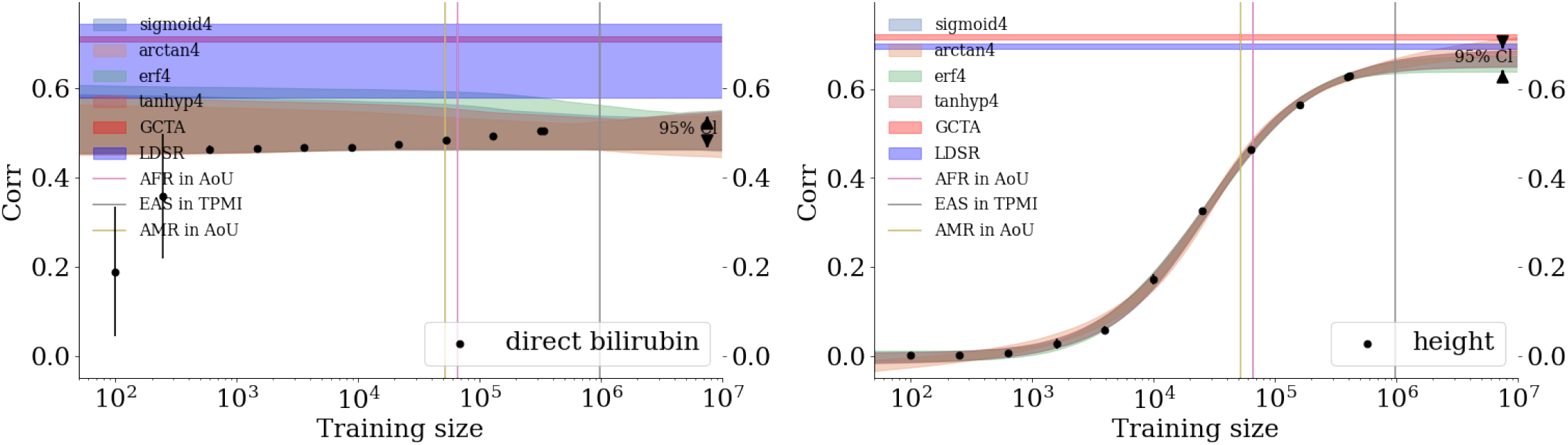
Growth of correlation as a function of training size in the UKB for direct bilirubin and height. Colored, curved bands come from fitting data with various 4 parameter functions. Width of the band corresponds to a ∼95% confidence interval, or 4 standard deviations. Vertical bars represent projections for de novo training in other biobanks using literature prevalences. Horizontal lines indicate the correlation predicted from GCTA and LDSR.

**Figure 16:**
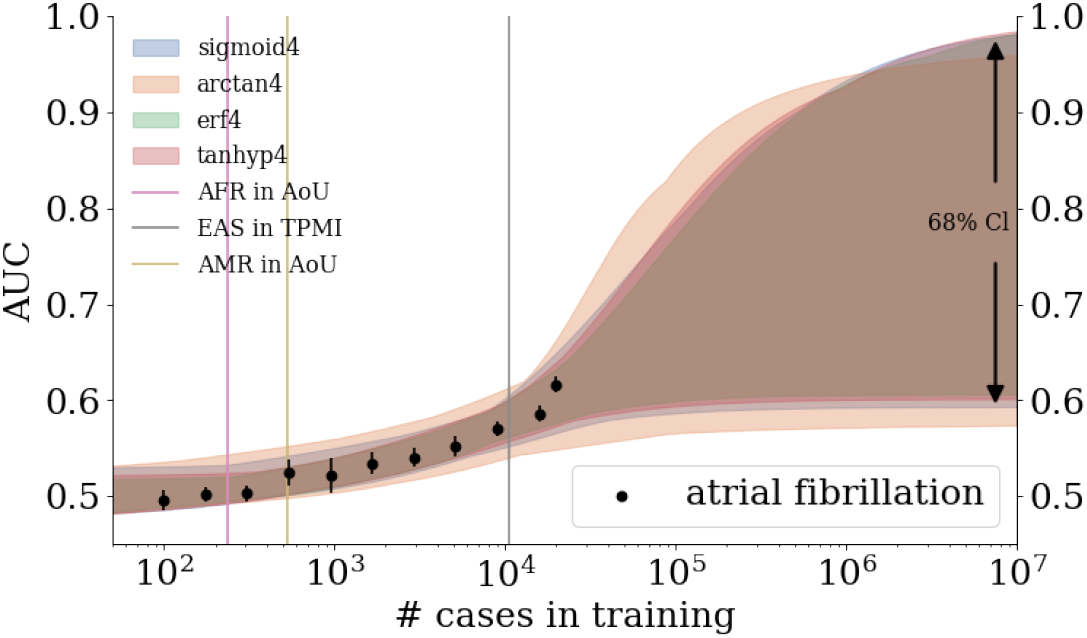
Growth of AUC as a function of training size in the UKB for atrial fibrillation. Colored, curved bands come from fitting data with various 4 parameter functions. Width of the band corresponds to a ∼68% confidence interval, or two standard deviations, on the predictions. Vertical bars represent projections for de novo training in other biobanks using literature prevalences.

## D Sparse methods comparisons on various phenotypes

**Figure 17:**
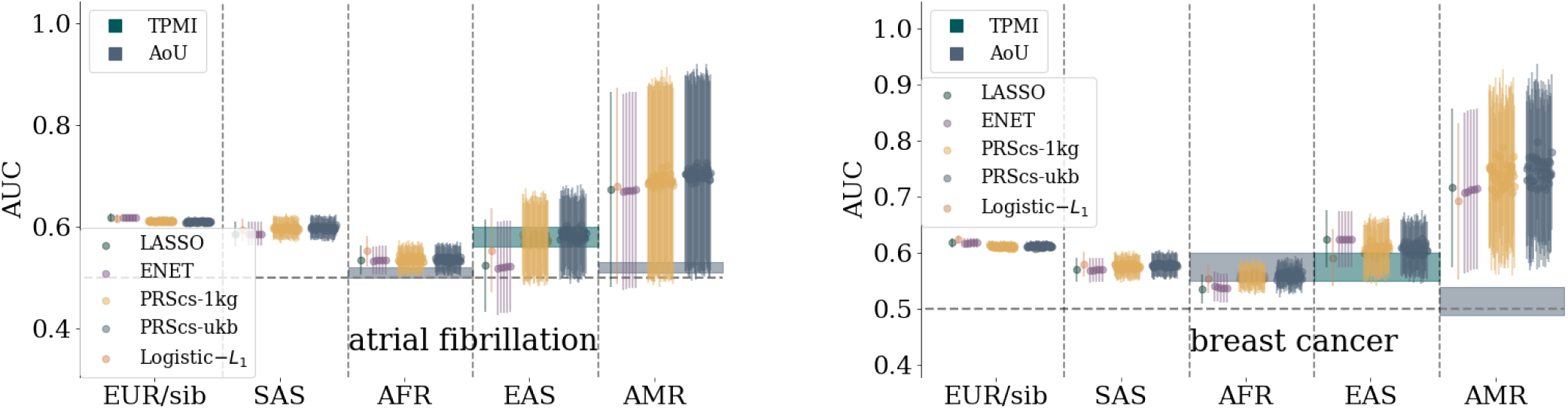
Comparison of sparse methods for atrial fibrillation and breast cancer predictors with a comparison to prediction bands for more diverse biobanks. Both trait predictors are trained on a UKB white population. Predictors are built with LASSO, *L*_1_-penalized Logistic regression, Elastic Net, and PRScs with UKB and 1,000 Genomes LD matrices. The specific parameters for the Elastic Net and PRScs are described in section 3.

**Figure 18:**
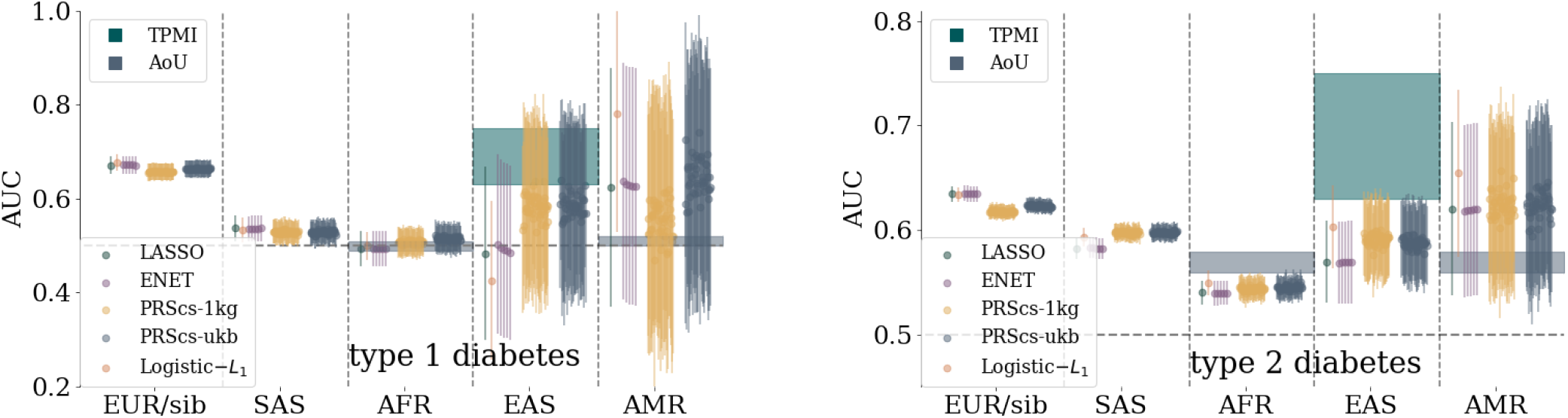
Comparison of sparse methods for type 1 diabetes and type 2 diabetes predictors with a comparison to prediction bands for more diverse biobanks. Both trait predictors are trained on a UKB white population. Predictors are built with LASSO, *L*_1_-penalized Logistic regression, Elastic Nets, and PRScs with UKB and 1,000 Genomes LD matrices. The specific parameters for the Elastic nets and PRScs are described in section 3.

**Figure 19:**
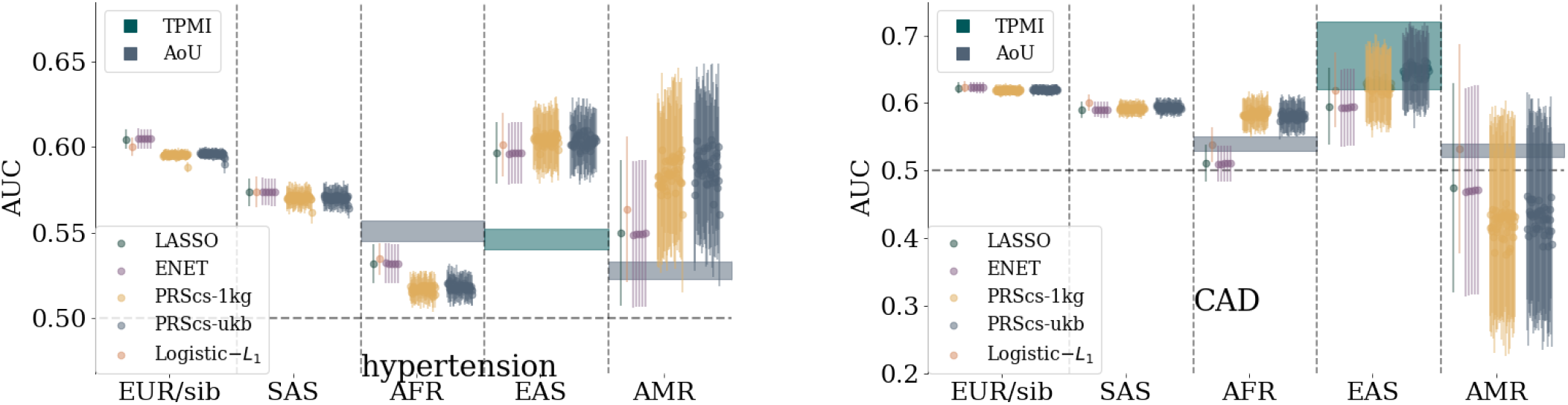
Comparison of sparse methods for hypertension and coronary artery disease predictors with a comparison to prediction bands for more diverse biobanks. Both trait predictors are trained on a UKB white population. Predictors are built with LASSO, *L*_1_-penalized Logistic regression, Elastic Nets, and PRScs with UKB and 1,000 Genomes LD matrices. The specific parameters for the Elastic nets and PRScs are described in section 3.

**Figure 20:**
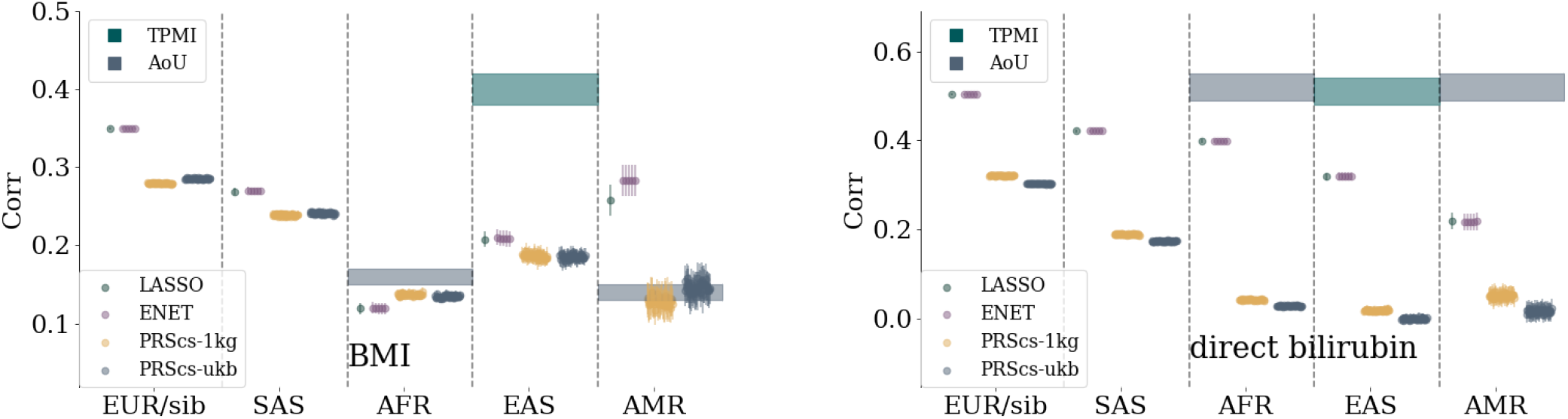
Comparison of sparse methods for BMI and direct bilirubin predictors with a comparison to prediction bands for more diverse biobanks. Both trait predictors are trained on a UKB white population. Predictors are built with LASSO, *L*_1_-penalized Logistic regression, Elastic Nets, and PRScs with UKB and 1,000 Genomes LD matrices. The specific parameters for the Elastic nets and PRScs are described in section 3.

**Figure 21:**
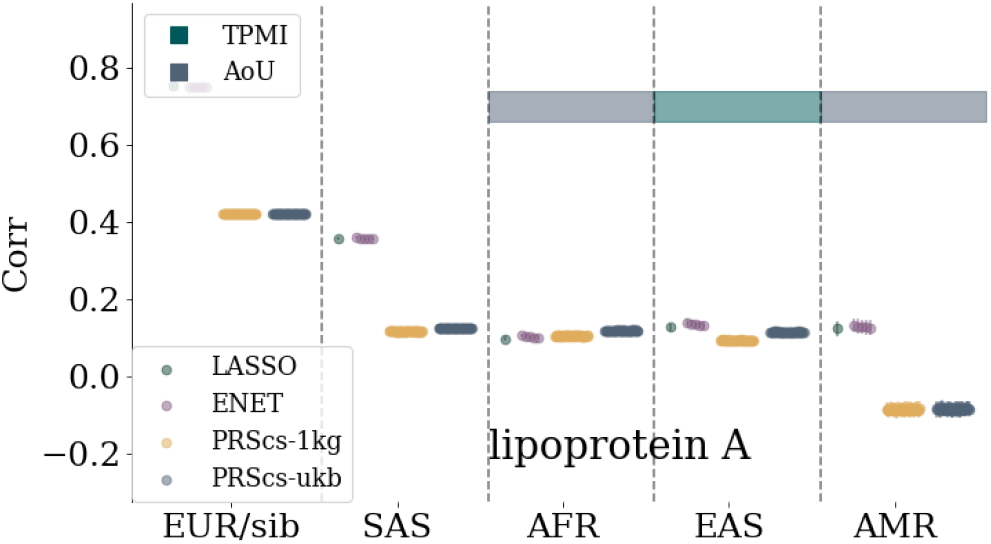
Comparison of sparse methods for lipoprotein A predictors with a comparison to prediction bands for more diverse biobanks. The trait predictors are trained on a UKB white population. Predictors are built with LASSO, *L*_1_-penalized Logistic regression, Elastic Nets, and PRScs with UKB and 1,000 Genomes LD matrices. The specific parameters for the Elastic nets and PRScs are described in section 3.

## E Sibling Tests

**Figure 22:**
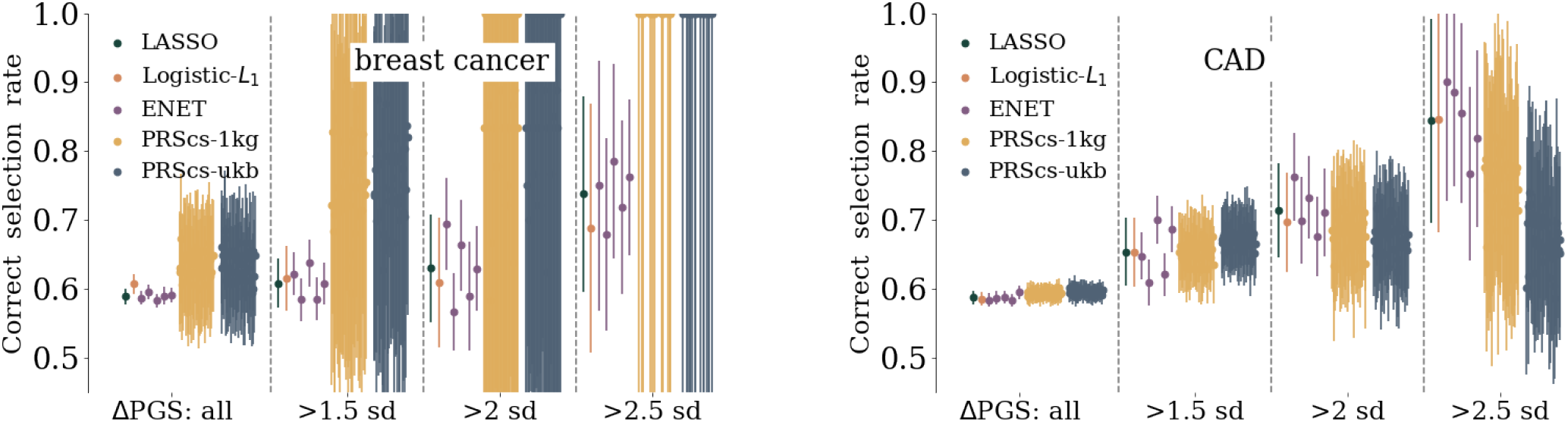
Affected sibling pair (ASP) selection rate for breast cancer and coronary artery disease. Pairs of siblings, where one person is a case and the other a control, are used and the rate corresponds to the number of times the case sibling has the higher PGS. The rate of correct selection, and uncertainty, increases if the siblings are also separated by at least 1.5, 2, or 2.5 standard deviations in PGS.

**Figure 23:**
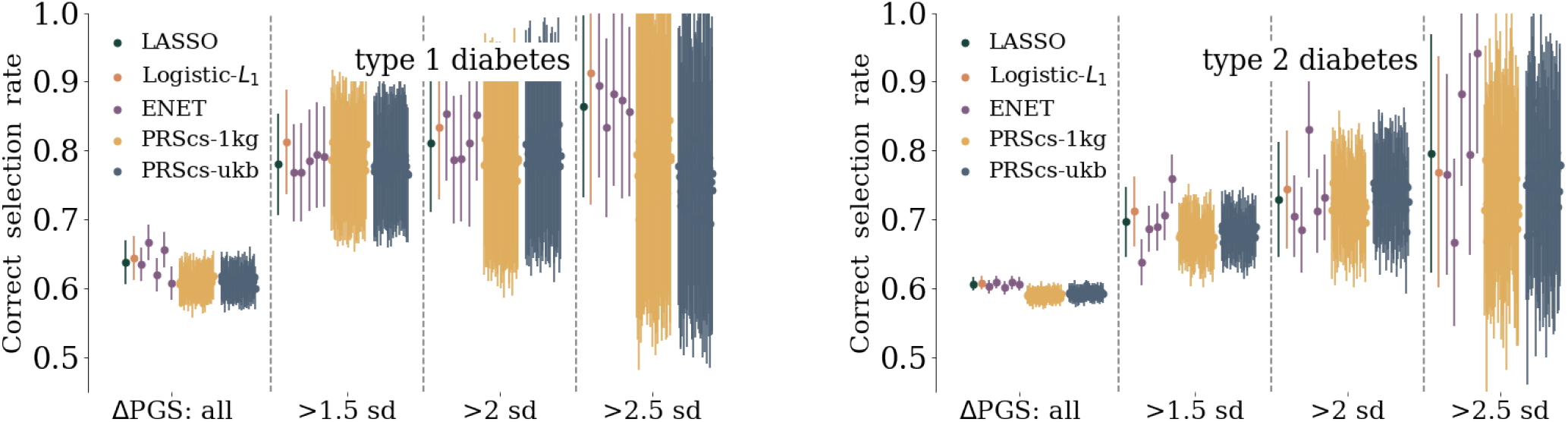
Affected sibling pair (ASP) selection rate for type 1 diabetes and type 2 diabetes. Pairs of siblings, where one person is a case and the other a control, are used and the rate corresponds to the number of times the case sibling has the higher PGS. The rate of correct selection, and uncertainty, increases if the siblings are also separated by at least 1.5, 2, or 2.5 standard deviations in PGS.

**Figure 24:**
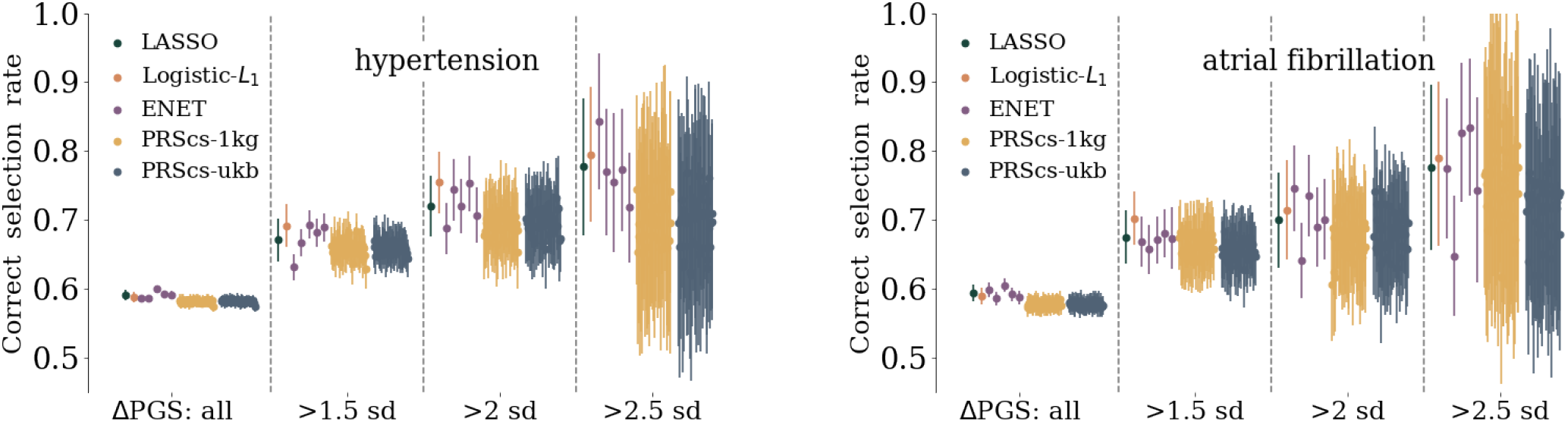
Affected sibling pair (ASP) selection rate for hypertension and atrial fibrillation. Pairs of siblings, where one person is a case and the other a control, are used and the rate corresponds to the number of times the case sibling has the higher PGS. The rate of correct selection, and uncertainty, increases if the siblings are also separated by at least 1.5, 2, or 2.5 standard deviations in PGS.

**Figure 25:**
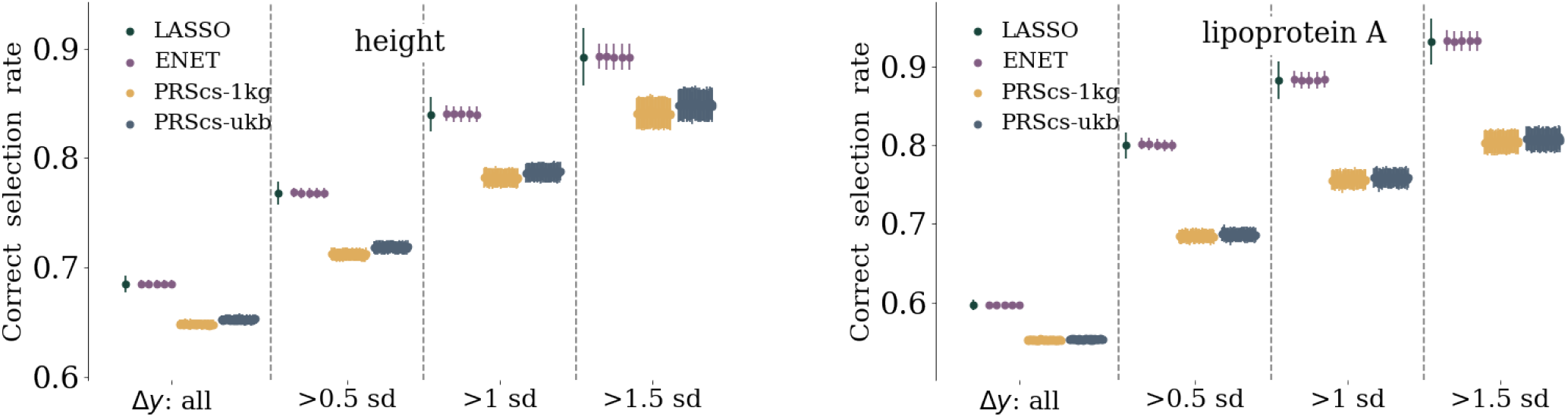
Rank order selection rate for height and lipoprotein A. The rate corresponds to frequency of the sibling with the larger phenotype also having the larger PGS. The selection rate, and uncertainty, increase if you require that the siblings phenotype is separated by at least 0.5, 1, or 1.5 standard deviations.

**Figure 26:**
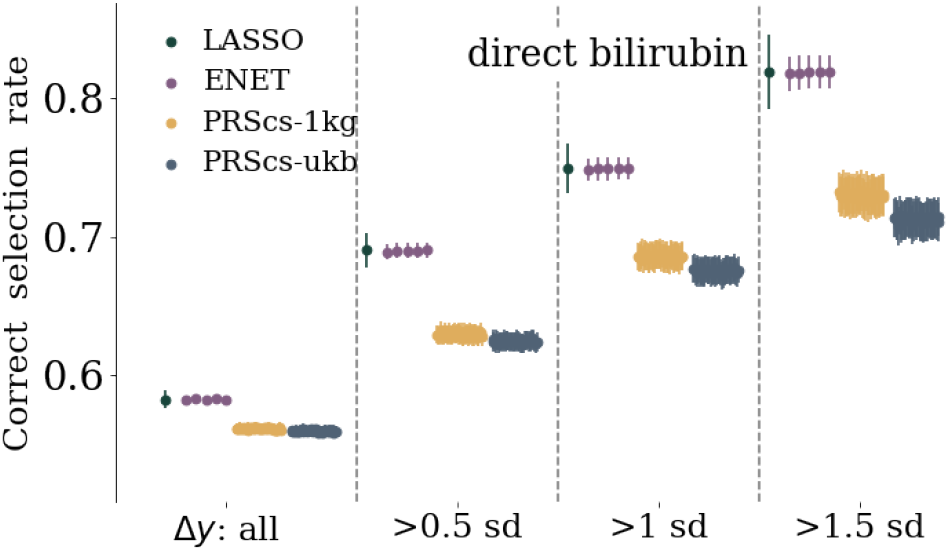
Rank order selection rate for direct bilirubin. The rate corresponds to frequency of the sibling with the larger phenotype also having the larger PGS. The selection rate, and uncertainty, increase if you require that the siblings phenotype is separated by at least 0.5, 1, or 1.5 standard deviations.

## F Impact Regions

Here we collect, in table form, the most impactful regions for LASSO predictors for each phenotype. The impactful SNPs are found by looking at SSV for each SNP. Associated genes are identified by using www.ncbi.nlm.nih.gov/gene/ gene database, for primary build GRCh37, and extending the gene region by 2 million base pairs in both directions to capture possibly associated SNPs.

**Table 41:**
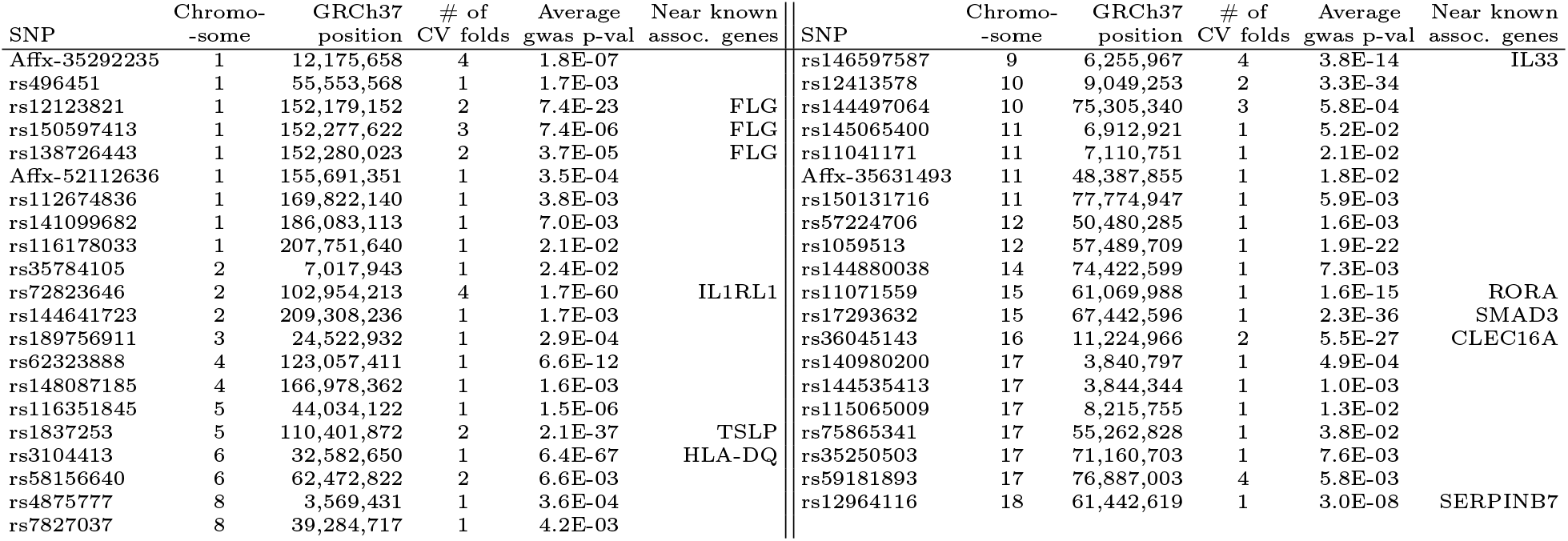
Asthma SNPs with SSV > 1% of total SSV.

**Table 42:**
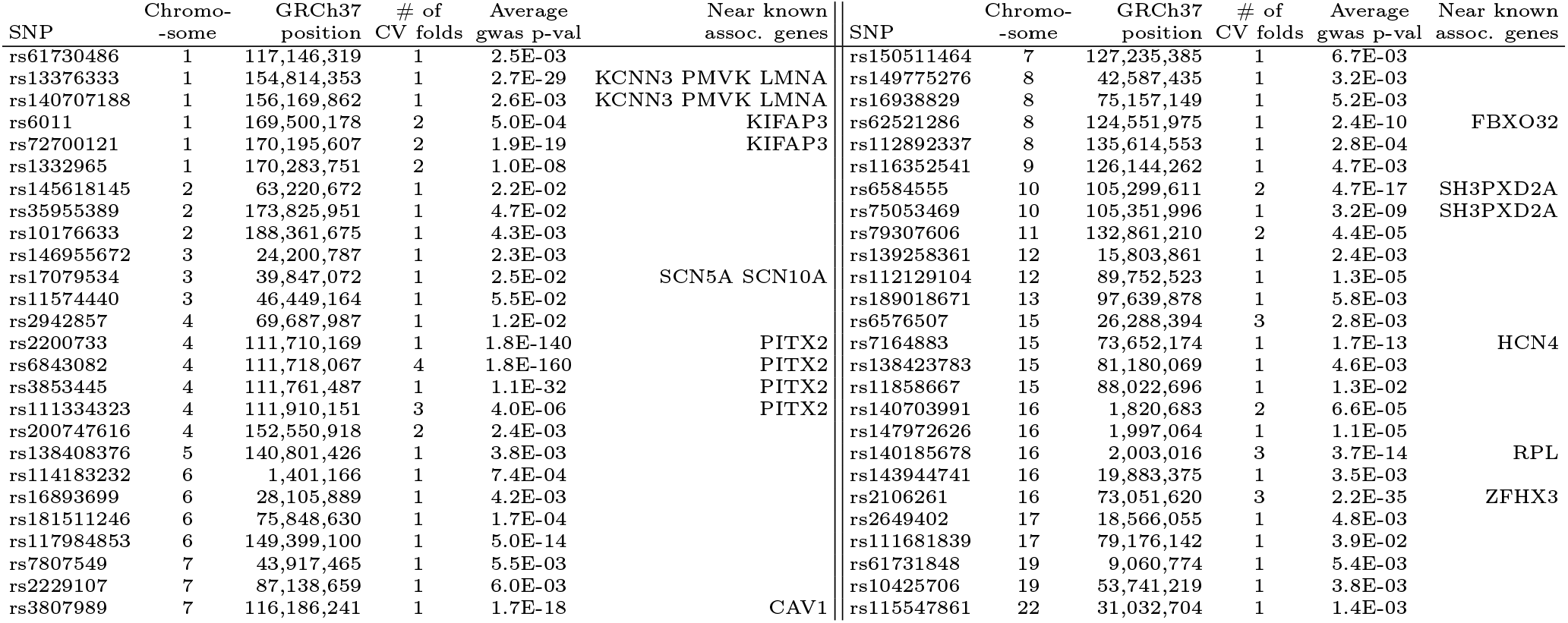
Atrial fibrillation SNPs with SSV > 1% of total SSV.

**Table 43:** Breast cancer SNPs with SSV > 1% of total SSV.

| SNP | Chromo-<br>-some | GRCh37<br>position | # of<br>CV folds | Average<br>gwas p-val | Near known<br>assoc. genes | SNP | Chromo-<br>-some | GRCh37<br>position | # of<br>CV folds | Average<br>gwas p-val | Near known<br>assoc. genes |
| --- | --- | --- | --- | --- | --- | --- | --- | --- | --- | --- | --- |
| rs77316894 | 1 | 90,804,699 | 1 | 1.4E-05 |  | rs3780548 | 9 | 99,525,487 | 2 | 8.6E-06 |  |
| rs138421943 | 1 | 152,192,110 | 3 | 9.8E-04 |  | rs74541872 | 9 | 116,798,596 | 4 | 5.4E-03 |  |
| rs150343459 | 2 | 54,021,461 | 4 | 1.1E-04 |  | rs1219648 | 10 | 123,346,190 | 1 | 7.5E-100 |  |
| rs72951831 | 2 | 217,957,699 | 2 | 1.4E-12 |  | rs2295878 | 10 | 123,996,934 | 1 | 8.2E-03 |  |
| rs7575022 | 2 | 237,964,265 | 2 | 1.2E-03 |  | rs7943891 | 11 | 48,843,790 | 1 | 8.2E-03 |  |
| rs74371893 | 3 | 4,345,484 | 3 | 1.6E-03 |  | rs75296154 | 11 | 69,343,815 | 2 | 9.3E-22 |  |
| rs76997204 | 3 | 20,027,251 | 1 | 1.9E-03 |  | rs151013524 | 11 | 134,062,648 | 1 | 3.7E-03 |  |
| rs75783758 | 4 | 137,761,414 | 1 | 1.3E-03 |  | rs3803466 | 15 | 75,648,650 | 1 | 1.7E-03 |  |
| rs148833559 | 5 | 172,755,066 | 2 | 9.3E-05 |  | rs75531903 | 16 | 3,613,720 | 2 | 8.5E-03 |  |
| rs16893699 | 6 | 28,105,889 | 4 | 1.5E-05 |  | rs4784227 | 16 | 52,599,188 | 1 | 2.3E-69 |  |
| rs2841646 | 6 | 43,270,326 | 1 | 1.6E-03 |  | rs61757659 | 16 | 88,951,594 | 1 | 7.1E-05 |  |
| rs41302073 | 9 | 12,709,125 | 1 | 1.7E-03 |  | rs16989263 | 21 | 33,694,195 | 4 | 5.0E-04 |  |

**Table 44:** Type 2 diabetes SNPs with SSV > 1% of total SSV

| SNP | Chromo-<br>-some | GRCh37<br>position | # of<br>CV folds | Average<br>gwas p-val | Near known<br>assoc. genes | SNP | Chromo-<br>-some | GRCh37<br>position | # of<br>CV folds | Average<br>gwas p-val | Near known<br>assoc. genes |
| --- | --- | --- | --- | --- | --- | --- | --- | --- | --- | --- | --- |
| rs74066236 | 1 | 37,919,465 | 2 | 5.5E-03 |  | rs7074231 | 10 | 3,304,976 | 1 | 7.9E-03 |  |
| rs156258 | 1 | 45,042,530 | 1 | 3.4E-03 |  | rs75106042 | 10 | 114,651,604 | 1 | 1.4E-07 | TCF7L2 |
| rs116473331 | 1 | 50,486,231 | 3 | 7.5E-05 |  | rs7903146 | 10 | 114,758,349 | 3 | 4.4E-143 | TCF7L2 |
| rs139971696 | 1 | 150,935,216 | 1 | 1.3E-02 |  | rs34796596 | 11 | 67,414,455 | 4 | 1.1E-03 |  |
| rs145020579 | 1 | 223,972,058 | 2 | 2.1E-03 |  | rs61001398 | 12 | 22,040,823 | 1 | 2.6E-03 |  |
| rs79802002 | 1 | 247,654,493 | 1 | 1.1E-01 |  | rs139495835 | 12 | 57,642,464 | 1 | 4.7E-02 |  |
| rs200225276 | 2 | 27,262,648 | 2 | 3.9E-05 | GCKR | rs73380117 | 12 | 100,346,464 | 2 | 7.3E-03 |  |
| rs143491198 | 2 | 27,695,208 | 2 | 8.8E-04 | GCKR | rs79111014 | 13 | 43,463,378 | 1 | 2.3E-03 |  |
| rs6715188 | 2 | 125,359,946 | 1 | 8.6E-03 |  | rs35716003 | 14 | 22,580,806 | 1 | 8.8E-03 |  |
| rs78499613 | 2 | 210,705,365 | 2 | 5.7E-05 |  | rs143602956 | 14 | 101,201,112 | 1 | 9.3E-03 |  |
| rs72628104 | 3 | 24,006,497 | 1 | 2.7E-02 |  | rs74553953 | 15 | 49,048,132 | 2 | 1.4E-05 | SLC12A1 |
| rs150271072 | 3 | 57,631,400 | 1 | 2.1E-04 |  | rs34437030 | 16 | 29,859,305 | 1 | 8.9E-03 |  |
| rs146206905 | 3 | 120,347,344 | 2 | 3.2E-04 |  | rs6500304 | 16 | 48,134,784 | 1 | 2.6E-03 |  |
| rs16837181 | 3 | 125,878,980 | 1 | 1.4E-04 |  | rs6500305 | 16 | 48,134,856 | 2 | 2.8E-03 |  |
| rs61753468 | 3 | 129,156,151 | 1 | 5.1E-04 |  | rs35840072 | 16 | 83,288,120 | 2 | 1.3E-04 |  |
| rs144574896 | 4 | 22,456,492 | 1 | 3.0E-04 |  | rs149760662 | 17 | 5,291,126 | 1 | 5.0E-04 |  |
| rs75978835 | 4 | 120,501,178 | 1 | 4.1E-03 |  | rs9748611 | 18 | 14,763,987 | 1 | 3.5E-02 |  |
| rs146886108 | 5 | 14,751,305 | 2 | 9.2E-08 |  | rs35554127 | 18 | 21,736,486 | 1 | 3.8E-02 |  |
| rs141074846 | 5 | 132,209,649 | 2 | 1.0E-03 |  | rs150579280 | 19 | 16,006,361 | 2 | 6.2E-04 |  |
| rs115673052 | 6 | 83,106,667 | 1 | 2.8E-02 |  | rs114897412 | 20 | 4,162,837 | 1 | 8.7E-03 |  |
| rs3802177 | 8 | 118,185,025 | 1 | 3.7E-21 | SLC30A8 | rs141568926 | 20 | 9,434,049 | 1 | 1.6E-03 |  |
| Affx-52298270 | 9 | 87,338,515 | 1 | 4.9E-03 |  | rs34875296 | 22 | 26,423,535 | 2 | 3.9E-03 |  |

**Table 45:**
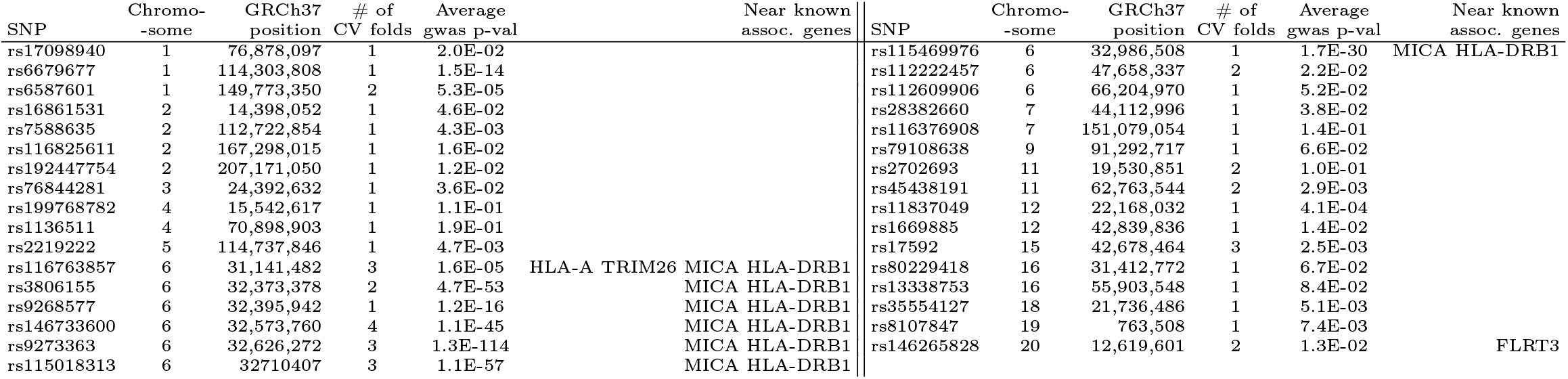
Type 1 diabetes SNPs with SSV > 1% of total SSV.

**Table 46:** CAD SNPs with SSV > 1% of total SSV.

| SNP | Chromo-<br>-some | GRCh37<br>position | # of<br>CV folds | Average<br>gwas p-val | Near known<br>assoc. genes | SNP | Chromo-<br>-some | GRCh37<br>position | # of<br>CV folds | Average<br>gwas p-val | Near known<br>assoc. genes |
| --- | --- | --- | --- | --- | --- | --- | --- | --- | --- | --- | --- |
| rs78426182 | 1 | 2,519,810 | 2 | 9.2E-04 |  | rs3734280 | 6 | 134,215,690 | 1 | 9.6E-03 | TCF21 |
| rs34561376 | 1 | 55,464,986 | 1 | 1.7E-03 | PCSK9 | rs3798220 | 6 | 160,961,137 | 2 | 3.0E-44 | SLC22A3 LPAL2 LPA PLG |
| rs11591147 | 1 | 55,505,647 | 2 | 5.9E-11 | PCSK9 | rs10455872 | 6 | 161,010,118 | 3 | 1.8E-73 | SLC22A3 LPAL2 LPA PLG |
| rs142460316 | 1 | 117,619,377 | 1 | 1.3E-04 |  | rs17847173 | 7 | 8,181,578 | 1 | 1.7E-04 |  |
| rs76443098 | 1 | 155,657,890 | 1 | 1.2E-02 | IL6R | rs60659894 | 8 | 94,772,183 | 3 | 2.2E-02 |  |
| rs76329326 | 1 | 222,803,199 | 1 | 9.8E-05 | MIA3 | rs116930274 | 8 | 144,590,049 | 1 | 1.2E-03 |  |
| rs73949680 | 2 | 74,756,242 | 3 | 1.4E-02 |  | rs7126678 | 11 | 48,505,913 | 1 | 2.0E-01 |  |
| rs115370220 | 2 | 87,016,830 | 2 | 1.4E-02 | VAMP5 | rs34796596 | 11 | 67,414,455 | 1 | 4.4E-02 |  |
| rs79904664 | 2 | 144,699,925 | 3 | 7.8E-03 | ZEB2 | rs58168448 | 12 | 109,895,854 | 1 | 4.9E-02 | SH2B3 |
| rs9827335 | 3 | 3,840,868 | 1 | 8.8E-03 |  | rs200297509 | 12 | 124,835,279 | 1 | 7.5E-03 |  |
| rs13326552 | 3 | 44,948,480 | 5 | 7.1E-06 |  | rs41561818 | 12 | 133,220,454 | 1 | 3.2E-04 |  |
| rs144662307 | 3 | 195,481,129 | 3 | 2.1E-04 |  | rs144567652 | 14 | 45,667,921 | 1 | 7.6E-03 |  |
| rs142412240 | 4 | 68,925,094 | 1 | 1.7E-04 |  | rs150953383 | 15 | 73,994,895 | 1 | 3.3E-04 |  |
| rs73786670 | 5 | 130,766,542 | 1 | 2.1E-02 | SLC22A4/A5 | rs16942445 | 16 | 57,935,443 | 1 | 6.5E-03 | CETP |
| rs80263204 | 5 | 140,627,509 | 1 | 1.3E-02 |  | rs58981829 | 19 | 51,738,465 | 1 | 2.4E-02 |  |
| rs2120783 | 5 | 161,007,916 | 1 | 9.8E-04 |  |  |  |  |  |  |  |

**Table 47:** Hypertension SNPs with SSV > 1% of total SSV.

| SNP | Chromo-<br>-some | GRCh37<br>position | # of<br>CV folds | Average<br>gwas p-val | Near known<br>assoc. genes | SNP | Chromo-<br>-some | GRCh37<br>position | # of<br>CV folds | Average<br>gwas p-val | Near known<br>assoc. genes |
| --- | --- | --- | --- | --- | --- | --- | --- | --- | --- | --- | --- |
| rs41270291 | 1 | 15,994,251 | 1 | 2.4E-01 |  | rs11016690 | 10 | 130,940,735 | 1 | 3.3E-01 |  |
| rs115862221 | 1 | 17,249,858 | 1 | 5.1E-02 |  | rs138242314 | 11 | 45,955,752 | 1 | 1.2E-02 |  |
| rs11466111 | 1 | 115,829,178 | 1 | 9.9E-06 |  | rs8187661 | 11 | 66,133,643 | 1 | 2.1E-02 |  |
| rs145467872 | 1 | 150,973,013 | 1 | 4.4E-02 |  | rs16914280 | 11 | 88,321,724 | 1 | 1.8E-01 |  |
| rs55909005 | 1 | 156,838,432 | 1 | 2.5E-01 |  | rs11838918 | 13 | 79,410,574 | 1 | 3.9E-01 |  |
| rs9661539 | 1 | 248,040,293 | 1 | 1.9E-01 |  | rs7317657 | 13 | 92,171,706 | 1 | 4.0E-01 |  |
| rs45612738 | 2 | 31,602,841 | 1 | 2.6E-01 |  | rs73296180 | 14 | 53,619,369 | 1 | 2.2E-02 |  |
| rs61731210 | 3 | 13,659,649 | 1 | 5.3E-01 |  | rs112118955 | 14 | 73,996,947 | 1 | 1.5E-01 |  |
| rs17051692 | 3 | 40,931,175 | 1 | 1.9E-01 | ULK4 | rs116488414 | 14 | 74,006,004 | 1 | 6.6E-02 |  |
| rs4234213 | 3 | 123,024,733 | 1 | 9.1E-02 |  | rs149239345 | 14 | 96,761,321 | 1 | 1.3E-01 |  |
| rs57958890 | 4 | 70,276,699 | 1 | 2.2E-01 |  | rs6576507 | 15 | 26,288,394 | 1 | 6.4E-01 |  |
| rs143057152 | 4 | 149,075,755 | 1 | 4.9E-03 | NR3C2 | rs114231576 | 16 | 2,016,184 | 1 | 5.9E-02 |  |
| rs6915612 | 6 | 32,218,625 | 1 | 2.8E-01 | PRRC2A | rs4889238 | 16 | 81,151,122 | 1 | 5.1E-02 |  |
| rs112460091 | 6 | 63,995,477 | 1 | 1.0E-01 |  | rs112966915 | 17 | 80,223,554 | 1 | 1.9E-01 |  |
| rs59147126 | 7 | 15,601,393 | 1 | 2.8E-01 |  | rs150705131 | 19 | 288,123 | 1 | 3.1E-02 |  |
| rs3735533 | 7 | 27,245,893 | 1 | 4.2E-08 |  | rs61740630 | 19 | 2,352,964 | 1 | 1.8E-01 |  |
| rs115281203 | 7 | 123,672,179 | 1 | 1.6E-01 |  | rs36078704 | 19 | 19,039,030 | 1 | 2.3E-01 |  |
| rs11977216 | 7 | 150,324,976 | 1 | 3.2E-01 | NOS3 | rs73932907 | 19 | 38,039,946 | 1 | 2.0E-01 |  |
| rs116751013 | 7 | 158,415,596 | 1 | 7.3E-02 |  | rs201635014 | 19 | 38,379,680 | 1 | 1.2E-01 |  |
| rs10958553 | 8 | 39,290,593 | 1 | 2.9E-01 |  | rs61760904 | 19 | 50,139,932 | 1 | 1.1E-03 |  |
| rs148545964 | 8 | 94,772,206 | 1 | 5.8E-01 |  | rs55641738 | 20 | 60,293,875 | 1 | 2.8E-01 |  |
| rs3847170 | 8 | 140,423,147 | 1 | 4.2E-01 |  | rs28587162 | 21 | 16,337,538 | 1 | 1.5E-01 |  |
| rs3025380 | 9 | 136,501,756 | 1 | 1.4E-03 |  | rs6003859 | 22 | 24,057,206 | 1 | 2.7E-01 |  |
| rs73270587 | 10 | 35,929,350 | 1 | 1.9E-01 |  | rs28915381 | 22 | 43,089,658 | 1 | 2.6E-01 |  |

**Table 48:** Direct bilirubin SNPs with SSV > 1% of total SSV.

| SNP | Chromo-<br>-some | GRCh37<br>position | # of<br>CV folds | Average<br>gwas p-val | Near known<br>assoc. genes | SNP | Chromo-<br>-some | GRCh37<br>position | # of<br>CV folds | Average<br>gwas p-val | Near known<br>assoc. genes |
| --- | --- | --- | --- | --- | --- | --- | --- | --- | --- | --- | --- |
| rs17131137 | 1 | 91,172,059 | 2 | 3.2E-04 |  | Affx-52351697 | 12 | 21,008,080 | 4 | 1.1E-39 | SLCO1B3 SLCO1B1 |
| rs4019811 | 1 | 203,822,304 | 4 | 1.8E-05 |  | rs34691116 | 12 | 21,027,327 | 3 | 2.3E-89 | SLCO1B3 SLCO1B1 |
| rs908327 | 1 | 235,092,600 | 4 | 1.6E-02 |  | rs76737149 | 12 | 21,235,850 | 2 | 2.0E-16 | SLCO1B3 SLCO1B1 |
| rs6755571 | 2 | 234,627,536 | 3 | 0.0E+00 | UGT1A1 | rs11045819 | 12 | 21,329,813 | 2 | 1.6E-28 | SLCO1B3 SLCO1B1 |
| rs34622615 | 2 | 234,652,308 | 4 | 0.0E+00 | UGT1A1 | rs4149056 | 12 | 21,331,549 | 2 | 7.9E-308 | SLCO1B3 SLCO1B1 |
| rs10929302 | 2 | 234,665,782 | 2 | 0.0E+00 | UGT1A1 | rs73117071 | 12 | 41,582,272 | 3 | 1.2E-03 |  |
| rs3755319 | 2 | 234,667,582 | 4 | 0.0E+00 | UGT1A1 | rs61742753 | 16 | 1,614,147 | 3 | 4.3E-05 |  |
| rs887829 | 2 | 234,668,570 | 5 | 0.0E+00 | UGT1A1 | rs114221795 | 20 | 42,086,724 | 2 | 1.5E-07 |  |
| rs4148323 | 2 | 234,669,144 | 5 | 6.6E-03 | UGT1A1 | rs8132639 | 21 | 27,012,154 | 5 | 2.7E-06 |  |
| rs2290189 | 3 | 13,677,946 | 1 | 1.2E-03 |  | rs16989263 | 21 | 33,694,195 | 2 | 1.1E-05 |  |
| rs142626656 | 8 | 41,575,677 | 2 | 1.8E-07 |  | rs35946782 | 21 | 40,763,754 | 1 | 1.9E-03 |  |

**Table 49:** BMI SNPs with SSV > 1% of total SSV.

| SNP | Chromo-<br>-some | GRCh37<br>position | # of<br>CV folds | Average<br>gwas p-val | Near known<br>assoc. genes | SNP | Chromo-<br>-some | GRCh37<br>position | # of<br>CV folds | Average<br>gwas p-val | Near known<br>assoc. genes |
| --- | --- | --- | --- | --- | --- | --- | --- | --- | --- | --- | --- |
| rs201703264 | 1 | 3428144 | 2 | 4.2E-04 |  | rs77620102 | 8 | 21986657 | 2 | 2.9E-03 |  |
| rs74066236 | 1 | 37919465 | 2 | 5.5E-03 |  | rs74062407 | 12 | 11083647 | 2 | 4.9E-03 |  |
| rs62106258 | 2 | 417167 | 3 | 1.2E-70 | TMEM18 | rs141556486 | 12 | 132404616 | 1 | 4.3E-05 |  |
| rs73019479 | 2 | 171627219 | 1 | 8.6E-03 |  | rs1421085 | 16 | 53800954 | 2 | 3.0E-262 | FTO |
| rs4685689 | 3 | 3886580 | 3 | 5.3E-03 |  | rs143242388 | 16 | 70380869 | 1 | 9.2E-03 |  |
| rs9843741 | 3 | 99568737 | 2 | 4.7E-04 |  | rs1060250 | 16 | 87874736 | 4 | 2.5E-03 |  |
| rs2871630 | 3 | 129372880 | 1 | 6.7E-03 |  | rs10405385 | 19 | 55856147 | 1 | 3.7E-04 |  |
| rs72976362 | 3 | 133567847 | 1 | 7.3E-03 |  | rs112799437 | 19 | 58609112 | 2 | 7.6E-04 |  |
| rs76693682 | 5 | 180482838 | 2 | 4.1E-03 |  | rs62224618 | 22 | 16057417 | 2 | 6.9E-01 |  |
| rs2229107 | 7 | 87138659 | 2 | 1.1E-03 |  | rs34379045 | 22 | 19511476 | 3 | 6.0E-03 |  |

**Table 50:** Height SNPs with SSV > 1% of total SSV.

| SNP | Chromo-<br>-some | GRCh37<br>position | # of<br>CV folds | Average<br>gwas p-val | Near known<br>assoc. genes | SNP | Chromo-<br>-some | GRCh37<br>position | # of<br>CV folds | Average<br>gwas p-val | Near known<br>assoc. genes |
| --- | --- | --- | --- | --- | --- | --- | --- | --- | --- | --- | --- |
| rs201823506 | 1 | 51,871,568 | 1 | 1.4E-03 | ORC1 | rs35796392 | 9 | 129,595,583 | 2 | 1.8E-05 |  |
| rs10019684 | 4 | 69,383,874 | 1 | 2.5E-03 |  | rs10987622 | 9 | 130,133,619 | 1 | 2.3E-03 |  |
| rs142228984 | 5 | 32,786,413 | 1 | 8.9E-09 |  | rs28407189 | 15 | 89,400,680 | 2 | 3.0E-135 |  |
| rs148833559 | 5 | 172,755,066 | 2 | 4.9E-29 |  | rs141308595 | 15 | 89,424,870 | 4 | 9.2E-30 |  |
| rs41271299 | 6 | 19,839,415 | 1 | 1.6E-157 |  | rs1362317 | 16 | 6,250,379 | 2 | 8.3E-05 |  |
| rs74841643 | 6 | 34,163,292 | 2 | 3.4E-37 | COL11A2 FANCE | rs4889238 | 16 | 81,151,122 | 3 | 2.9E-04 |  |
| rs9470004 | 6 | 35,341,850 | 1 | 2.1E-07 | FANCE | rs202127176 | 16 | 88,782,205 | 1 | 5.7E-15 | ANKRD11 CDK10 CDT1 FANCA GALN |
| Affx-29864875 | 7 | 142,331,582 | 1 | 4.7E-04 | BRAF | rs58680048 | 19 | 51,870,706 | 1 | 1.2E-03 |  |
| rs112892337 | 8 | 135,614,553 | 1 | 8.7E-49 |  |  |  |  |  |  |  |

**Table 51:** Lipoprotein A SNPs with SSV > 1% of total SSV.

| SNP | Chromo-<br>-some | GRCh37<br>position | # of<br>CV folds | Average<br>gwas p-val | Near known<br>assoc. genes | SNP | Chromo-<br>-some | GRCh37<br>position | # of<br>CV folds | Average<br>gwas p-val | Near known<br>assoc. genes |
| --- | --- | --- | --- | --- | --- | --- | --- | --- | --- | --- | --- |
| rs146534110 | 6 | 160,578,069 | 2 | 0.0E+00 | LPA LPAL2 SLC22A3 | rs74617384 | 6 | 160,997,118 | 2 | 0.0E+00 | LPA LPAL2 SLC22A3 |
| rs16891156 | 6 | 160,608,804 | 4 | 0.0E+00 | LPA LPAL2 SLC22A3 | rs41272114 | 6 | 161,006,077 | 1 | 3.0E-209 | LPA LPAL2 SLC22A3 |
| rs8177505 | 6 | 160,679,656 | 2 | 0.0E+00 | LPA LPAL2 SLC22A3 | rs41272112 | 6 | 161,006,105 | 1 | 1.4E-31 | LPA LPAL2 SLC22A3 |
| rs10080815 | 6 | 160,687,412 | 1 | 0.0E+00 | LPA LPAL2 SLC22A3 | rs147936725 | 6 | 161,007,619 | 3 | 1.3E-13 | LPA LPAL2 SLC22A3 |
| rs421913 | 6 | 160,742,369 | 1 | 1.3E-09 | LPA LPAL2 SLC22A3 | rs10455872 | 6 | 161,010,118 | 5 | 0.0E+00 | LPA LPAL2 SLC22A3 |
| rs540713 | 6 | 160,766,321 | 3 | 2.2E-117 | LPA LPAL2 SLC22A3 | rs41272078 | 6 | 161,010,150 | 2 | 1.3E-89 | LPA LPAL2 SLC22A3 |
| rs3918291 | 6 | 160,828,142 | 2 | 0.0E+00 | LPA LPAL2 SLC22A3 | rs140306630 | 6 | 161,013,826 | 3 | 6.4E-09 | LPA LPAL2 SLC22A3 |
| rs12214416 | 6 | 160,910,517 | 1 | 0.0E+00 | LPA LPAL2 SLC22A3 | rs73596816 | 6 | 161,017,363 | 5 | 0.0E+00 | LPA LPAL2 SLC22A3 |
| rs72501790 | 6 | 160,915,856 | 3 | 1.6E-18 | LPA LPAL2 SLC22A3 | rs41259144 | 6 | 161,022,107 | 2 | 6.9E-62 | LPA LPAL2 SLC22A3 |
| rs41267807 | 6 | 160,952,816 | 1 | 3.0E-149 | LPA LPAL2 SLC22A3 | rs143079629 | 6 | 161,128,812 | 1 | 1.1E-33 | LPA LPAL2 SLC22A3 |
| rs3798220 | 6 | 160,961,137 | 5 | 0.0E+00 | LPA LPAL2 SLC22A3 | rs4252128 | 6 | 161,152,819 | 3 | 1.6E-11 | LPA LPAL2 SLC22A3 |
| rs116089584 | 6 | 160,962,370 | 1 | 6.7E-65 | LPA LPAL2 SLC22A3 | rs4252152 | 6 | 161,159,366 | 5 | 0.0E+00 | LPA LPAL2 SLC22A3 |
| rs7767084 | 6 | 160,962,503 | 1 | 1.8E-76 | LPA LPAL2 SLC22A3 |  |  |  |  |  |  |

## G Predictor variance

### G.1 Fraction of variance explained

**Figure 27:**
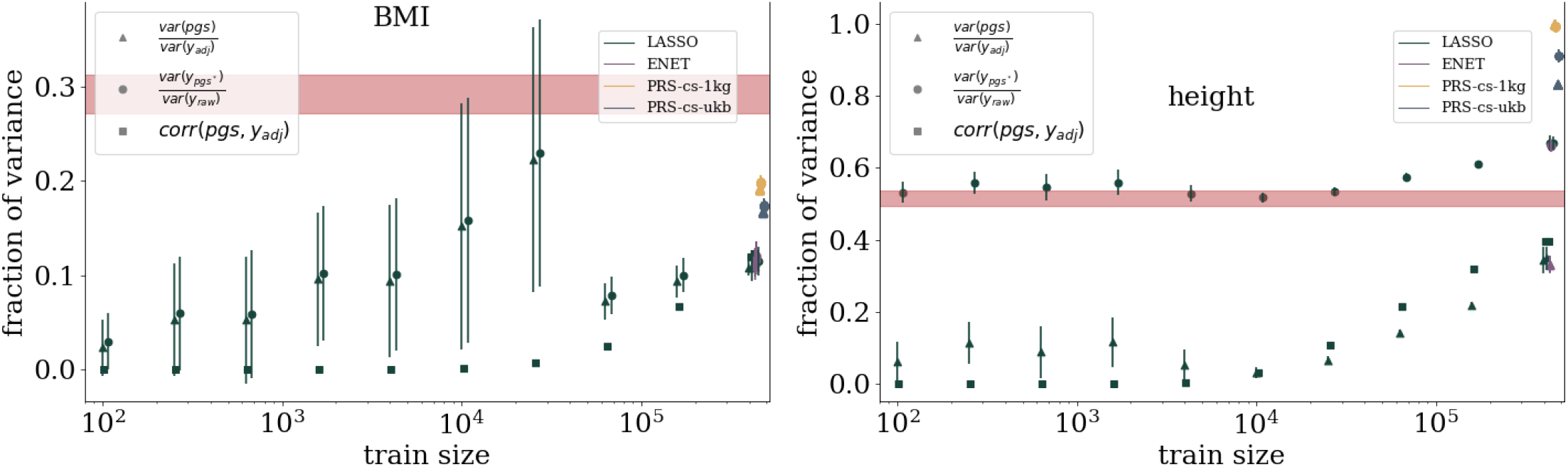
Fraction of variance explained estimates for BMI and height. Red band is the result from using GCTA.

**Figure 28:**
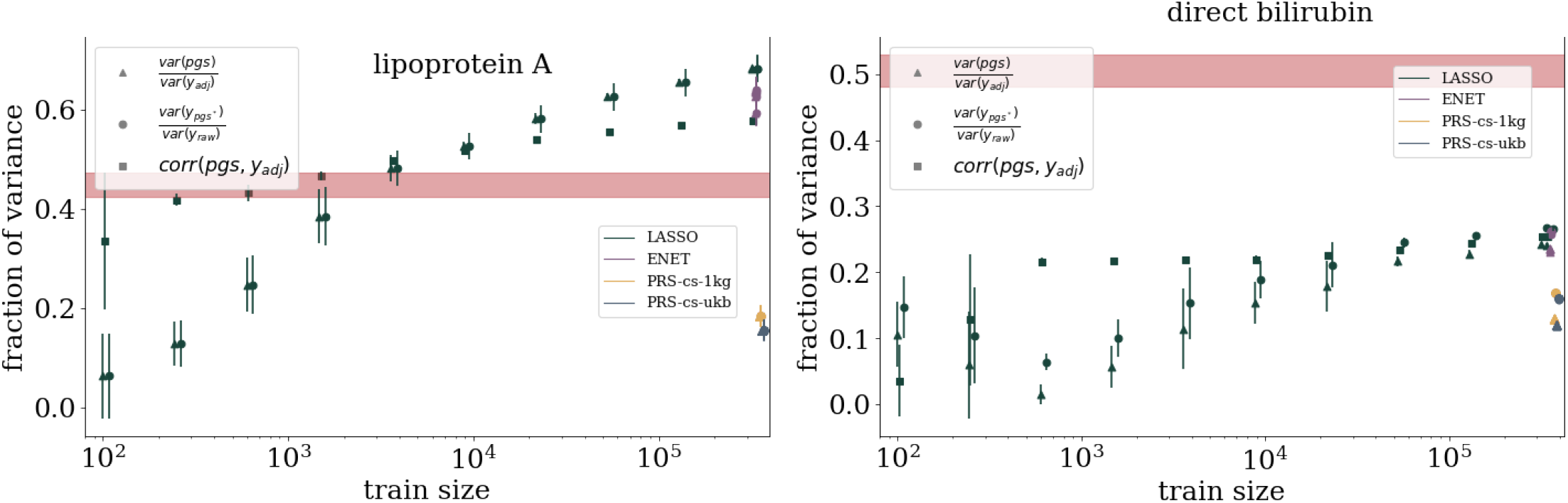
Fraction of variance explained estimates for lipoprotein A and direct bilirubin. Red band is the result from using GCTA.

**Figure 29:**
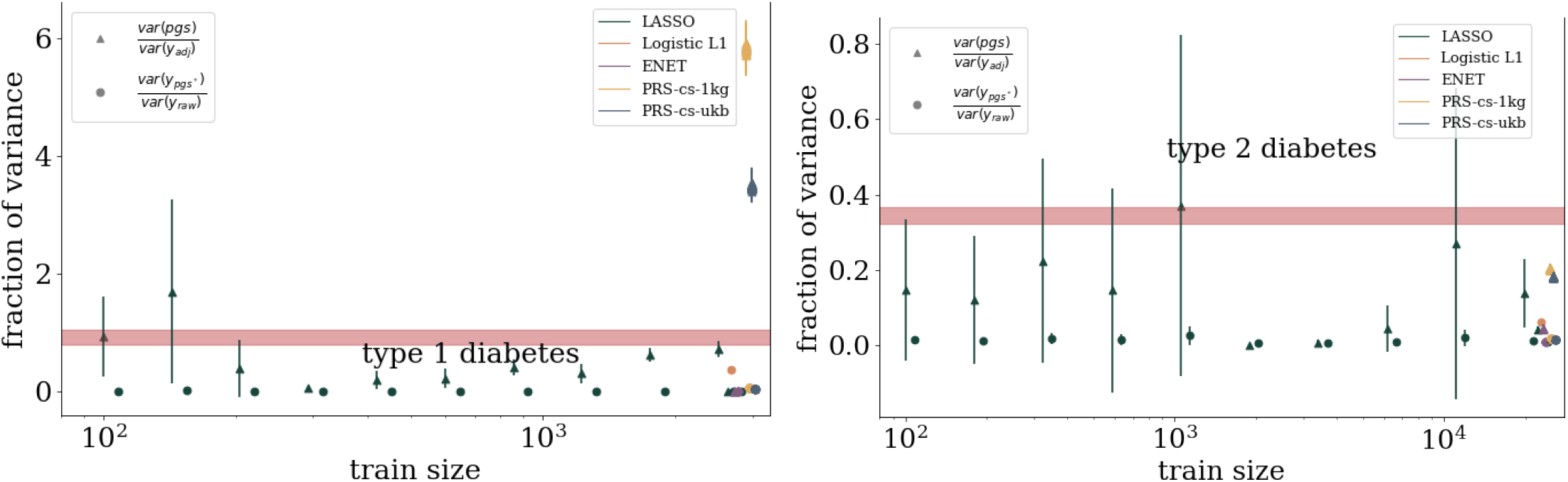
Fraction of variance explained estimates for diabetes. Red band is the result from using GCTA.

**Figure 30:**
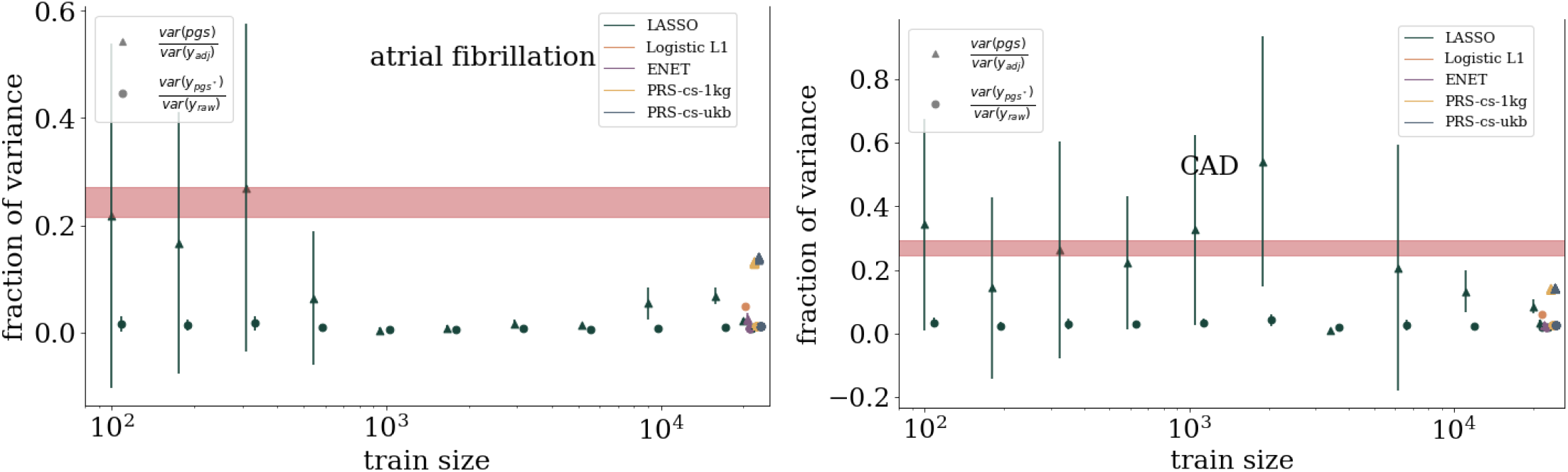
Fraction of variance explained estimates for atrial fibrillation and CAD. Red band is the result from using GCTA.

**Figure 31:**
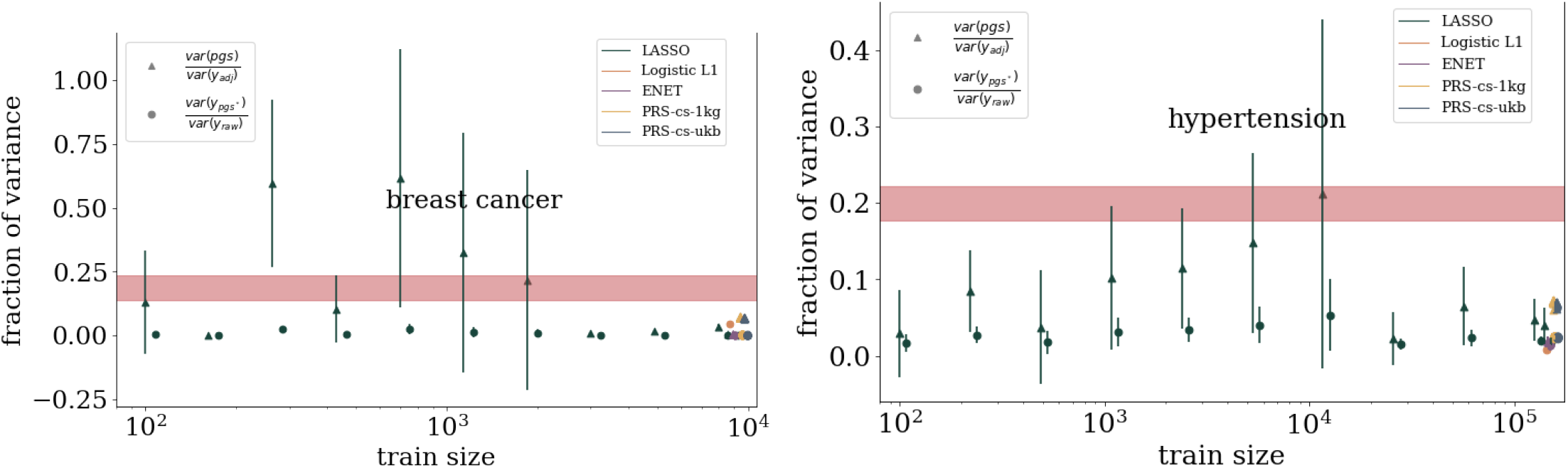
Fraction of variance explained estimates for breast cancer and hypertension. Red band is the result from using GCTA.

**Figure 32:**
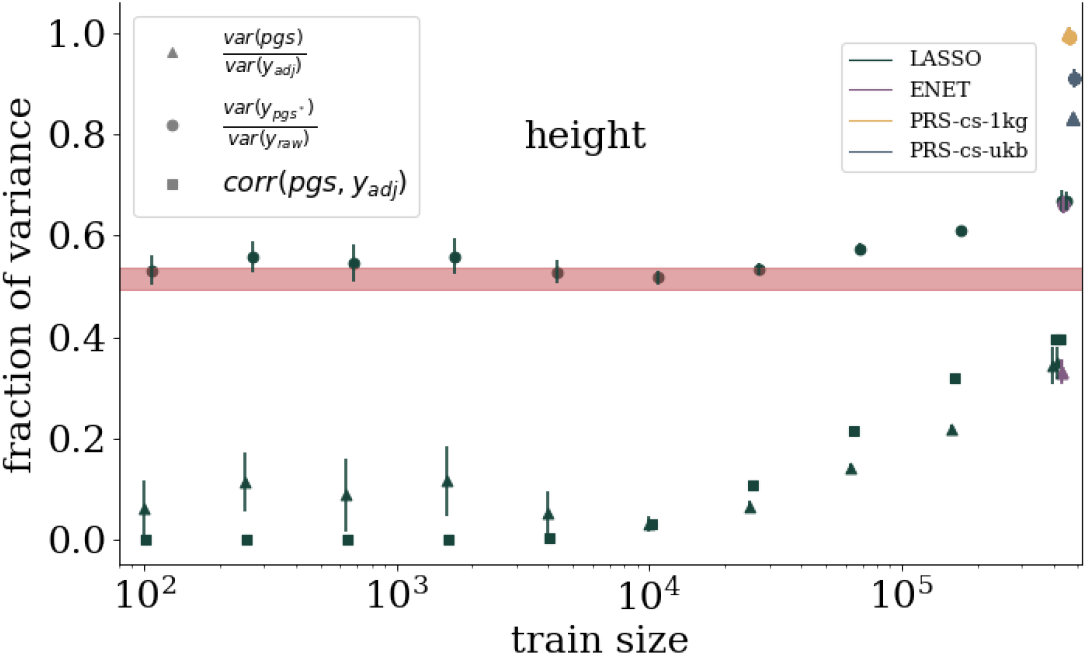
Fraction of variance explained estimates for height. Red band is the result from using GCTA.

### G.2 SSV vs total variance

**Figure 33:**
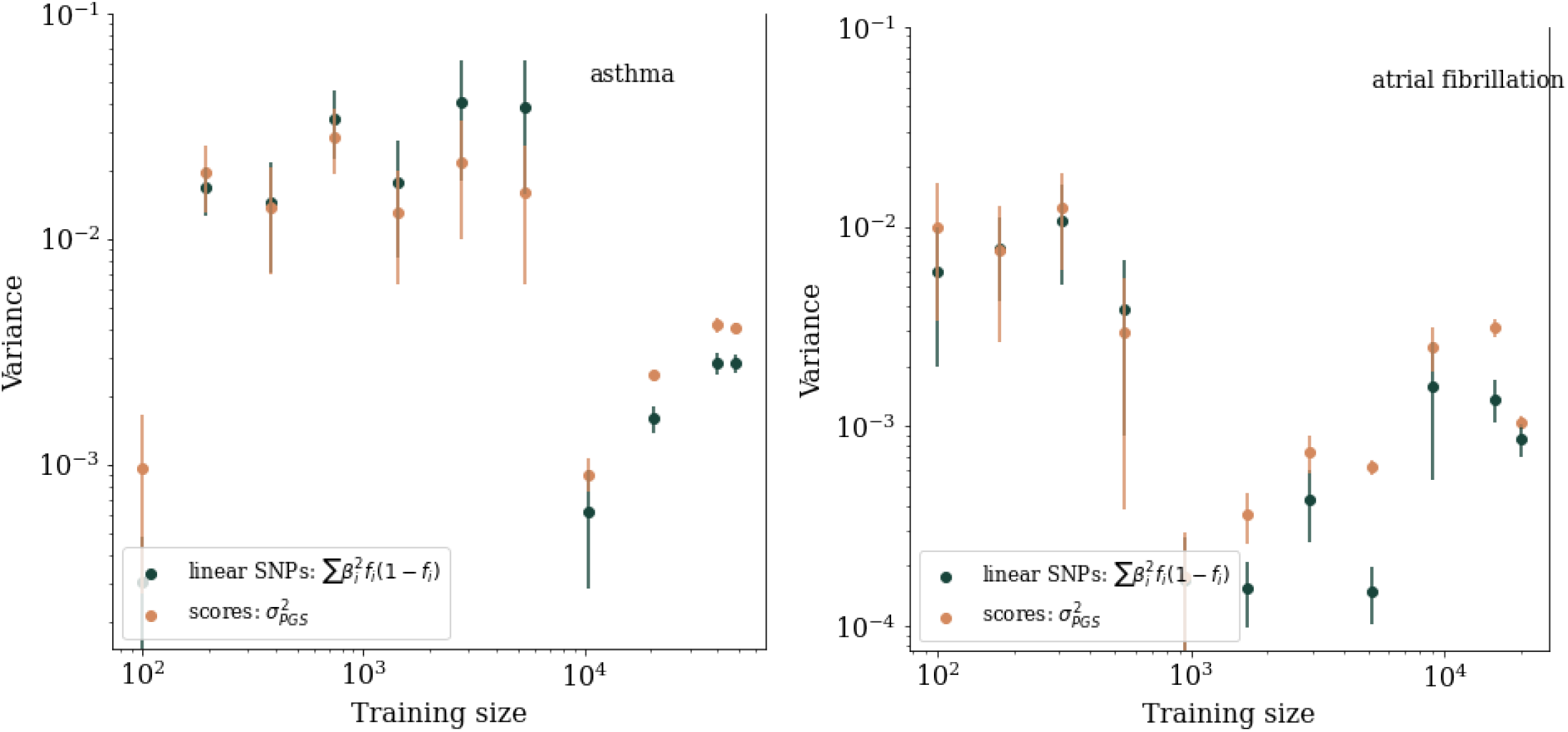
Comparison of SSV to total LASSO predictor variance for asthma and atrial fibrillation.

**Figure 34:**
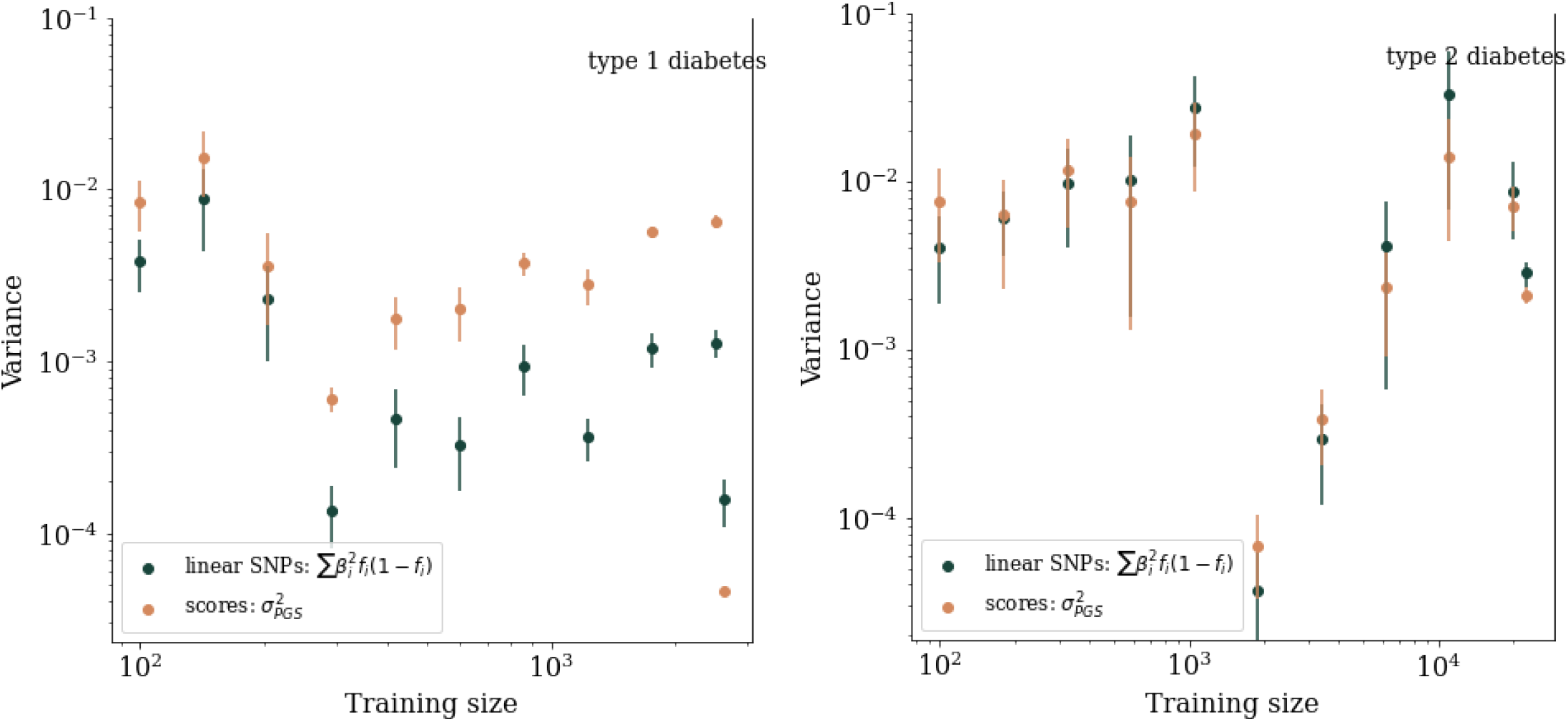
Comparison of SSV to total LASSO predictor variance for type 1 and 2 diabetes.

**Figure 35:**
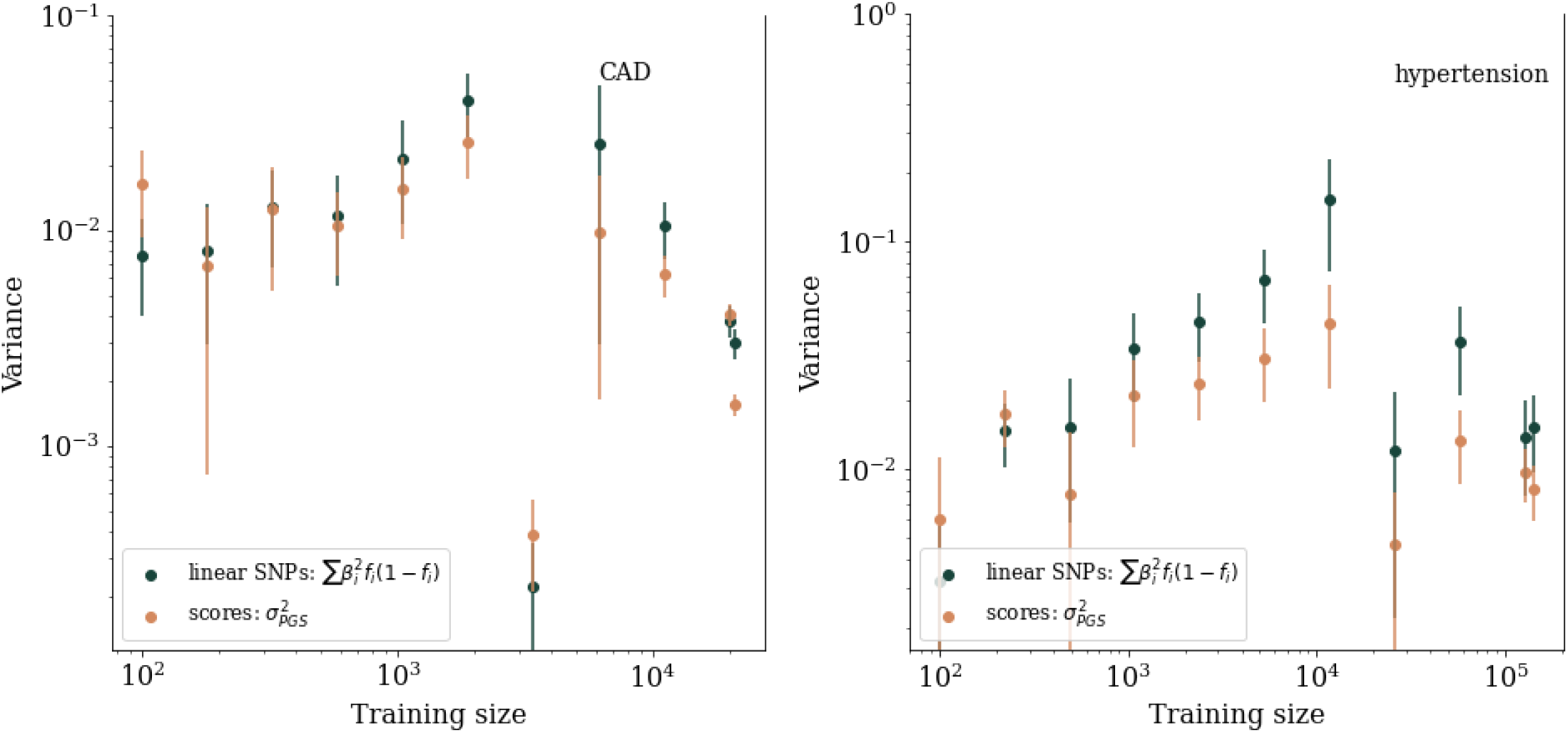
Comparison of SSV to total LASSO predictor variance for CAD and hypertension.

**Figure 36:**
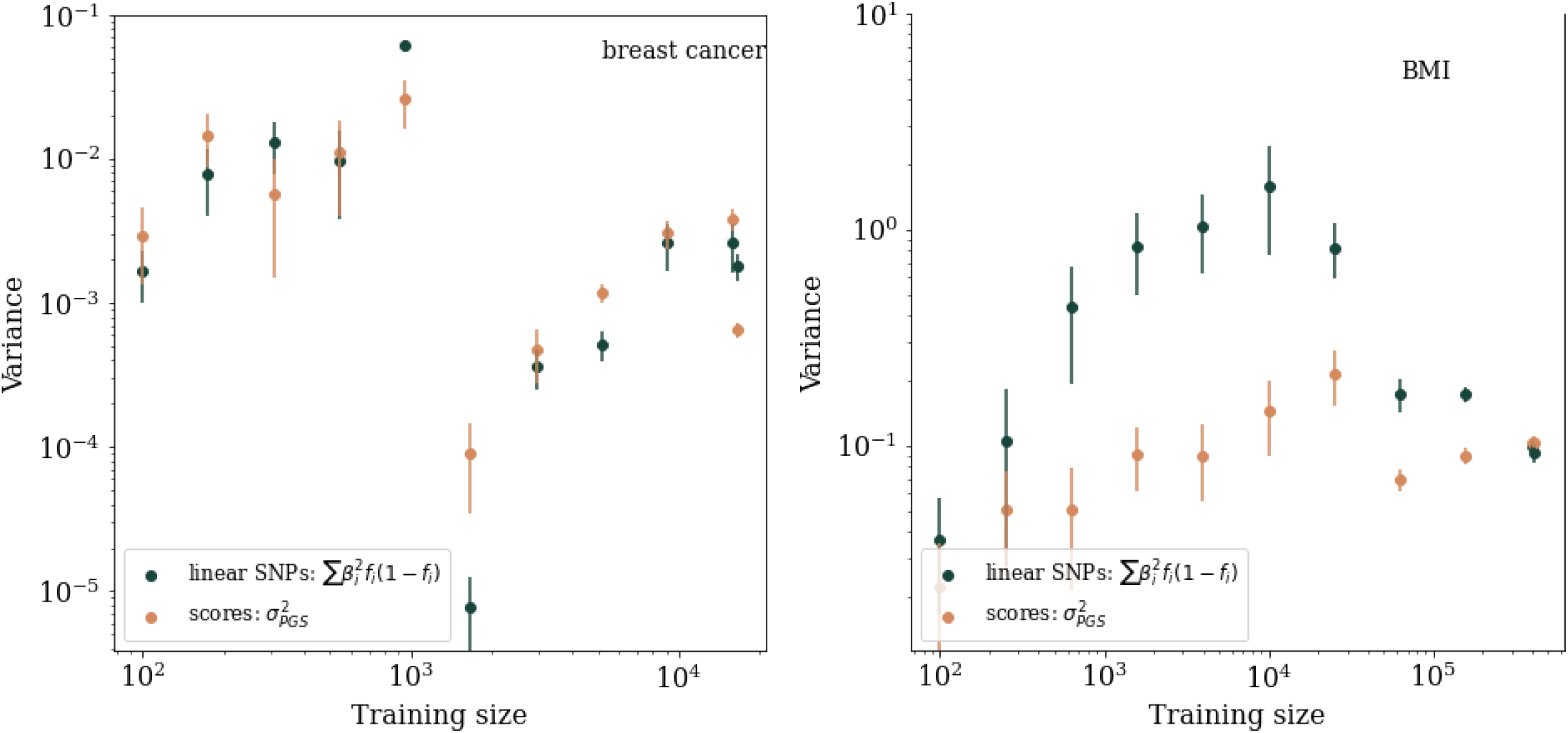
Comparison of SSV to total LASSO predictor variance for breast cancer and BMI.

**Figure 37:**
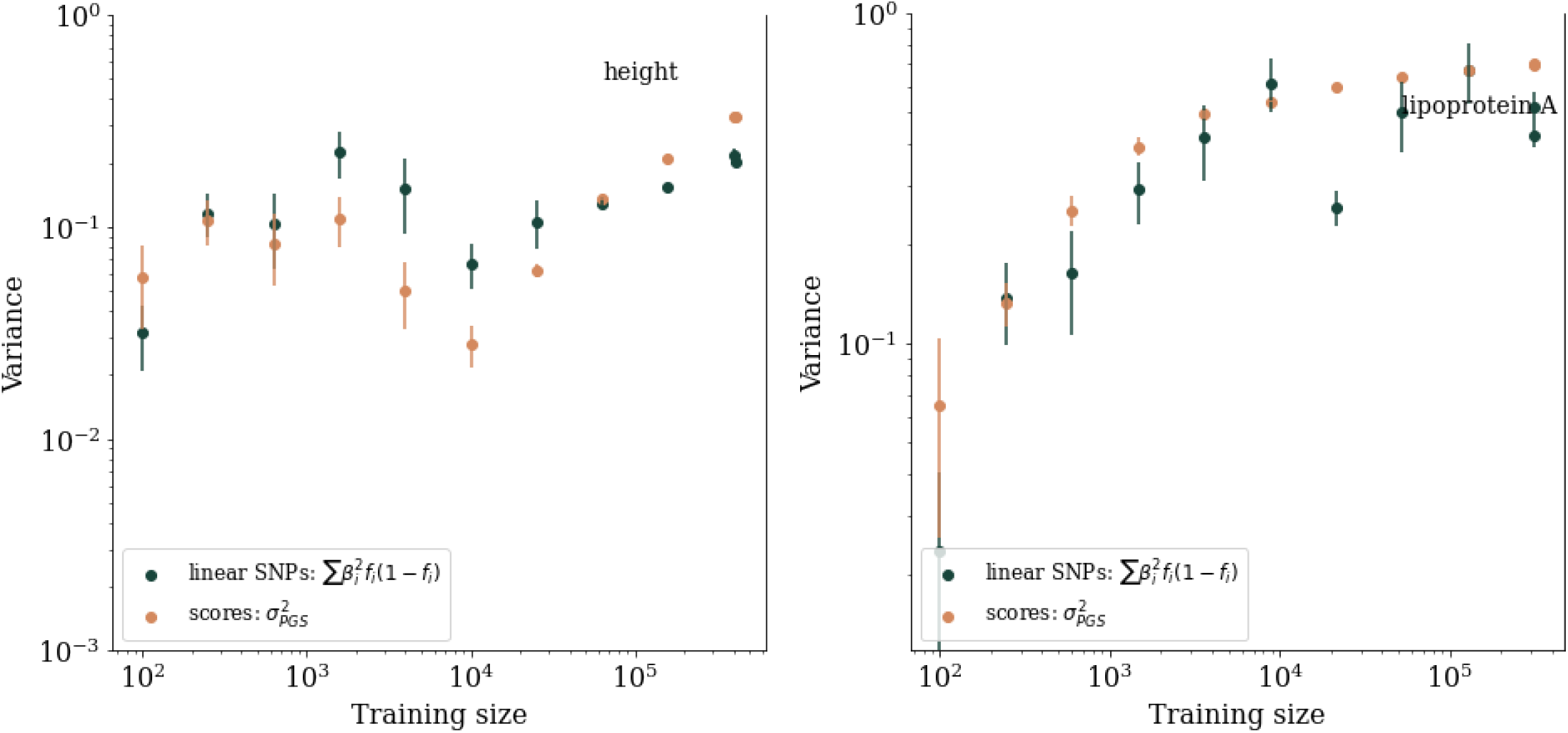
Comparison of SSV to total LASSO predictor variance for height and lipoprotein A.

**Figure 38:**
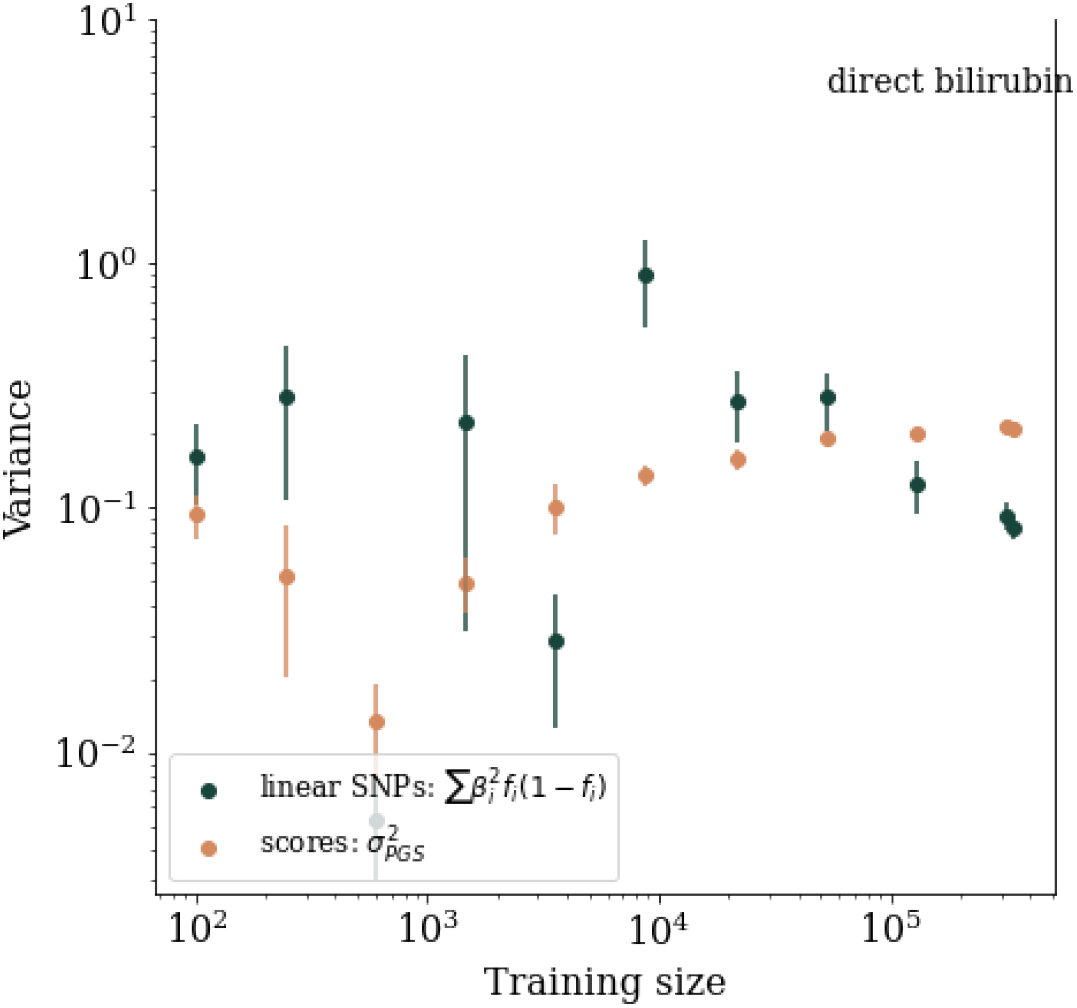
Comparison of SSV to total LASSO predictor variance for direct bilirubin.

### G.3 Predictor impact regions

**Figure 39:**
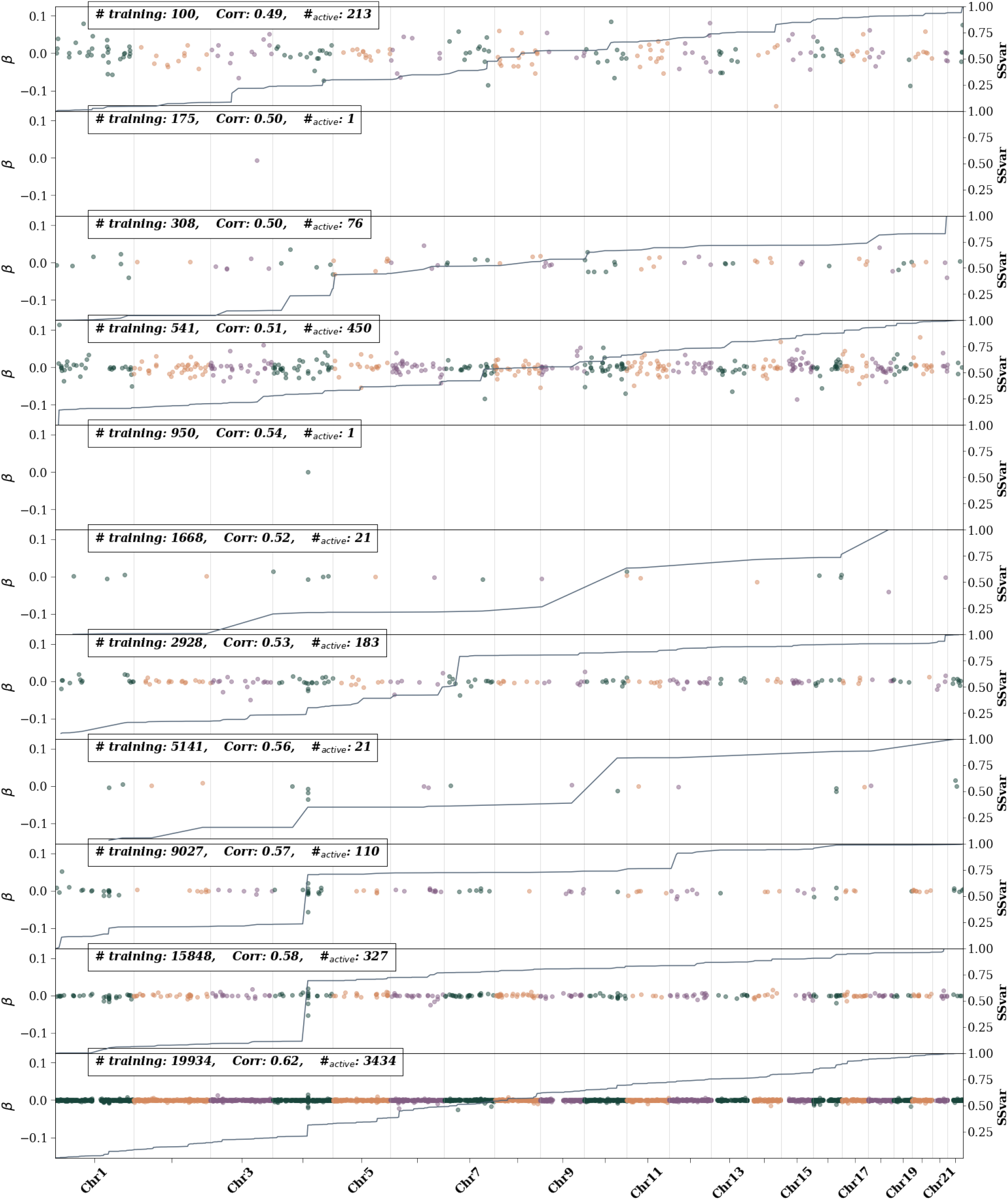
Atrial fibrillation active SNPs – i.e., SNPs with non-zero *β* weights– as training size is increased. The left axis shows the *β* value and is represented by colored dots. Different colors are used to differentiate chromosomes. The right axis represents the single SNP variance (SSV) normalized to the total SSV. The solid line showes the cumulative SSV. The “training” label represents the number of cases used in training. The first 10 (from the top) training sizes use equal number of cases and controls. The final training size uses all possible remaining controls.

**Figure 40:**
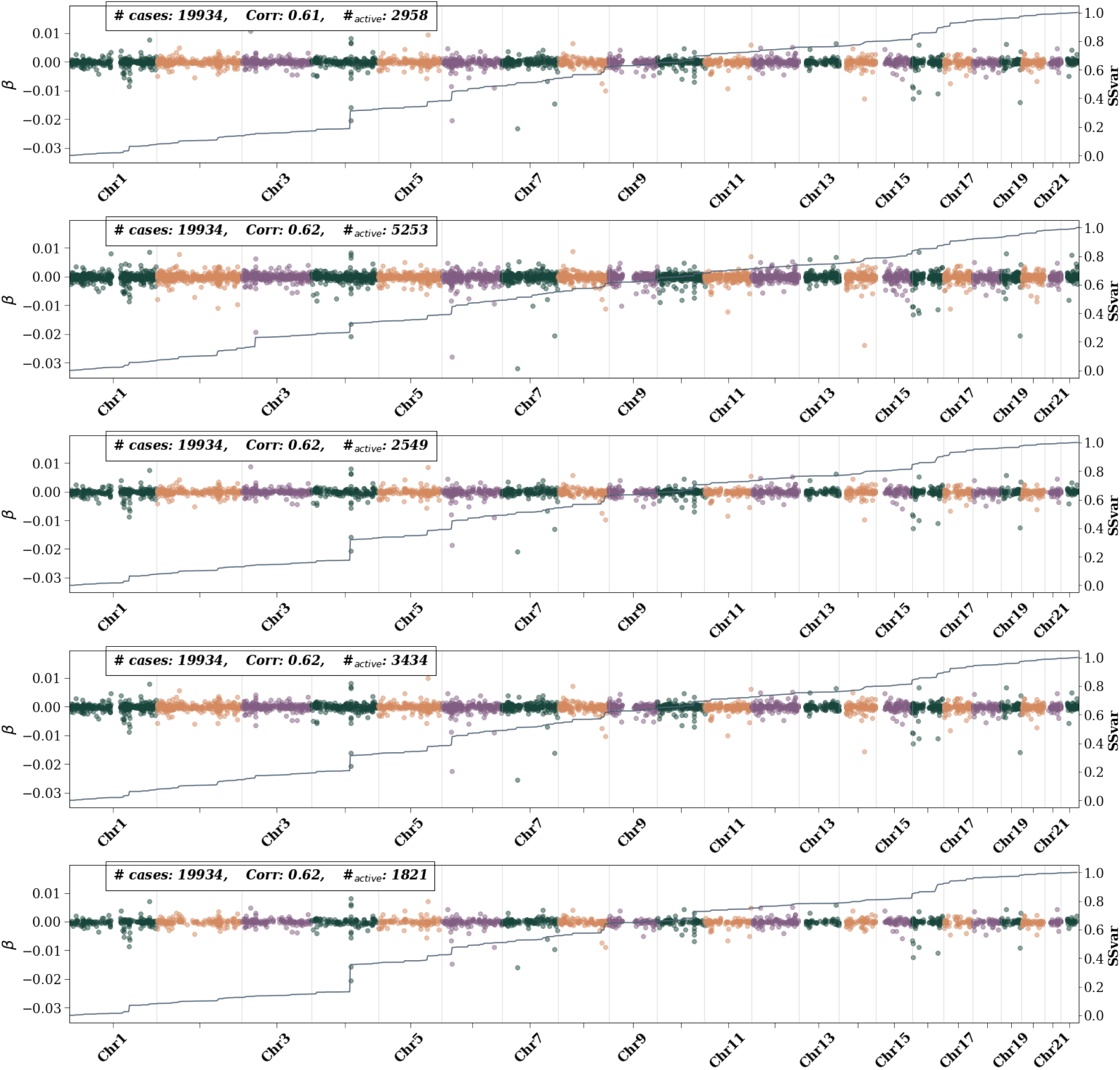
Atrial fibrillation active SNPs – i.e., SNPs with non-zero *β* weights– for 5 CV folds at maximum training size. Left axis shows the *β* value and is represented by colored dots. Different colors are used to differentiate chromosomes. The right axis represents the single SNP variance (SSV) normalized to the total SSV. The “training” label represents the number of cases used in training. All possible controls were used in each fold. While features generally appear consistent across folds, i.e., the presence of a bump in the SSV line, the size of the bump varies.

**Figure 41:**
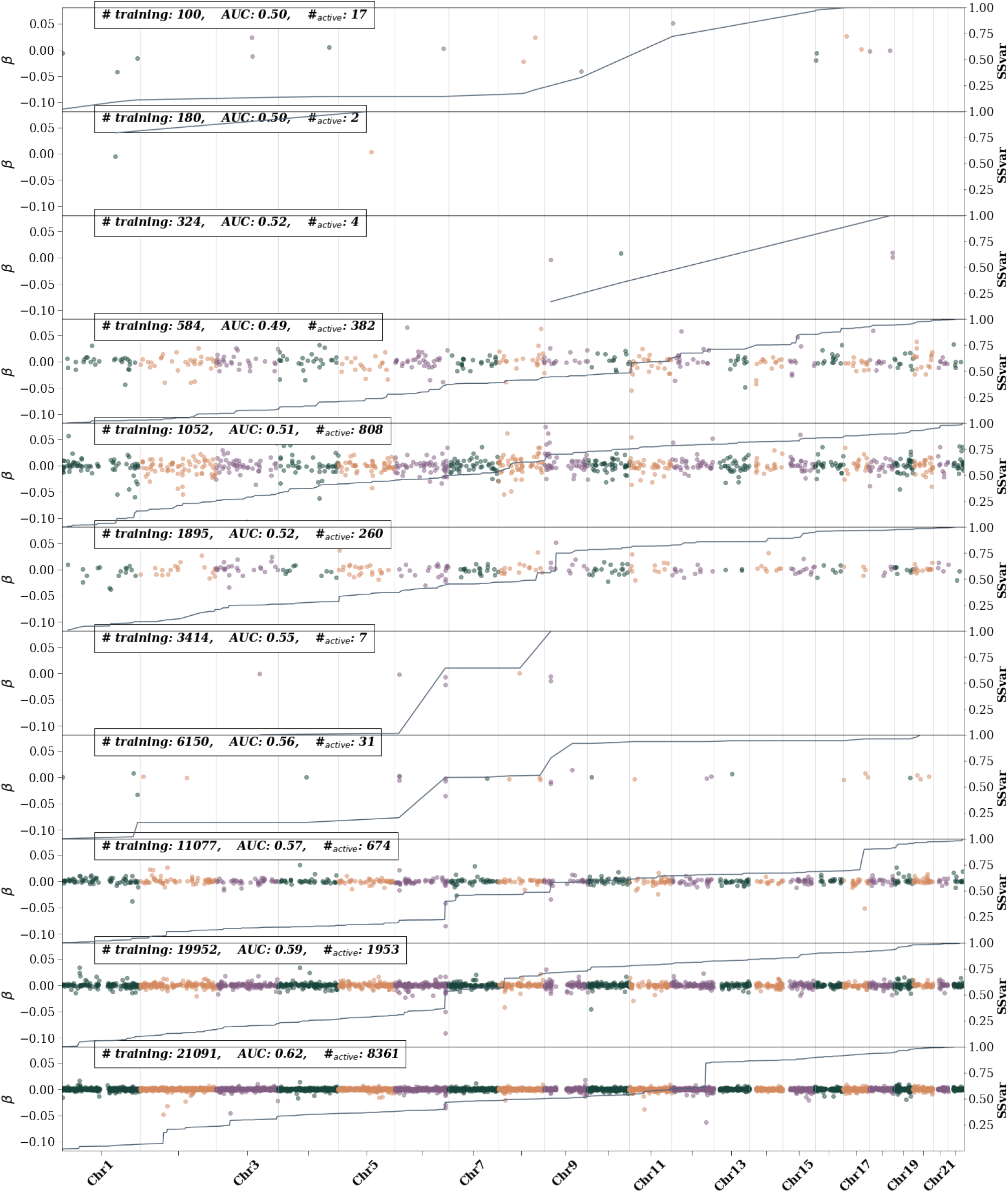
Coronary artery disease active SNPs – i.e., SNPs with non-zero *β* weights– as training size is increased. The left axis shows the *β* value and is represented by colored dots. Different colors are used to differentiate chromosomes. The right axis represents the single SNP variance (SSV) normalized to the total SSV. The solid line showes the cumulative SSV. The “training” label represents the number of cases used in training. The first 10 (from the top) training sizes use equal number of cases and controls. The final training size uses all possible remaining controls

**Figure 42:**
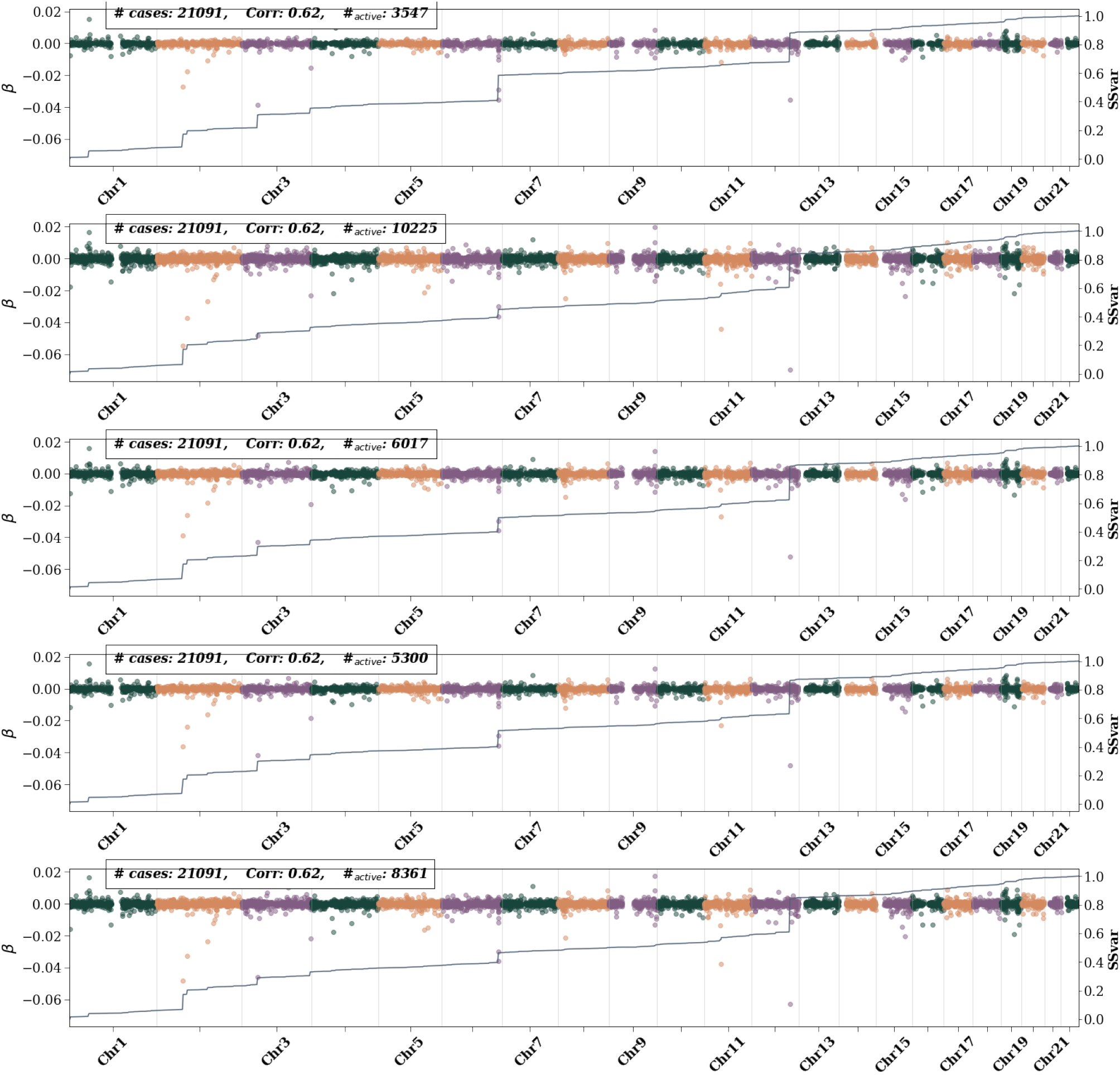
CAD active SNPs – i.e., SNPs with non-zero *β* weights– for 5 CV folds at maximum training size. Left axis shows the *β* value and is represented by colored dots. Different colors are used to differentiate chromosomes. The right axis represents the single SNP variance (SSV) normalized to the total SSV. The “training” label represents the number of cases used in training. All possible controls were used in each fold. While features generally appear consistent across folds, i.e., the presence of a bump in the SSV line, the size of the bump varies.

**Figure 43:**
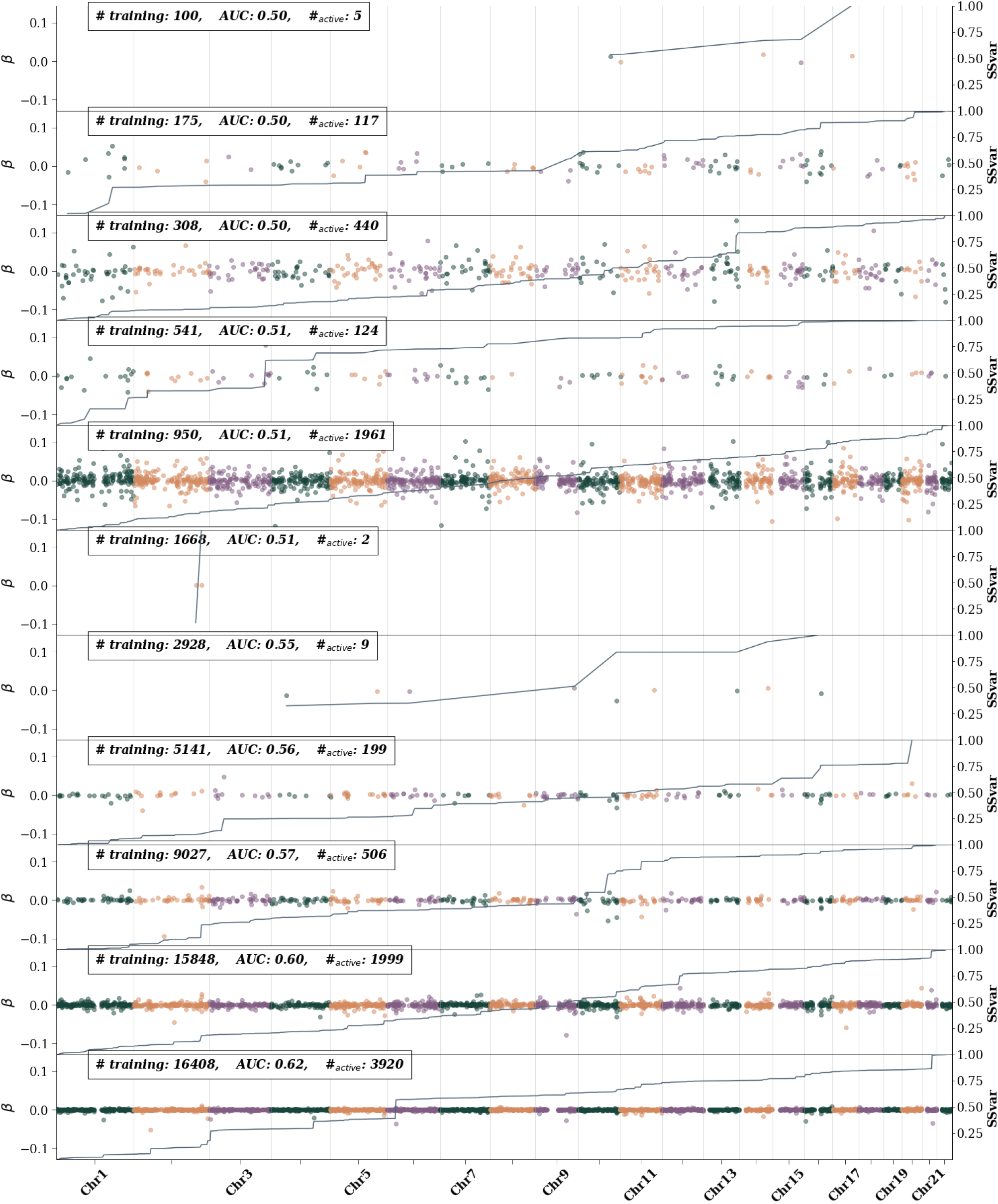
Breast cancer active SNPs – i.e., SNPs with non-zero *β* weights– as training size is increased. The left axis shows the *β* value and is represented by colored dots. Different colors are used to differentiate chromosomes. The right axis represents the single SNP variance (SSV) normalized to the total SSV. The solid line showes the cumulative SSV. The “training” label represents the number of cases used in training. The first 10 (from the top) training sizes use equal number of cases and controls. The final training size uses all possible remaining controls

**Figure 44:**
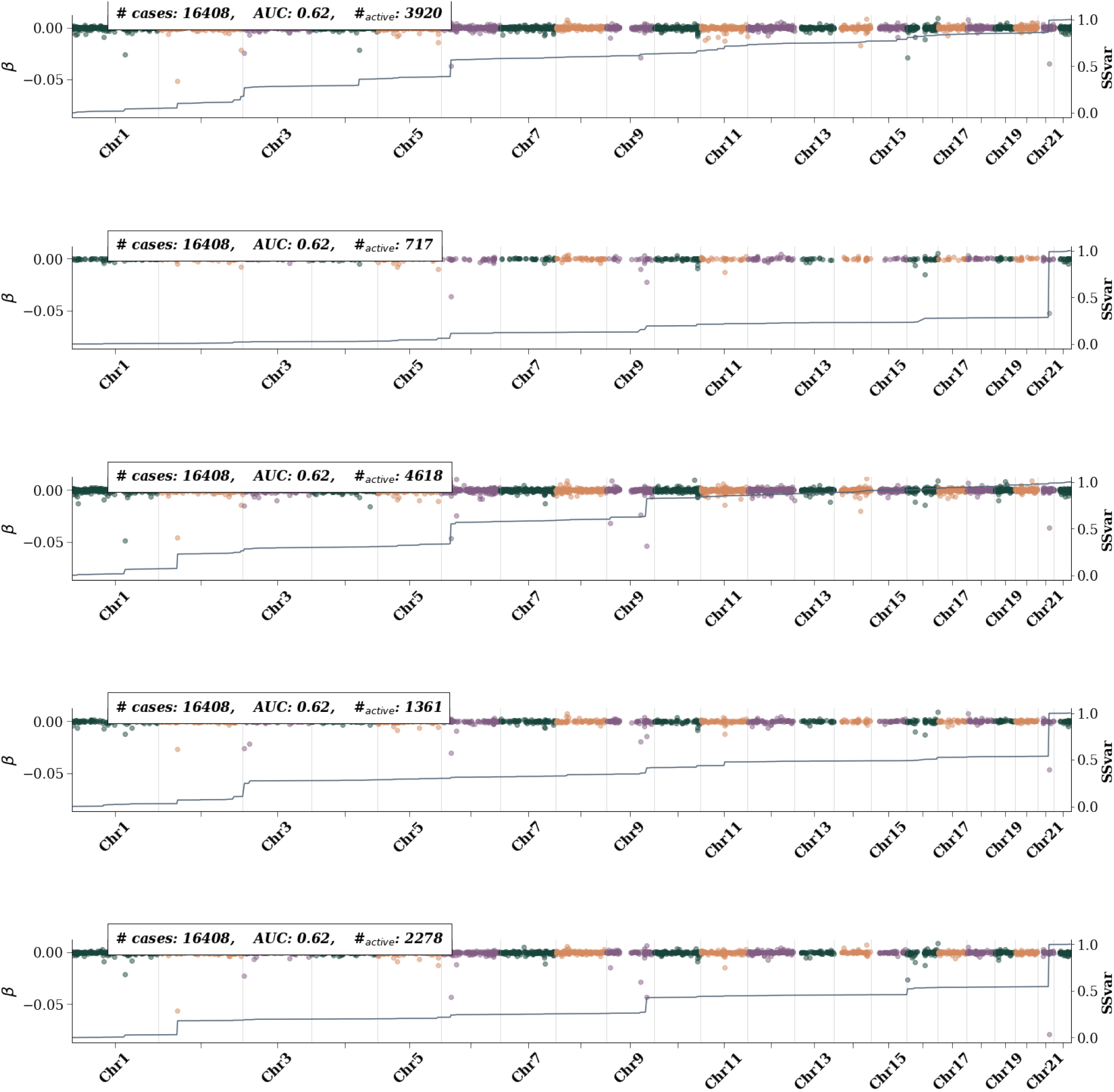
Breast cancer active SNPs – i.e., SNPs with non-zero *β* weights– for 5 CV folds at maximum training size. Left axis shows the *β* value and is represented by colored dots. Different colors are used to differentiate chromosomes. The right axis represents the single SNP variance (SSV) normalized to the total SSV. The “training” label represents the number of cases used in training. All possible controls were used in each fold. While features generally appear consistent across folds, i.e., the presence of a bump in the SSV line, the size of the bump varies.

**Figure 45:**
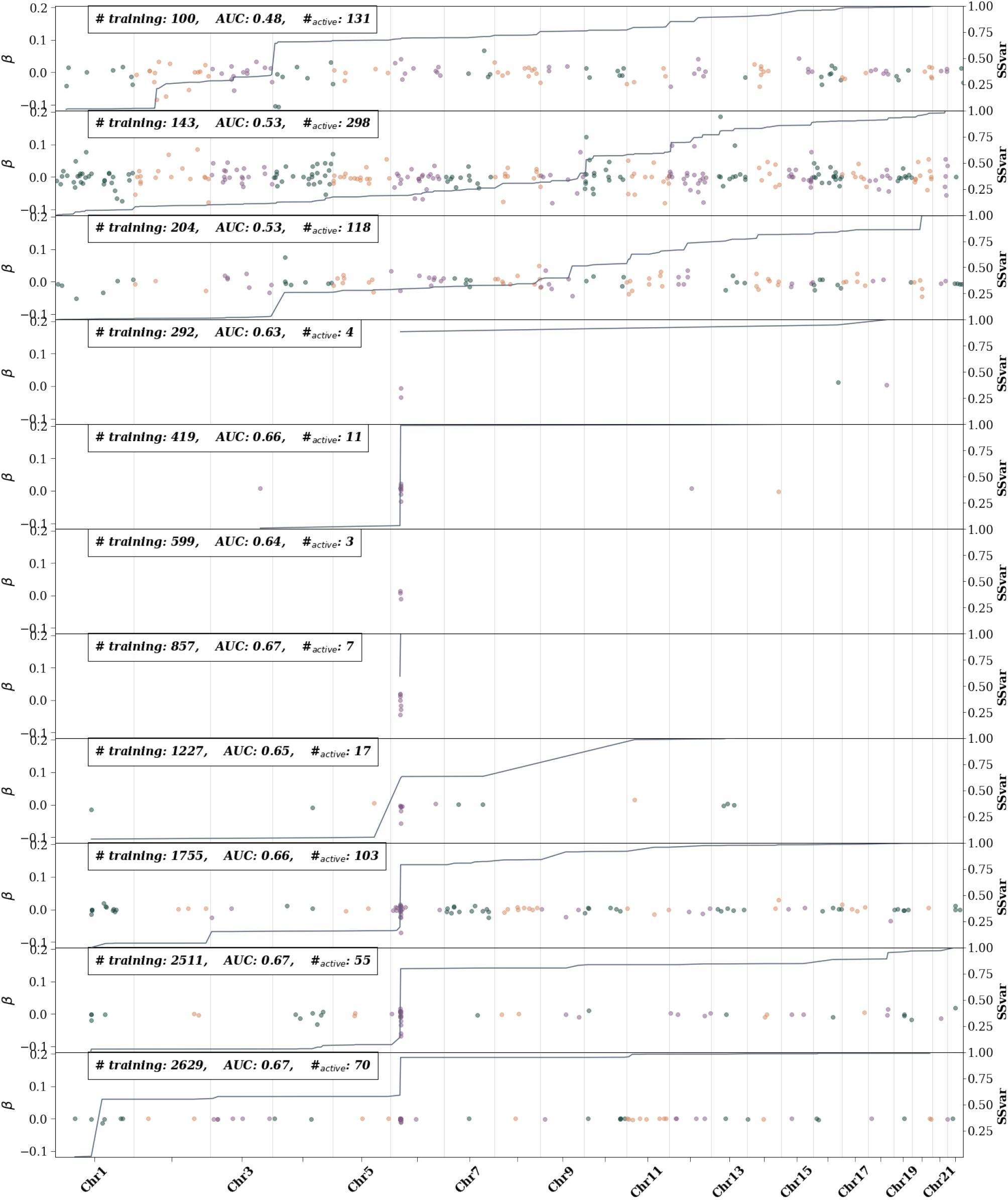
Type 1 diabetes active SNPs – i.e., SNPs with non-zero *β* weights– as training size is increased. The left axis shows the *β* value and is represented by colored dots. Different colors are used to differentiate chromosomes. The right axis represents the single SNP variance (SSV) normalized to the total SSV. The solid line showes the cumulative SSV. The “training” label represents the number of cases used in training. The first 10 (from the top) training sizes use equal number of cases and controls. The final training size uses all possible remaining controls

**Figure 46:**
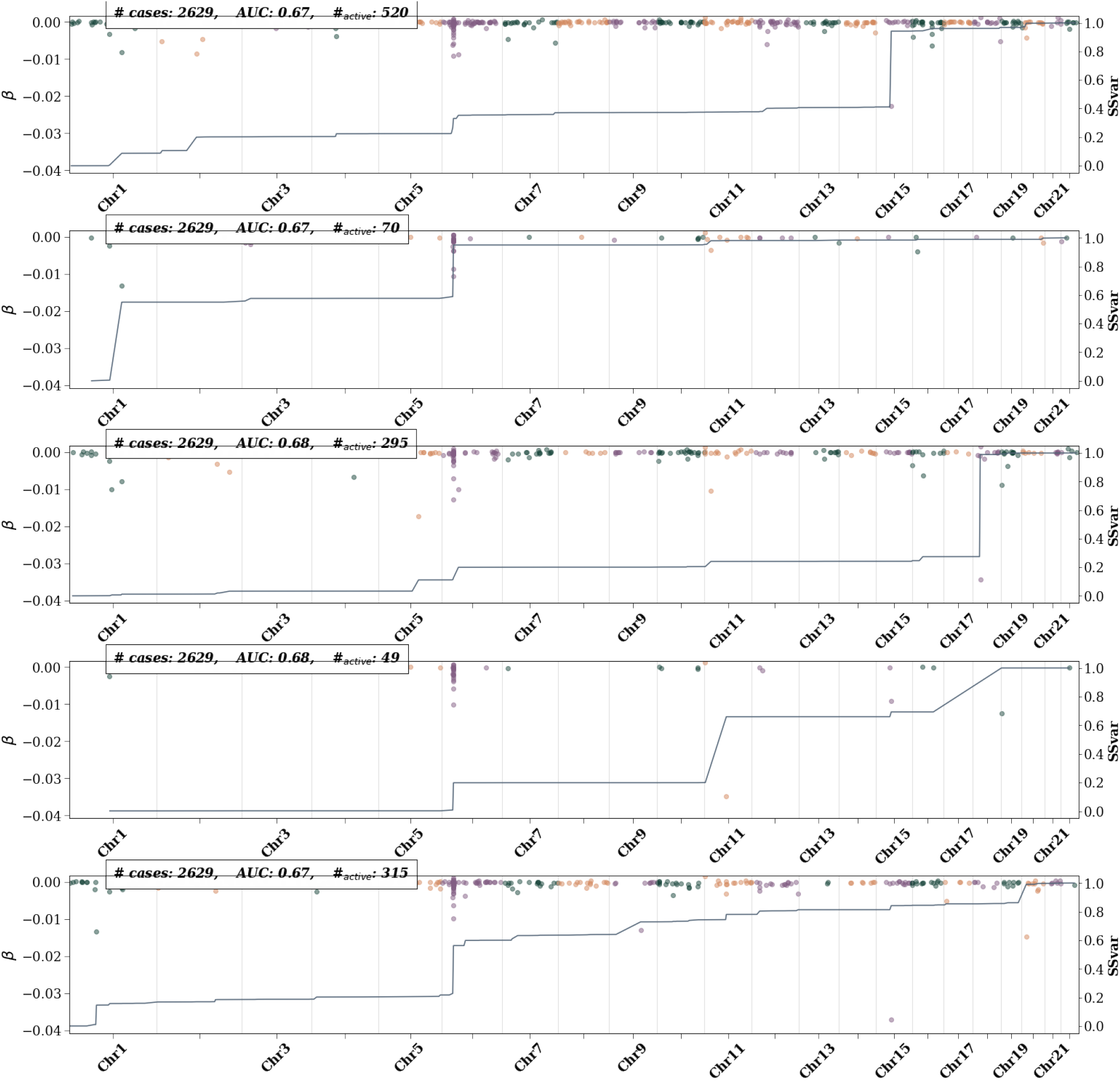
Type 1 diabetes active SNPs – i.e., SNPs with non-zero *β* weights– for 5 CV folds at maximum training size. Left axis shows the *β* value and is represented by colored dots. Different colors are used to differentiate chromosomes. The right axis represents the single SNP variance (SSV) normalized to the total SSV. The “training” label represents the number of cases used in training. All possible controls were used in each fold. While features generally appear consistent across folds, i.e., the presence of a bump in the SSV line, the size of the bump varies.

**Figure 47:**
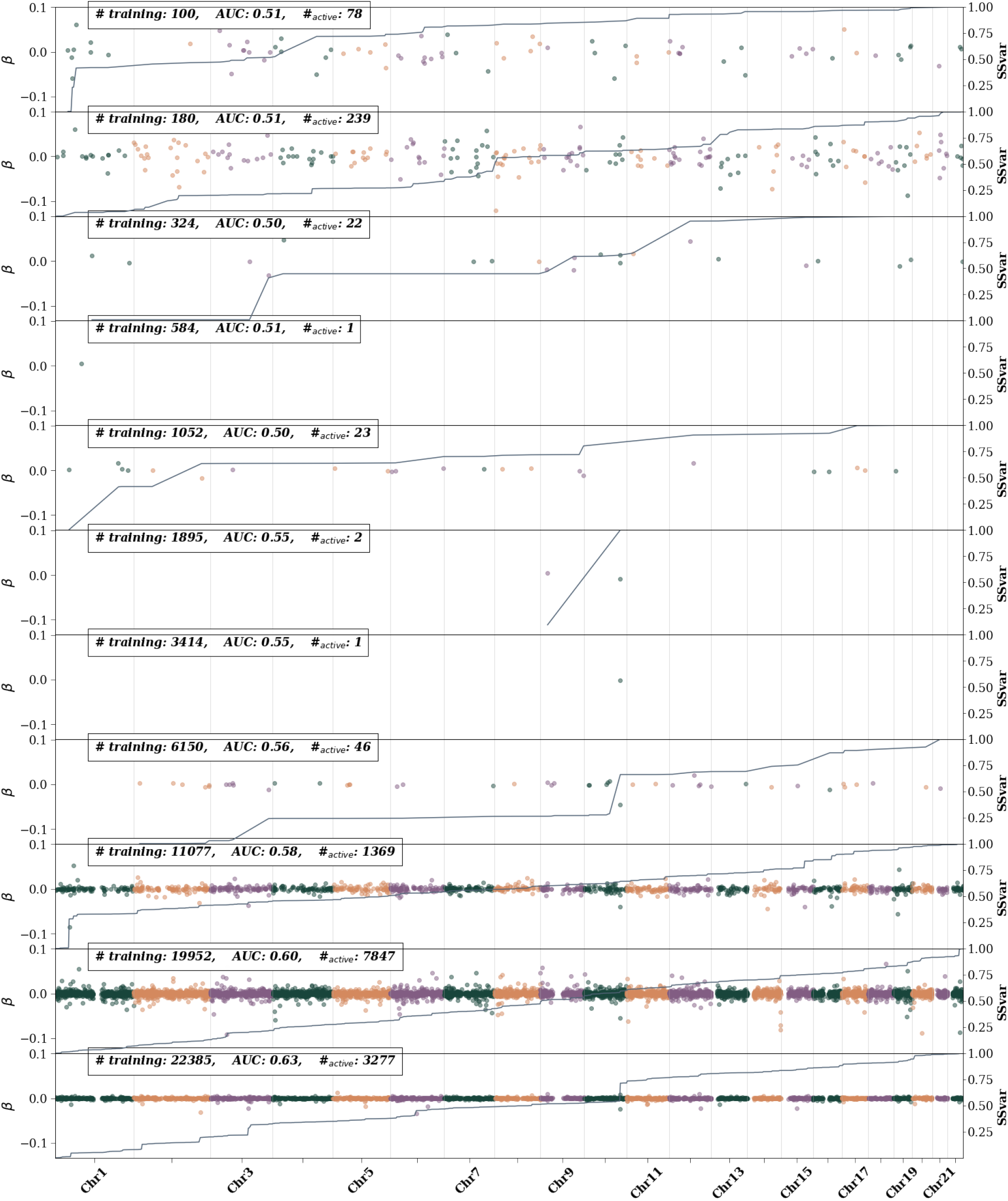
Type 2 diabetes active SNPs – i.e., SNPs with non-zero *β* weights– as training size is increased. The left axis shows the *β* value and is represented by colored dots. Different colors are used to differentiate chromosomes. The right axis represents the single SNP variance (SSV) normalized to the total SSV. The solid line showes the cumulative SSV. The “training” label represents the number of cases used in training. The first 10 (from the top) training sizes use equal number of cases and controls. The final training size uses all possible remaining controls

**Figure 48:**
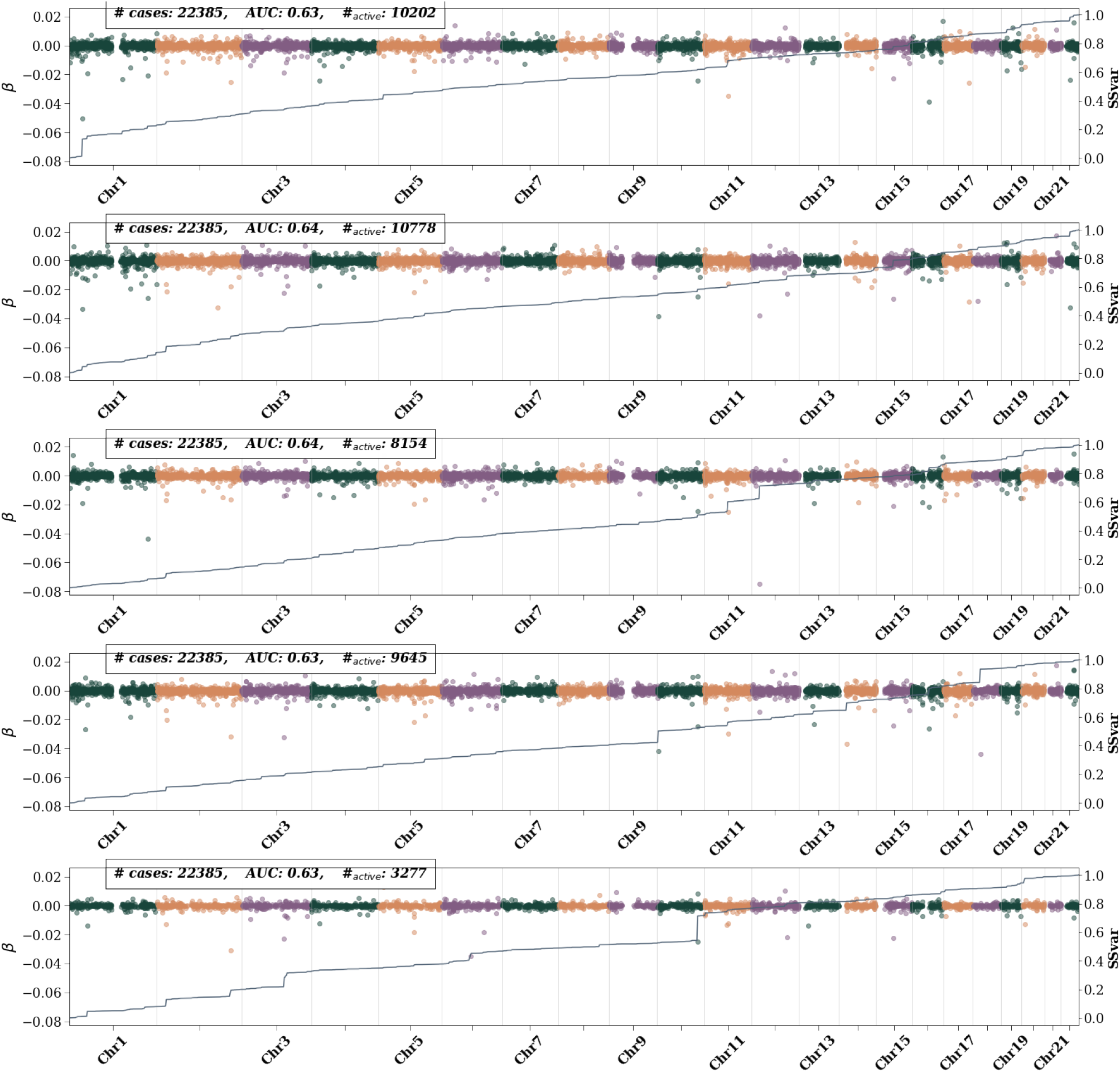
Type 2 diabetes active SNPs – i.e., SNPs with non-zero *β* weights– for 5 CV folds at maximum training size. Left axis shows the *β* value and is represented by colored dots. Different colors are used to differentiate chromosomes. The right axis represents the single SNP variance (SSV) normalized to the total SSV. The “training” label represents the number of cases used in training. All possible controls were used in each fold. While features generally appear consistent across folds, i.e., the presence of a bump in the SSV line, the size of the bump varies.

**Figure 49:**
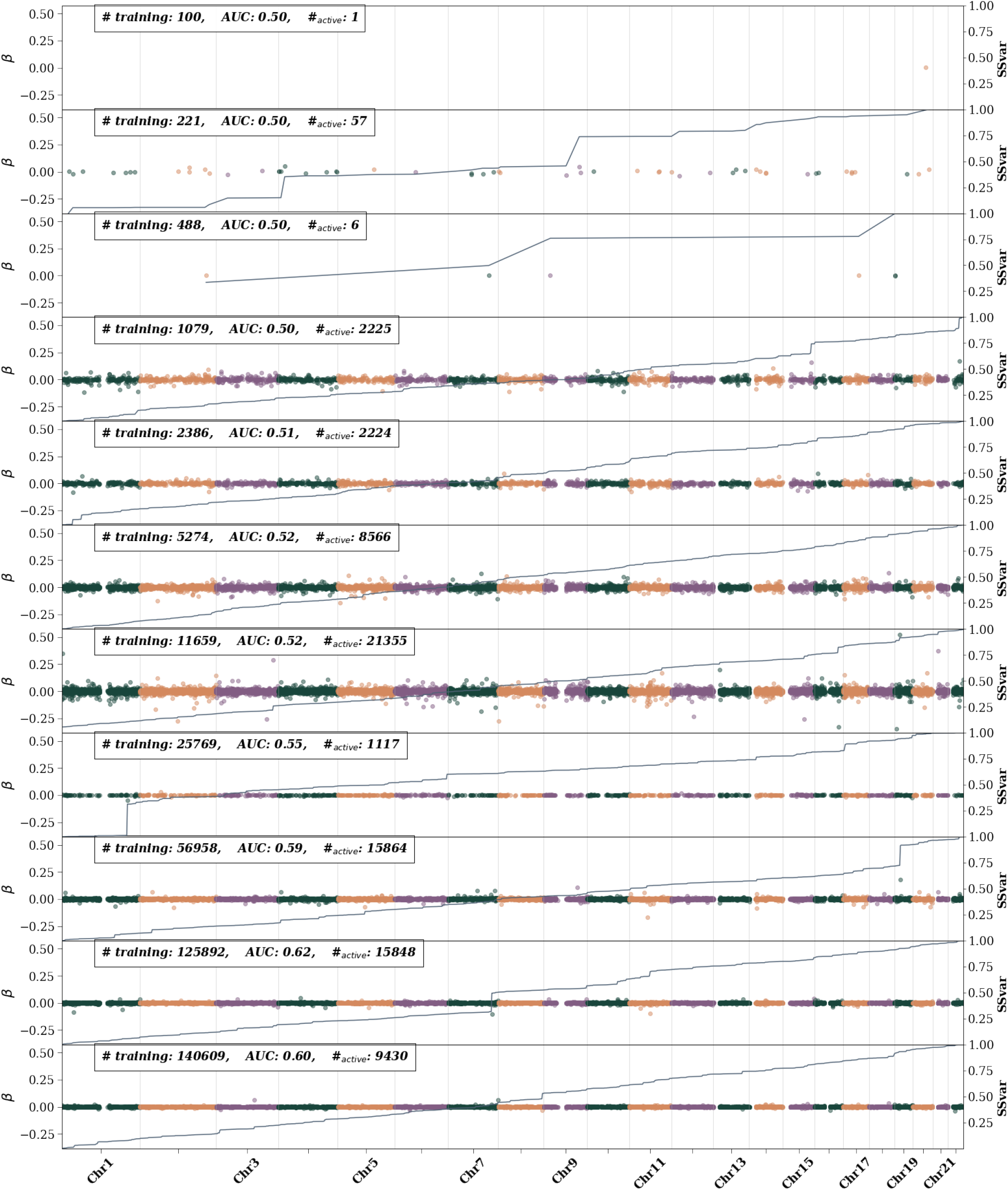
Hypertension active SNPs – i.e., SNPs with non-zero *β* weights– as training size is increased. The left axis shows the *β* value and is represented by colored dots. Different colors are used to differentiate chromosomes. The right axis represents the single SNP variance (SSV) normalized to the total SSV. The solid line showes the cumulative SSV. The “training” label represents the number of cases used in training. The first 10 (from the top) training sizes use equal number of cases and controls. The final training size uses all possible remaining controlse

**Figure 50:**
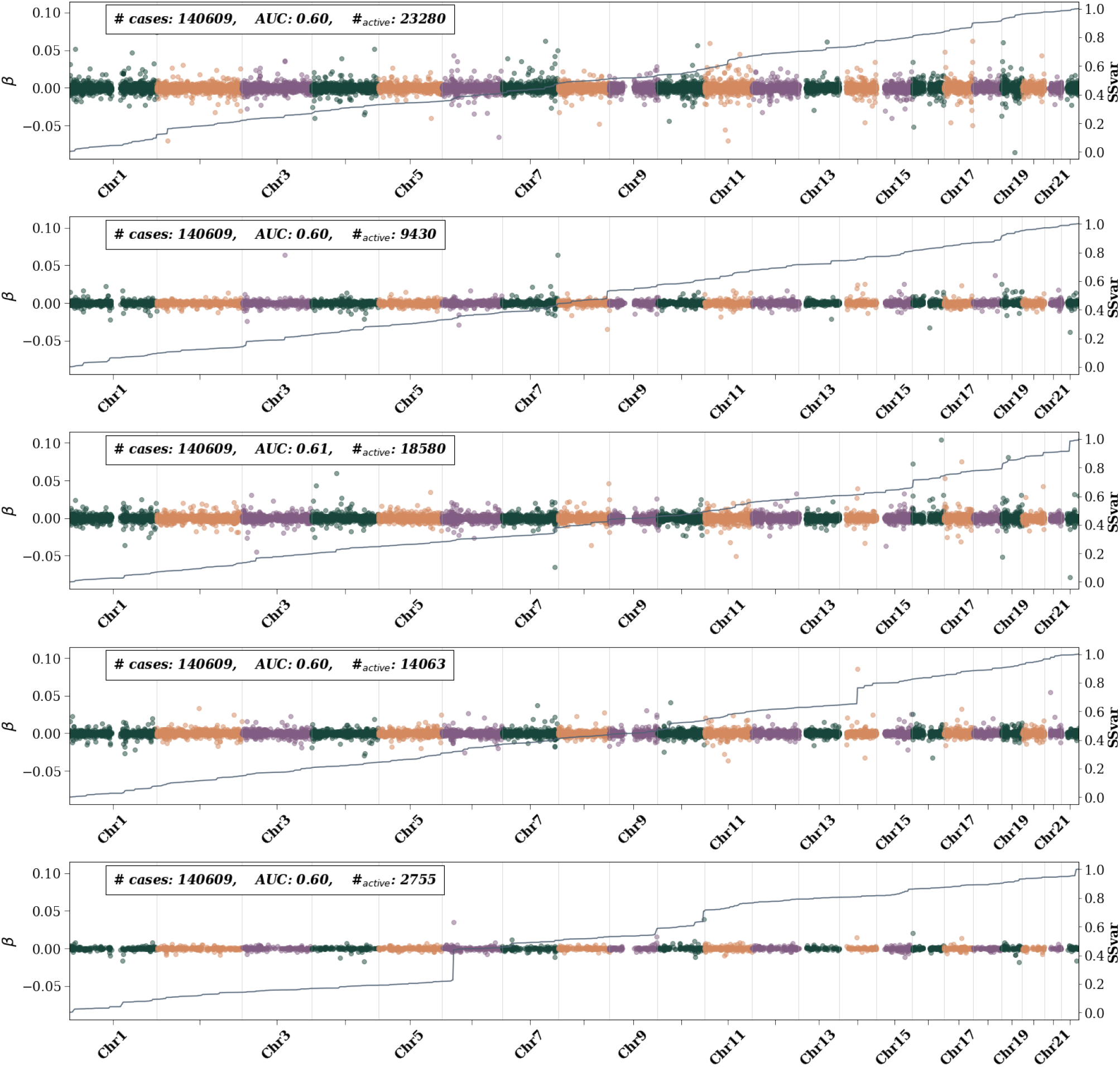
Hypertension active SNPs – i.e., SNPs with non-zero *β* weights– for 5 CV folds at maximum training size. Left axis shows the *β* value and is represented by colored dots. Different colors are used to differentiate chromosomes. The right axis represents the single SNP variance (SSV) normalized to the total SSV. The “training” label represents the number of cases used in training. All possible controls were used in each fold. While features generally appear consistent across folds, i.e., the presence of a bump in the SSV line, the size of the bump varies.

**Figure 51:**
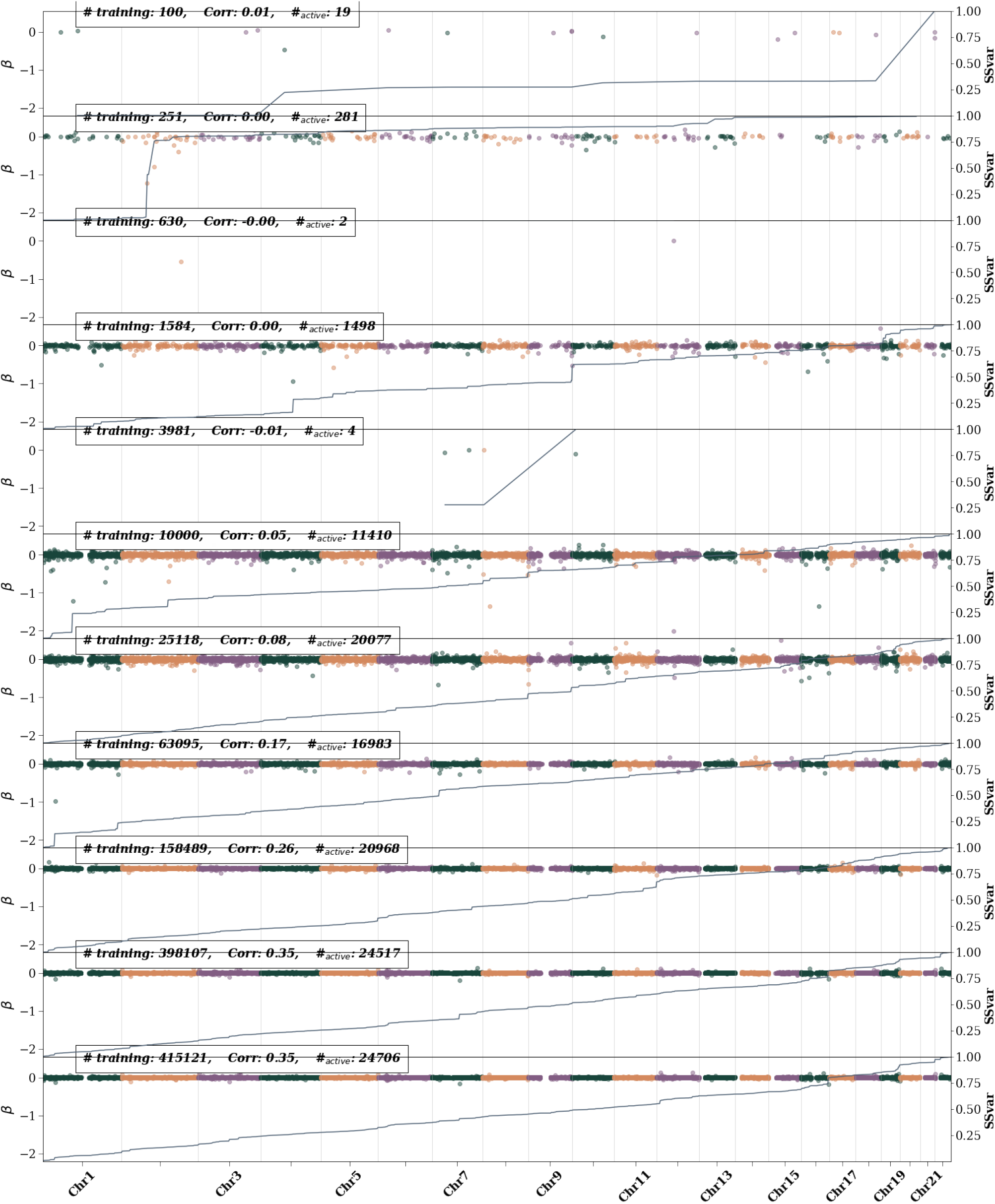
BMI active SNPs – i.e., SNPs with non-zero *β* weights– as training size is increased. The left axis shows the *β* value and is represented by colored dots. Different colors are used to differentiate chromosomes. The right axis represents the single SNP variance (SSV) normalized to the total SSV. The solid line showes the cumulative SSV. The “training” label represents the number of cases used in training. The first 10 (from the top) training sizes use equal number of cases and controls. The final training size uses all possible remaining controlse

**Figure 52:**
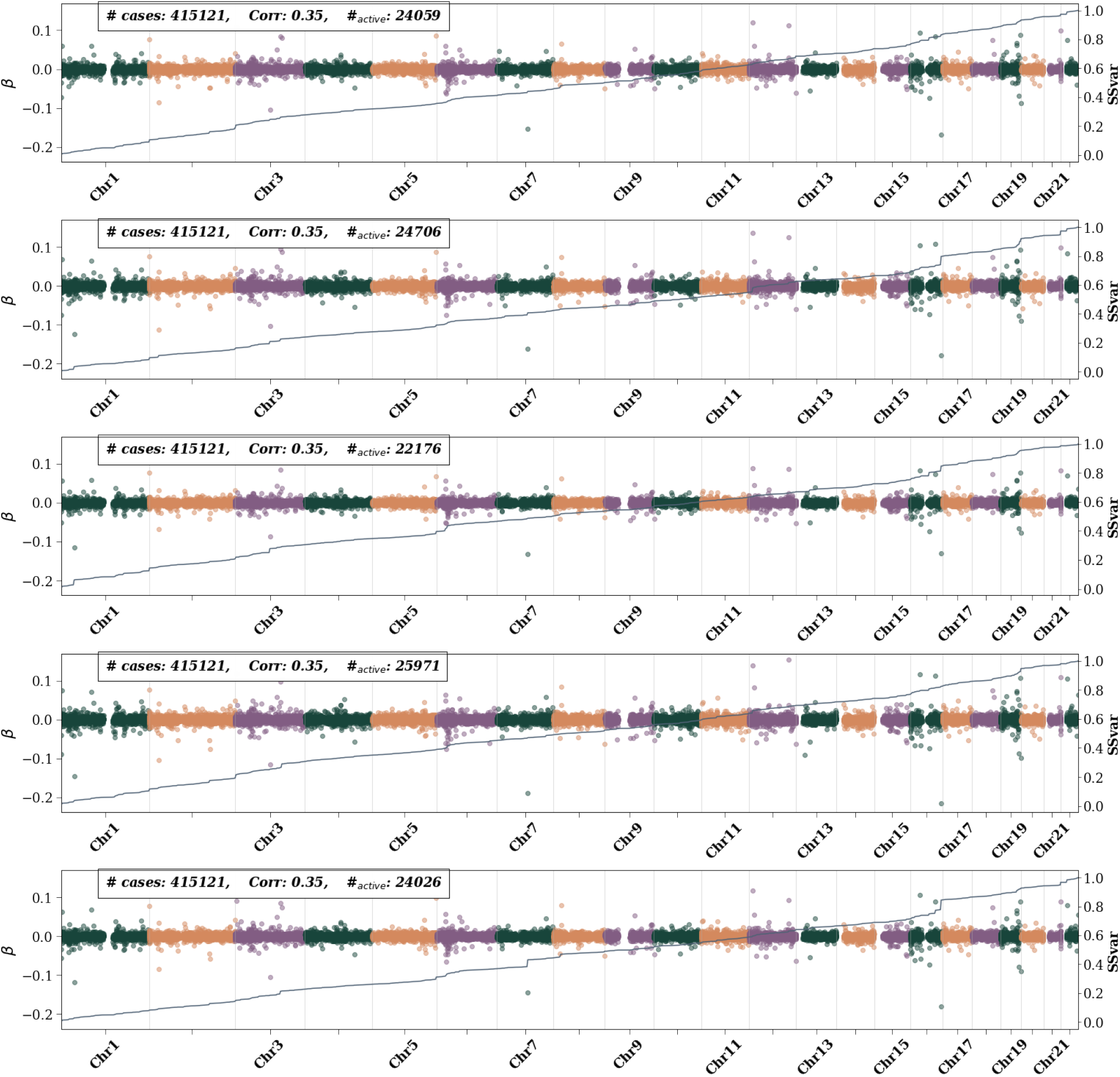
BMI active SNPs – i.e., SNPs with non-zero *β* weights– for 5 CV folds at maximum training size. Left axis shows the *β* value and is represented by colored dots. Different colors are used to differentiate chromosomes. The right axis represents the single SNP variance (SSV) normalized to the total SSV. The “training” label represents the number of cases used in training. All possible controls were used in each fold. While features generally appear consistent across folds, i.e., the presence of a bump in the SSV line, the size of the bump varies.

**Figure 53:**
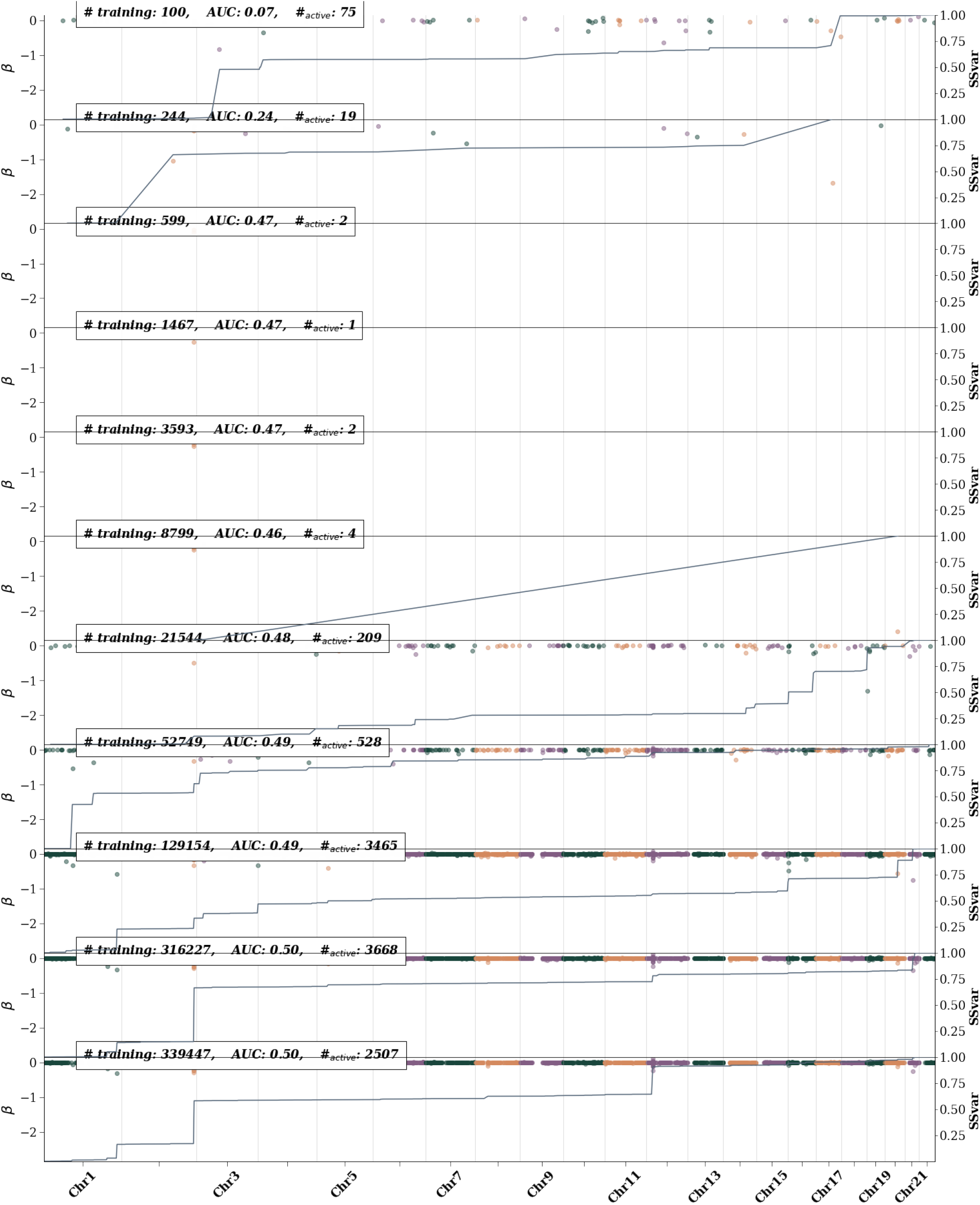
Direct bilirubin active SNPs – i.e., SNPs with non-zero *β* weights– as training size is increased. The left axis shows the *β* value and is represented by colored dots. Different colors are used to differentiate chromosomes. The right axis represents the single SNP variance (SSV) normalized to the total SSV. The solid line showes the cumulative SSV. The “training” label represents the number of cases used in training. The first 10 (from the top) training sizes use equal number of cases and controls. The final training size uses all possible remaining controls

**Figure 54:**
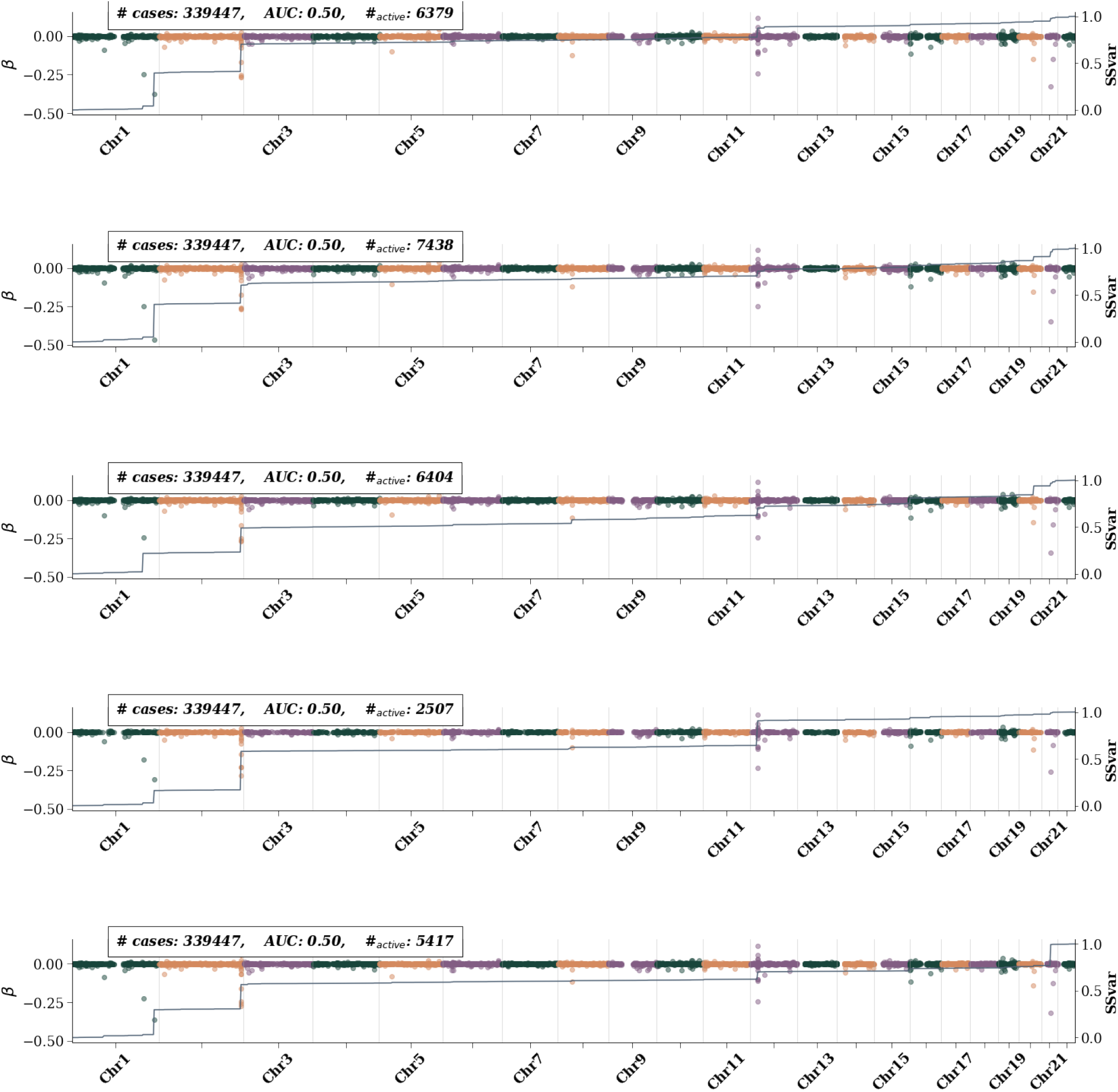
Direct bilirubin active SNPs – i.e., SNPs with non-zero *β* weights– for 5 CV folds at maximum training size. Left axis shows the *β* value and is represented by colored dots. Different colors are used to differentiate chromosomes. The right axis represents the single SNP variance (SSV) normalized to the total SSV. The “training” label represents the number of cases used in training. All possible controls were used in each fold. While features generally appear consistent across folds, i.e., the presence of a bump in the SSV line, the size of the bump varies.

**Figure 55:**
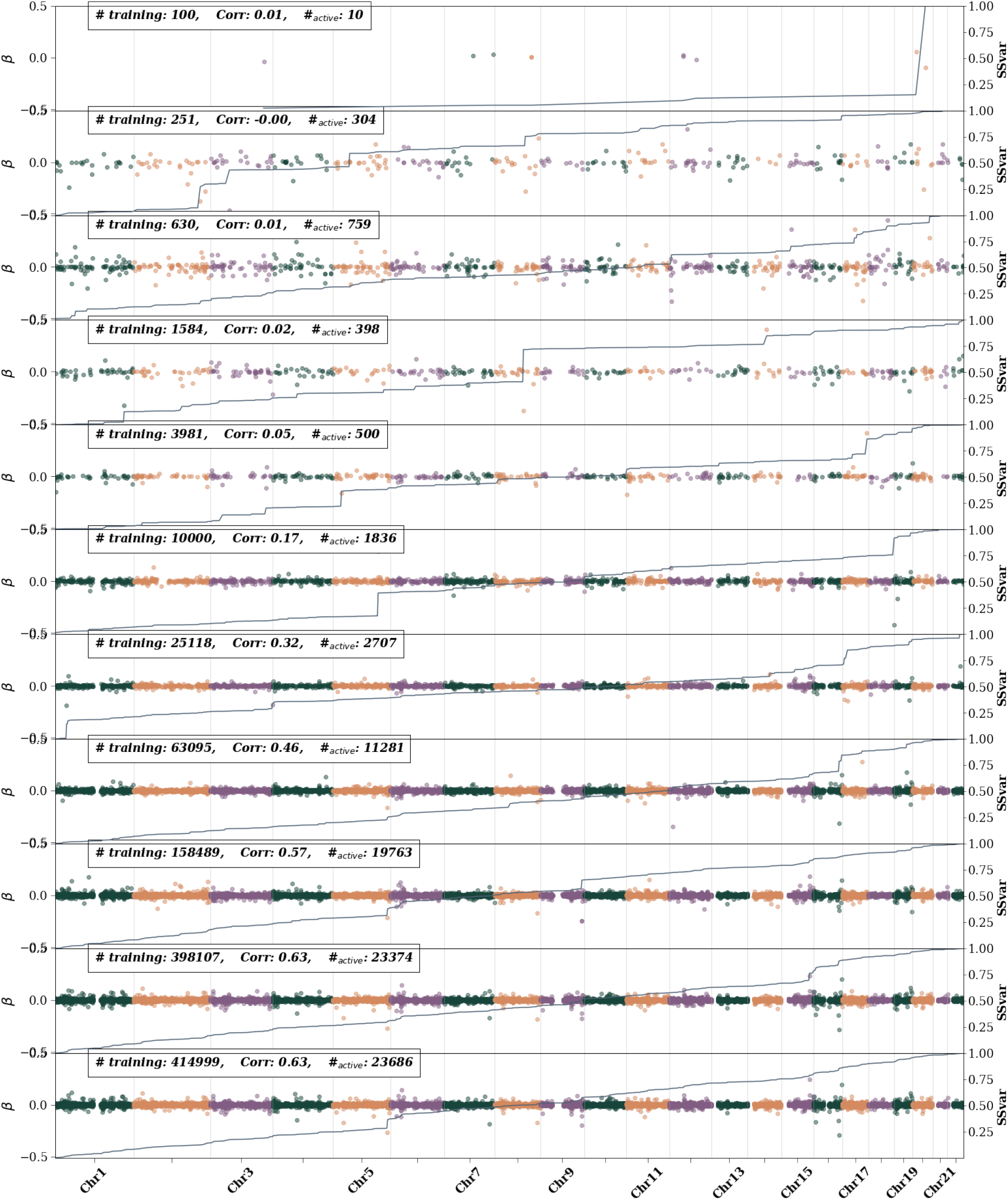
Height active SNPs – i.e., SNPs with non-zero *β* weights– as training size is increased. The left axis shows the *β* value and is represented by colored dots. Different colors are used to differentiate chromosomes. The right axis represents the single SNP variance (SSV) normalized to the total SSV. The solid line showes the cumulative SSV. The “training” label represents the number of cases used in training. The first 10 (from the top) training sizes use equal number of cases and controls. The final training size uses all possible remaining controls

**Figure 56:**
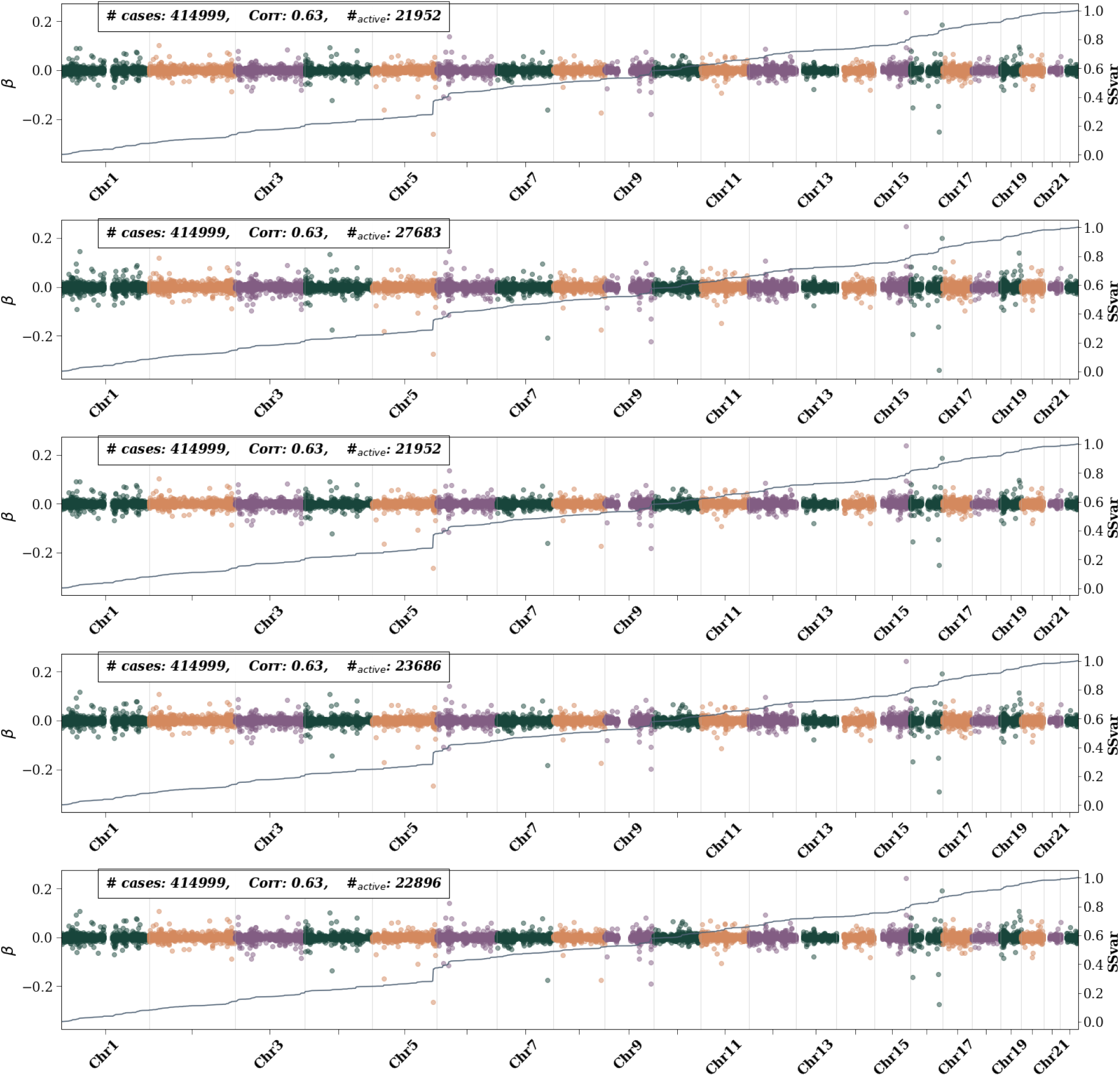
Height active SNPs – i.e., SNPs with non-zero *β* weights– for 5 CV folds at maximum training size. Left axis shows the *β* value and is represented by colored dots. Different colors are used to differentiate chromosomes. The right axis represents the single SNP variance (SSV) normalized to the total SSV. The “training” label represents the number of cases used in training. All possible controls were used in each fold. While features generally appear consistent across folds, i.e., the presence of a bump in the SSV line, the size of the bump varies.

**Figure 57:**
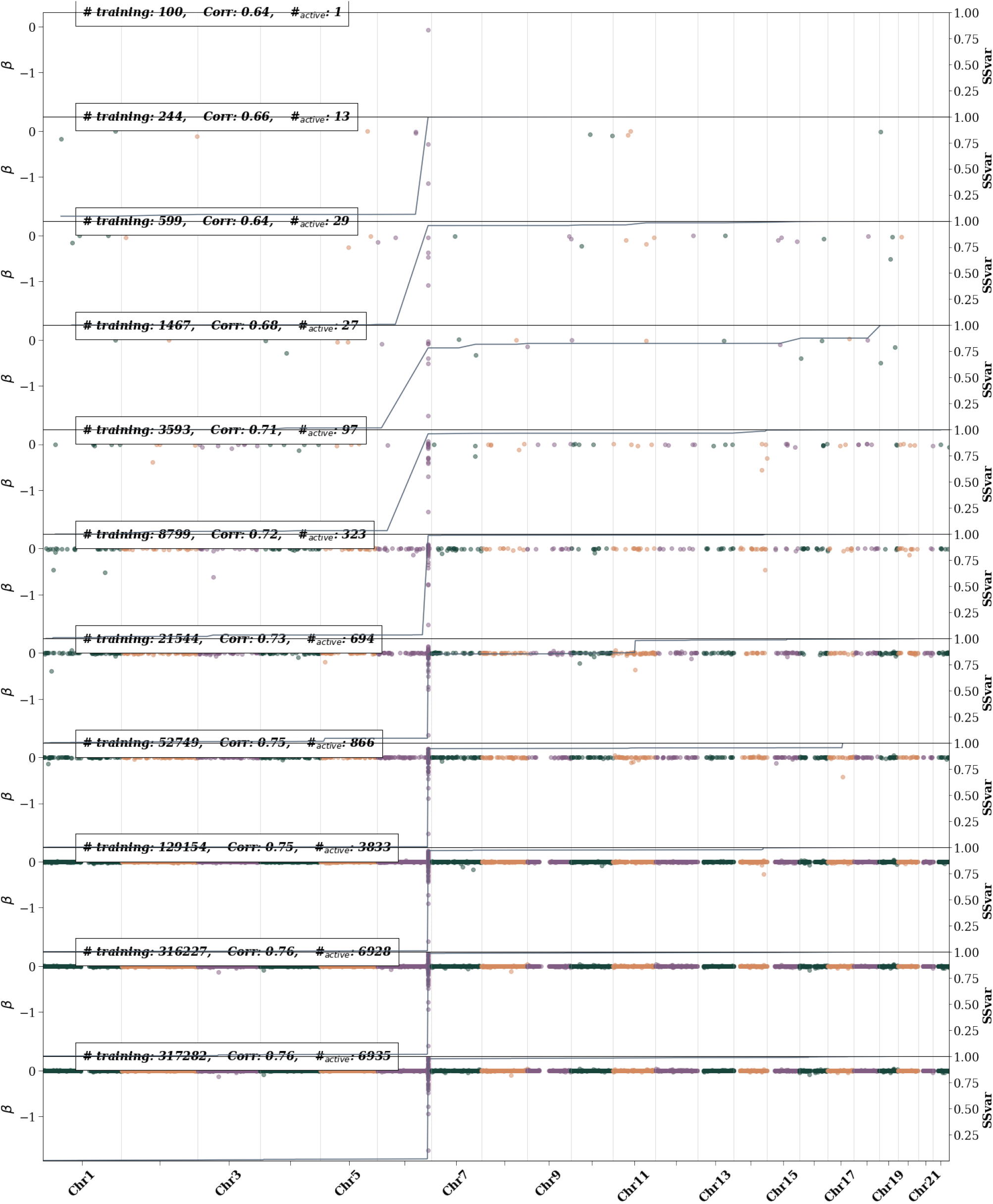
Lipoprotein A active SNPs – i.e., SNPs with non-zero *β* weights– as training size is increased. The left axis shows the *β* value and is represented by colored dots. Different colors are used to differentiate chromosomes. The right axis represents the single SNP variance (SSV) normalized to the total SSV. The solid line showes the cumulative SSV. The “training” label represents the number of cases used in training. The first 10 (from the top) training sizes use equal number of cases and controls. The final training size uses all possible remaining controls

**Figure 58:**
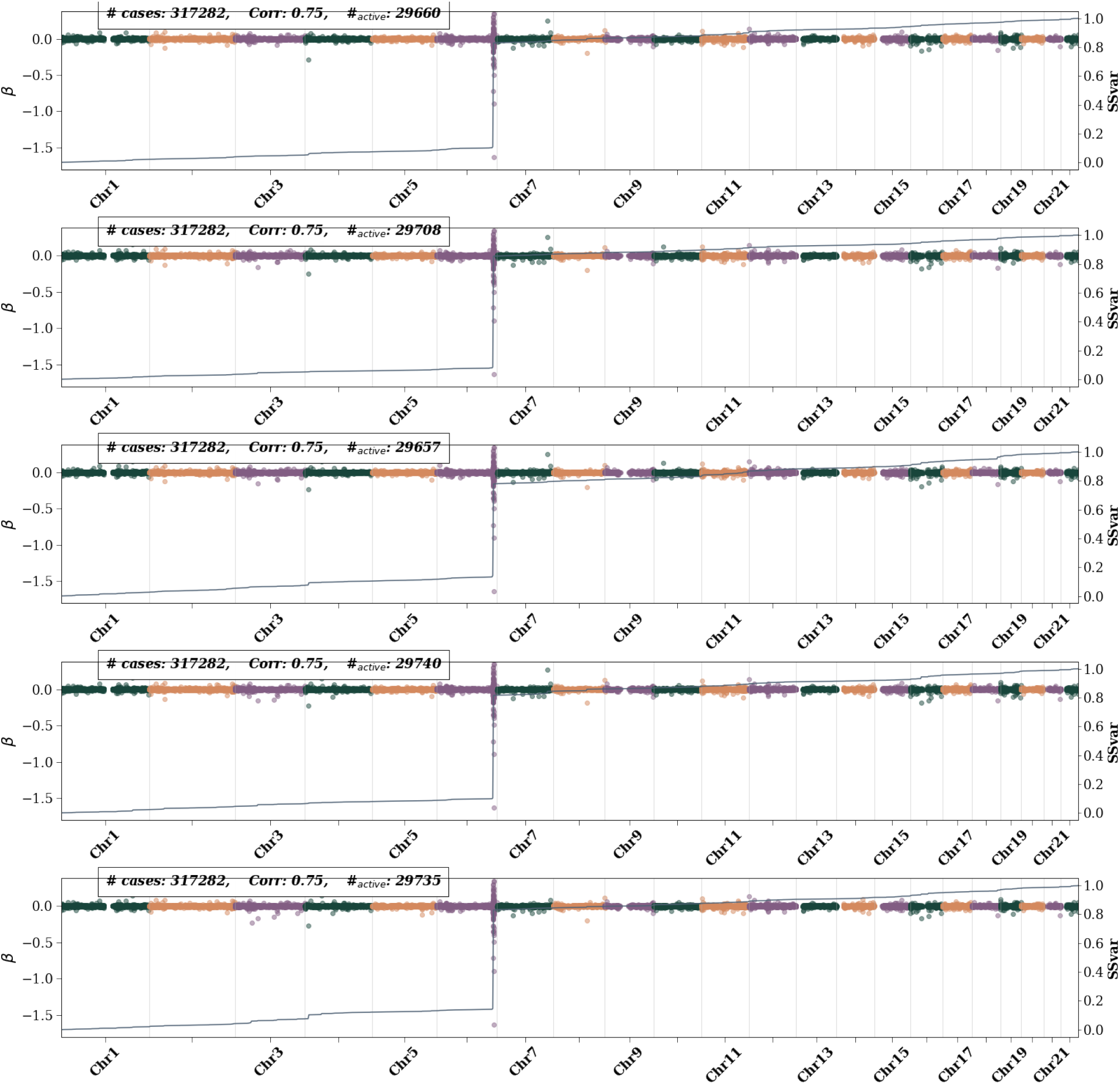
Lipoprotein A active SNPs – i.e., SNPs with non-zero *β* weights– for 5 CV folds at maximum training size. Left axis shows the *β* value and is represented by colored dots. Different colors are used to differentiate chromosomes. The right axis represents the single SNP variance (SSV) normalized to the total SSV. The “training” label represents the number of cases used in training. All possible controls were used in each fold. While features generally appear consistent across folds, i.e., the presence of a bump in the SSV line, the size of the bump varies.

**Figure 59:**
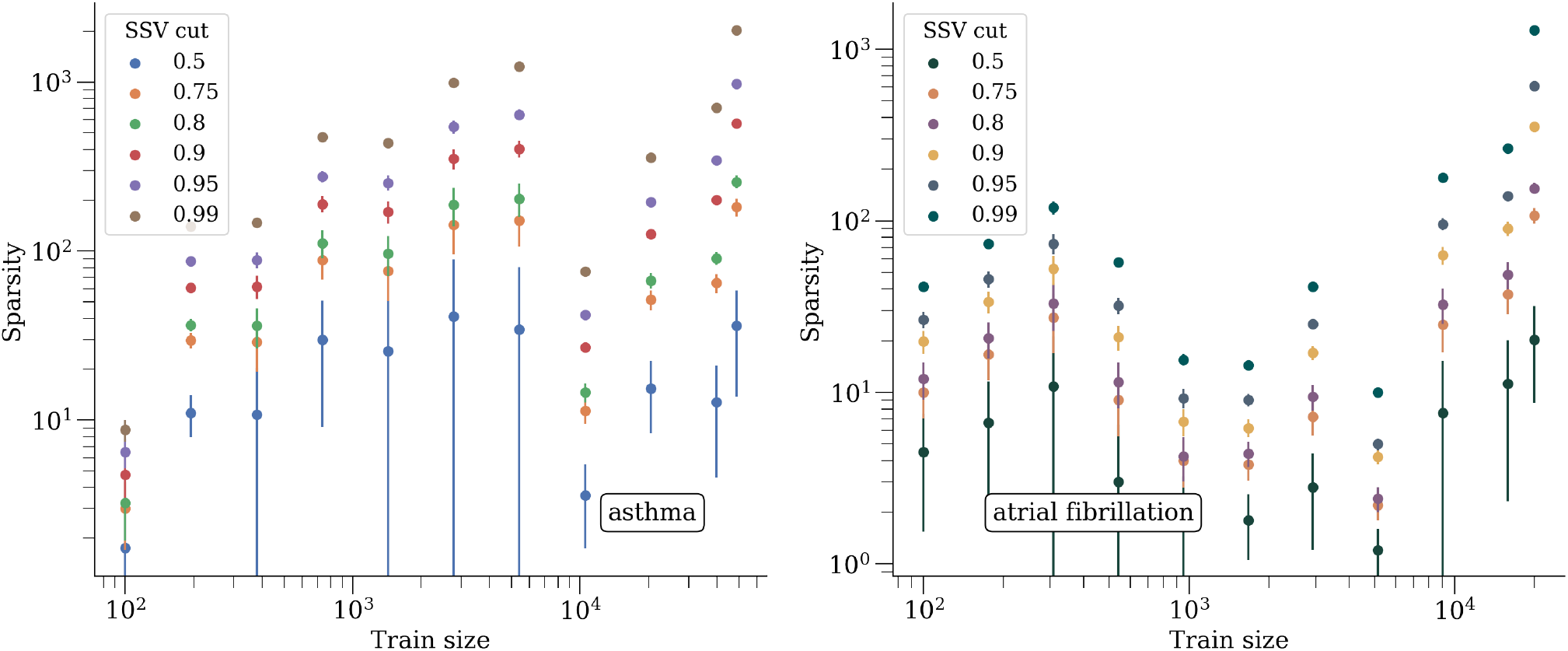
Sparsity as a function of training size, after keeping SNPs that account for 50% - 99% of SSV.

**Figure 60:**
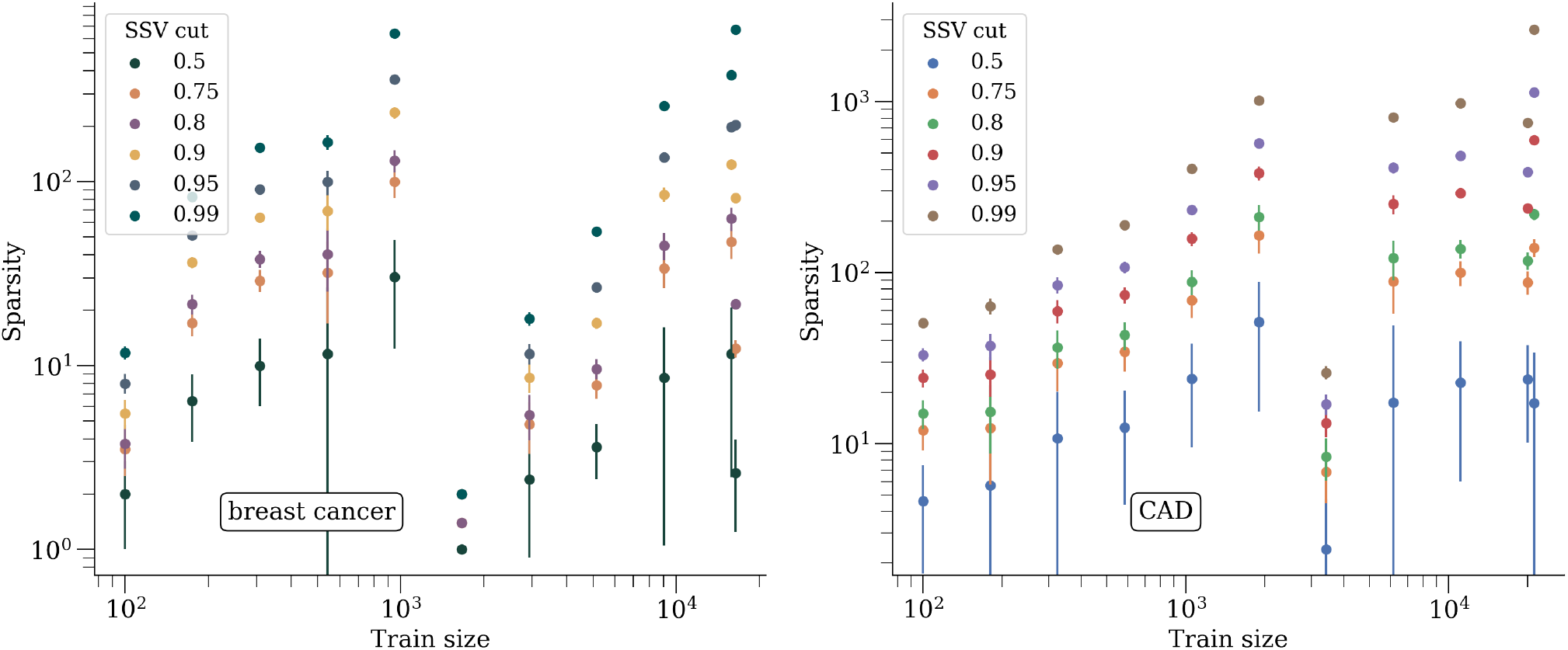
Sparsity as a function of training size, after keeping SNPs that account for 50% - 99% of SSV.

**Figure 61:**
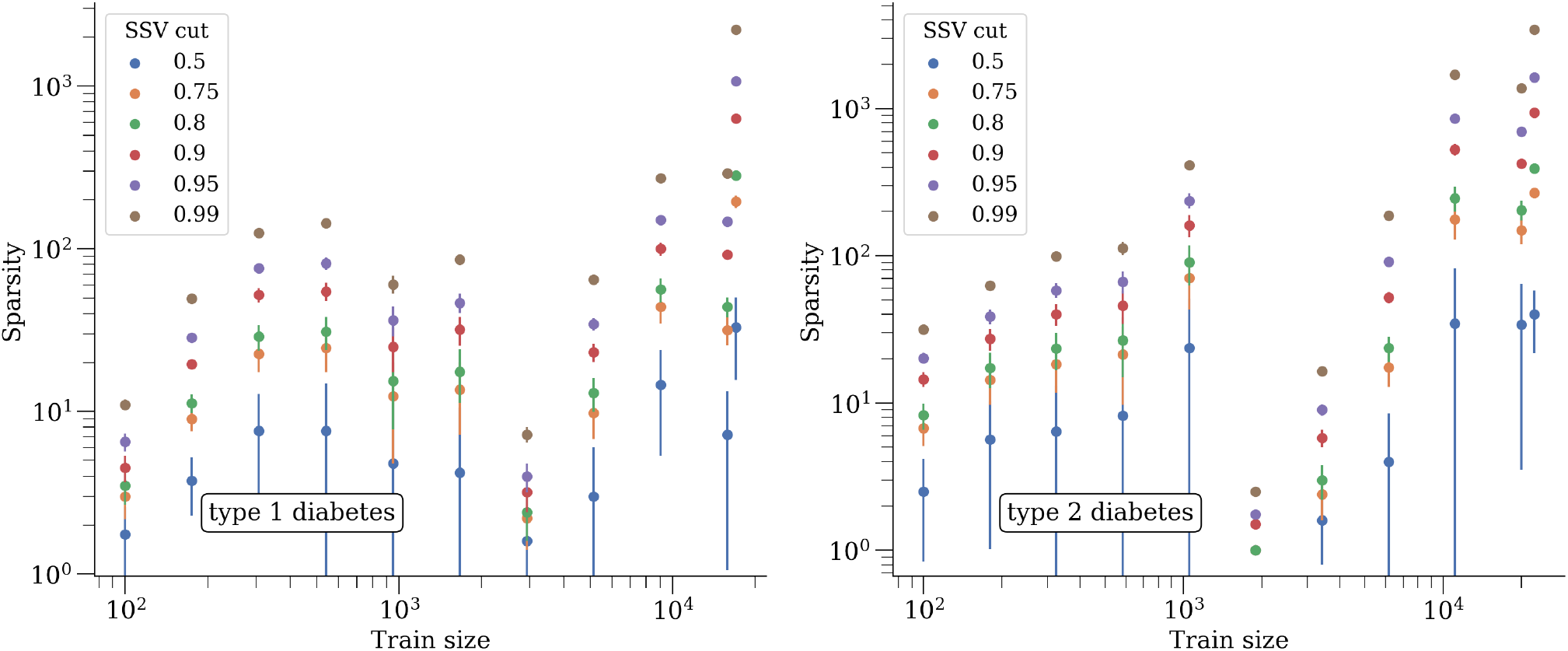
Sparsity as a function of training size, after keeping SNPs that account for 50% - 99% of SSV.

**Figure 62:**
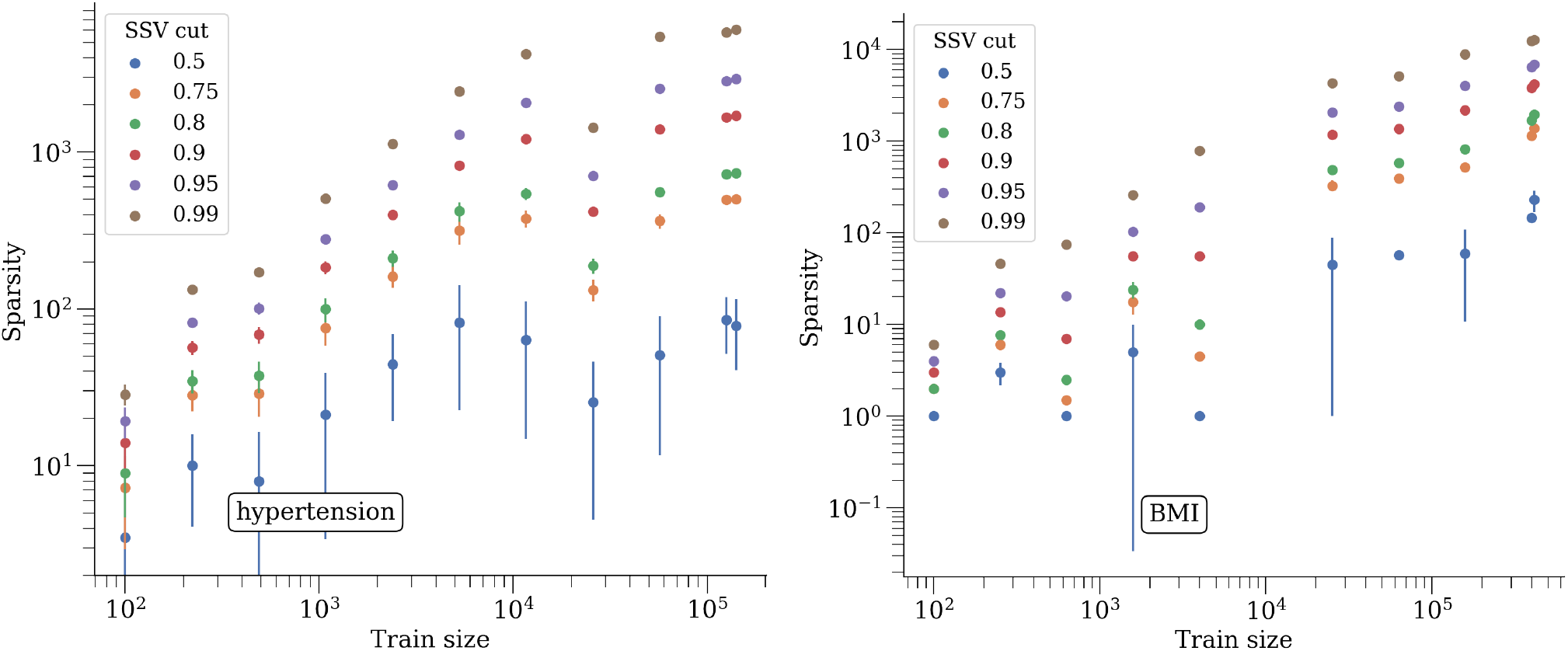
Sparsity as a function of training size, after keeping SNPs that account for 50% - 99% of SSV.

**Figure 63:**
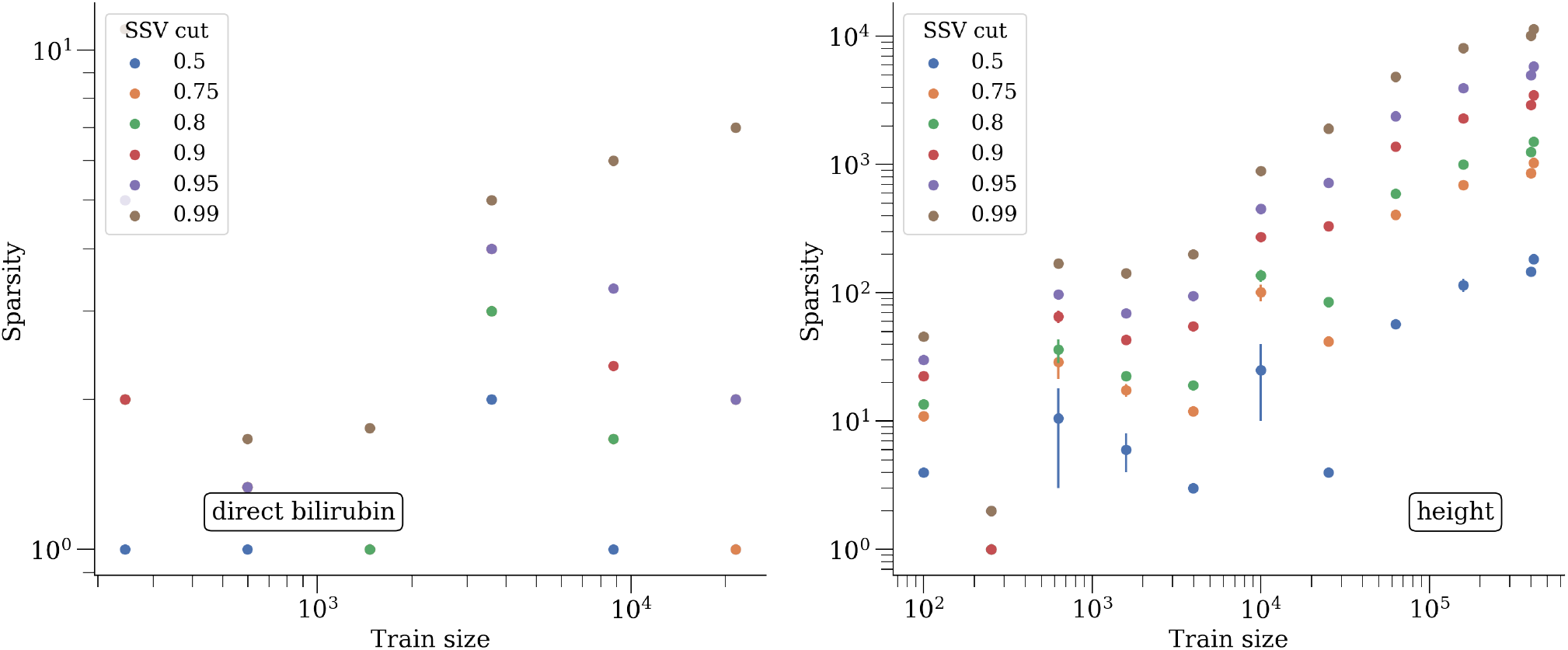
Sparsity as a function of training size, after keeping SNPs that account for 50% - 99% of SSV.

**Figure 64:**
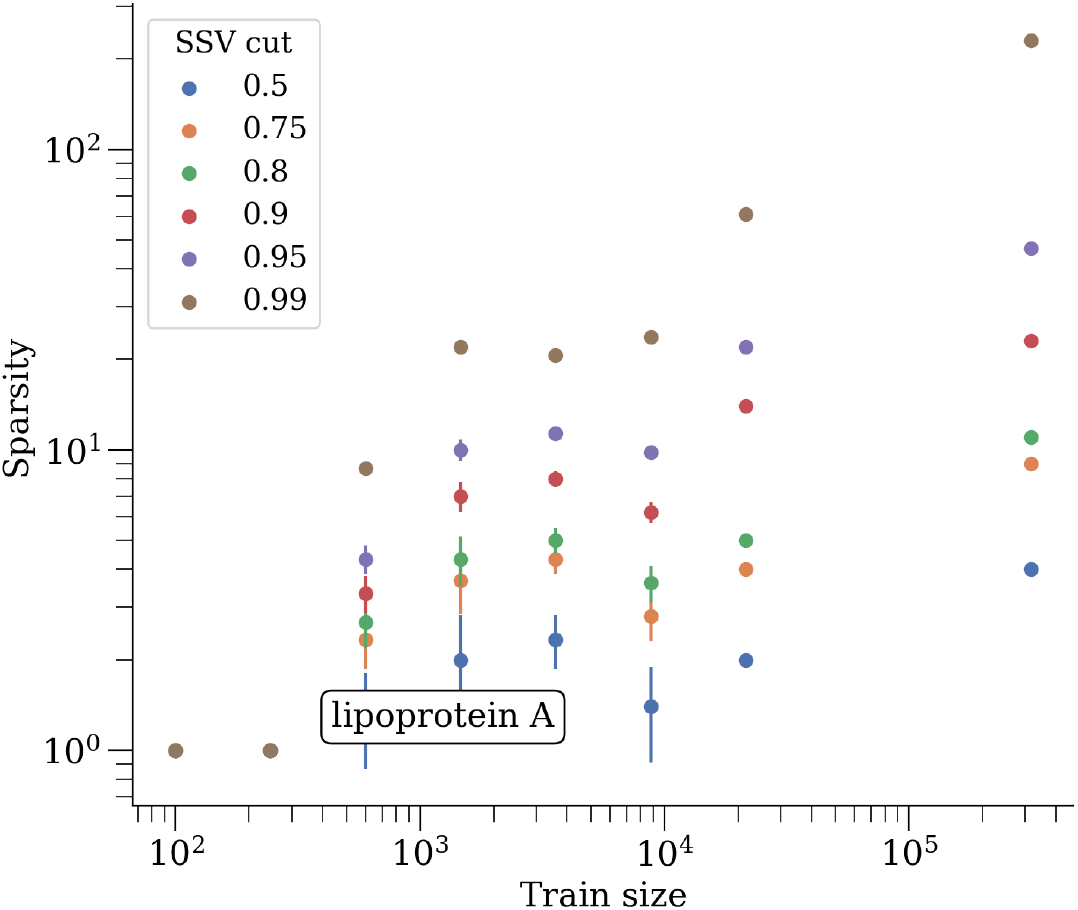
Sparsity as a function of training size, after keeping SNPs that account for 50% - 99% of SSV.

**Figure 65:**
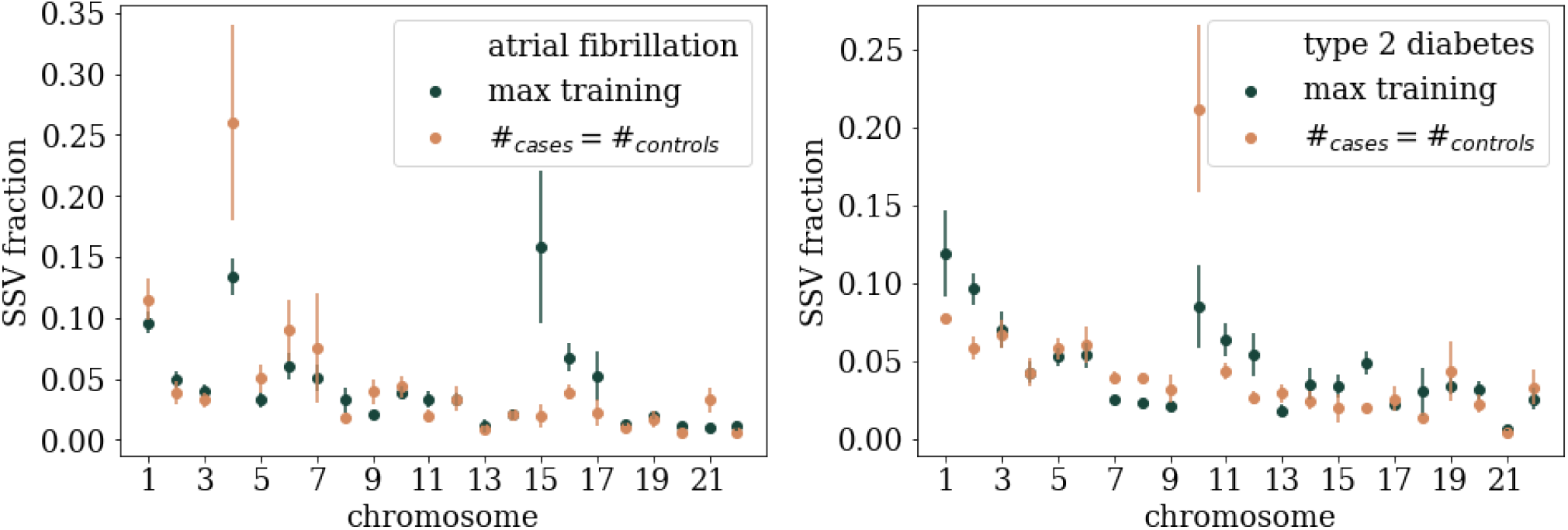
Fraction of SSV per chromosome for max training and near-max number of cases and equal number of controls.

**Figure 66:**
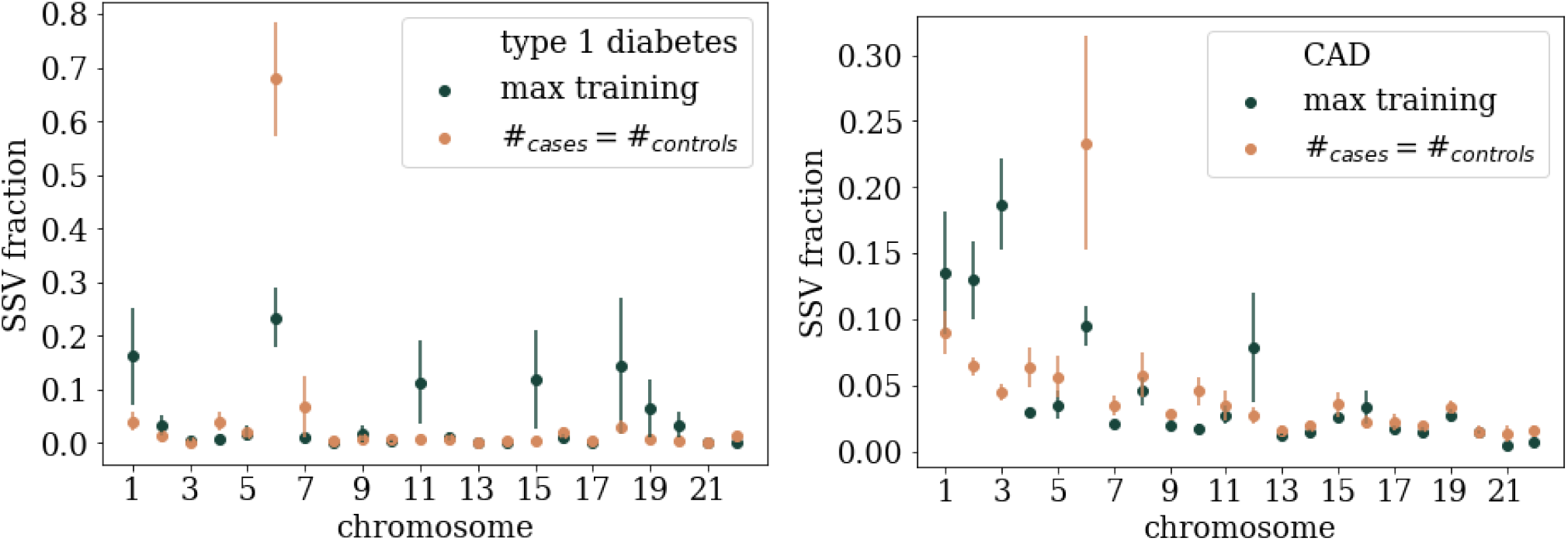
Fraction of SSV per chromosome for max training and near-max number of cases and equal number of controls.

**Figure 67:**
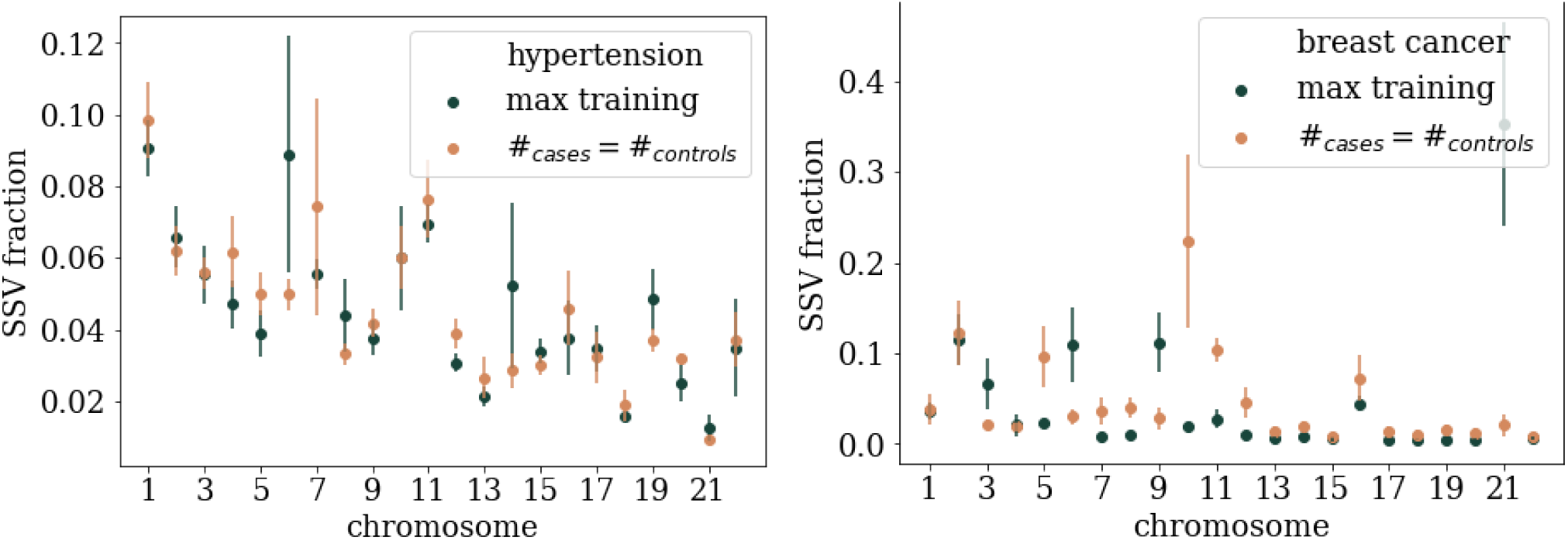
Fraction of SSV per chromosome for max training and near-max number of cases and equal number of controls.

**Figure 68:**
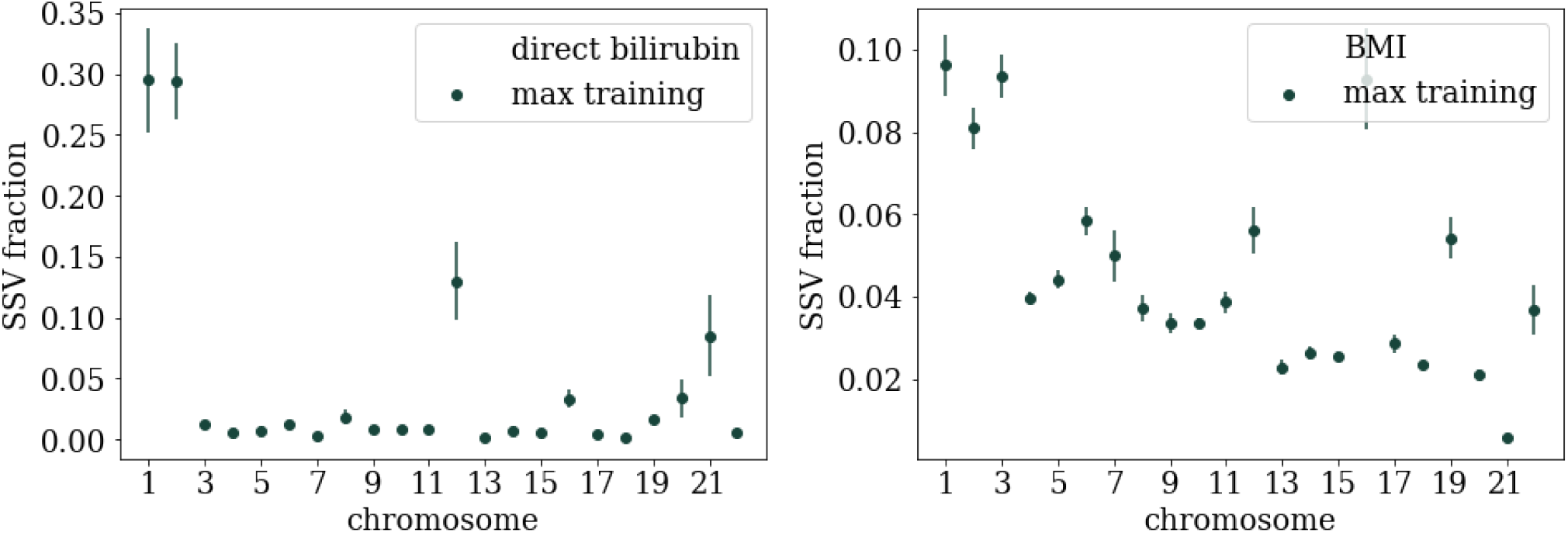
Fraction of SSV per chromosome for max training.

**Figure 69:**
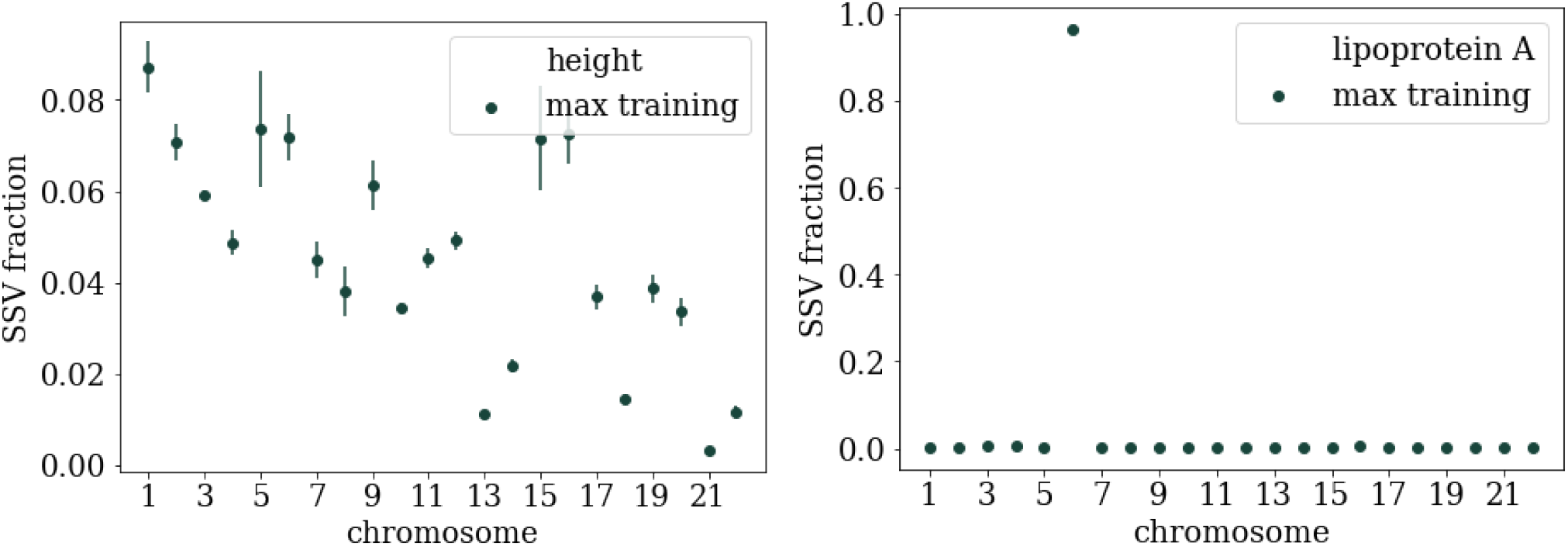
Fraction of SSV per chromosome for max training.

**Figure 70:**
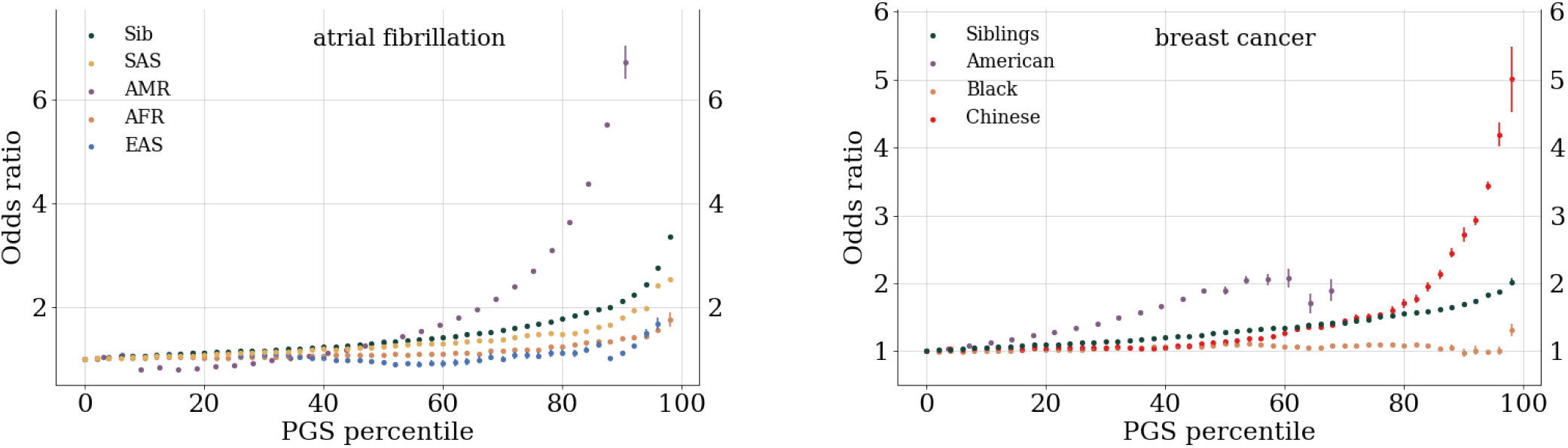
Odds ratio for atrial fibrillation and breast cancer.

## H Odds ratios

**Figure 71:**
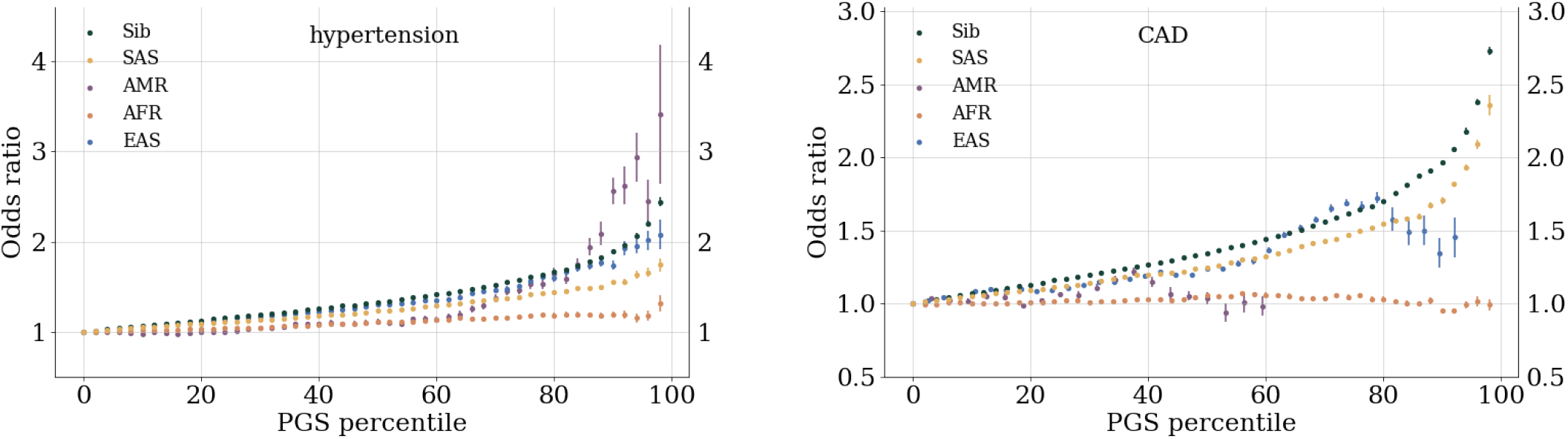
Odds ratio for hypertension and CAD.

**Figure 72:**
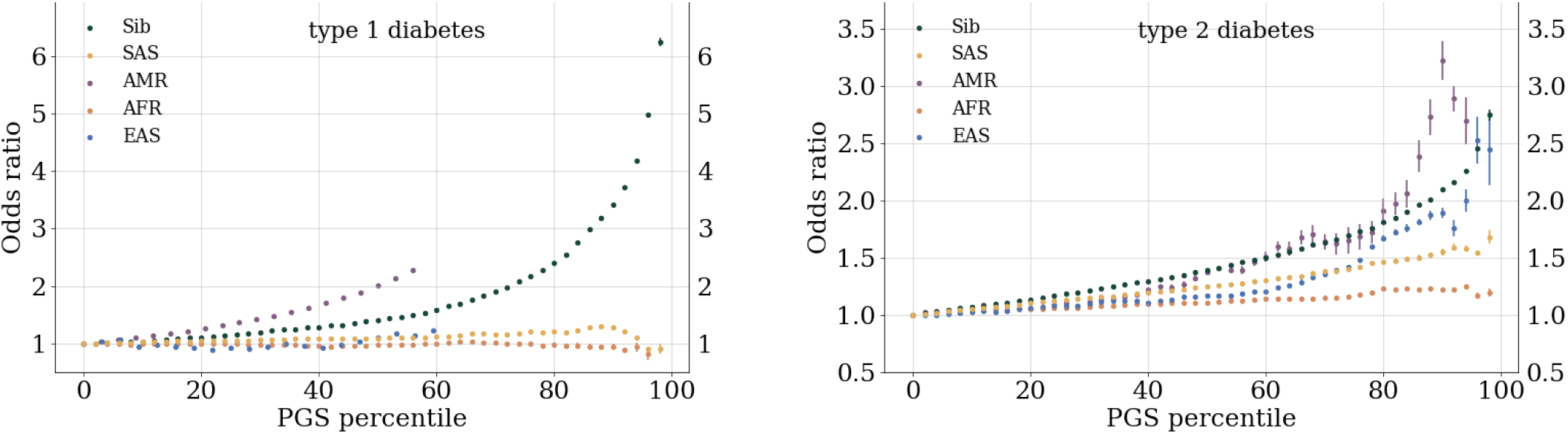
Odds ratio for type 1 and 2 diabetes.

## I Computing details

**Figure 73:**
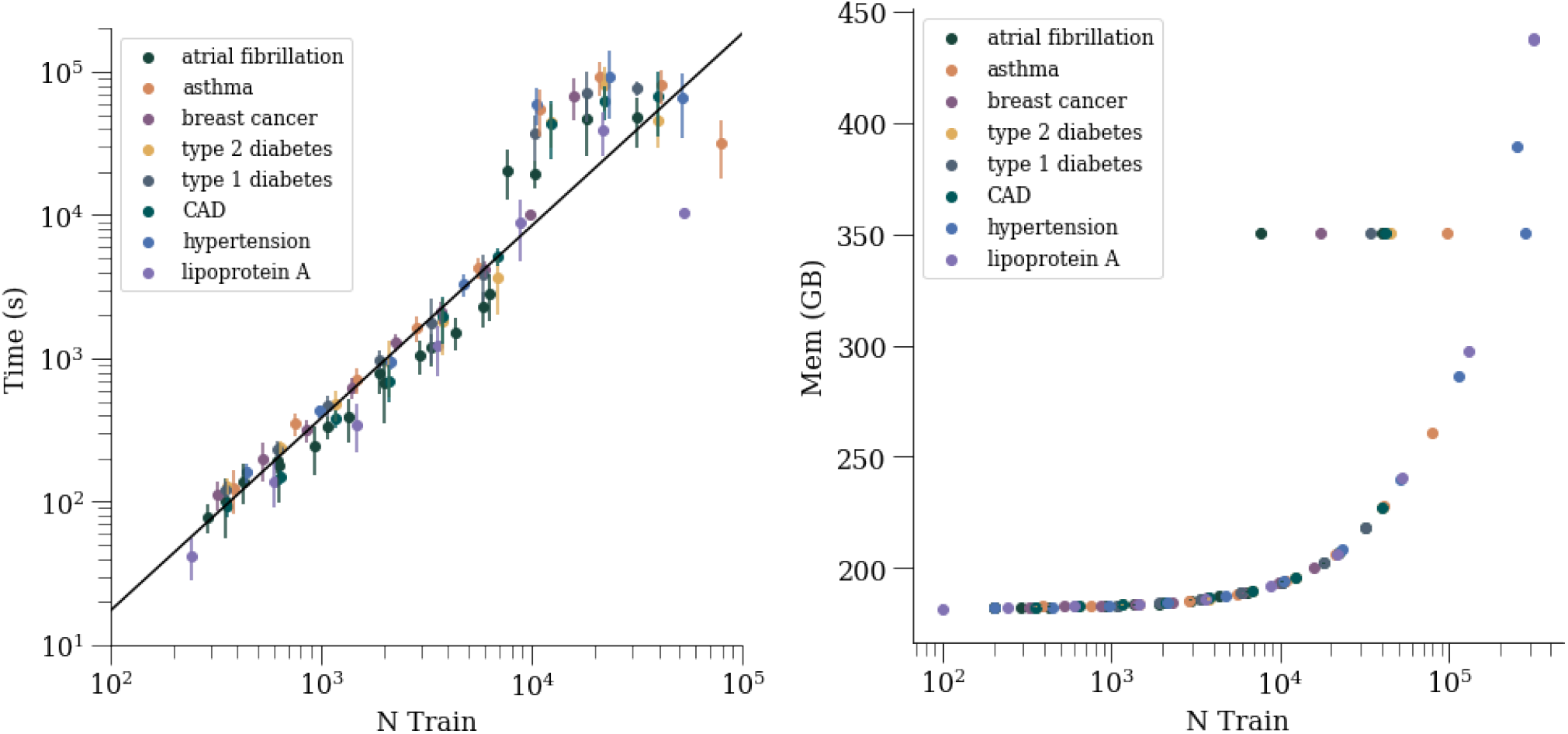
Time and memory usage as a function of training size for LASSO training.

**Figure 74:**
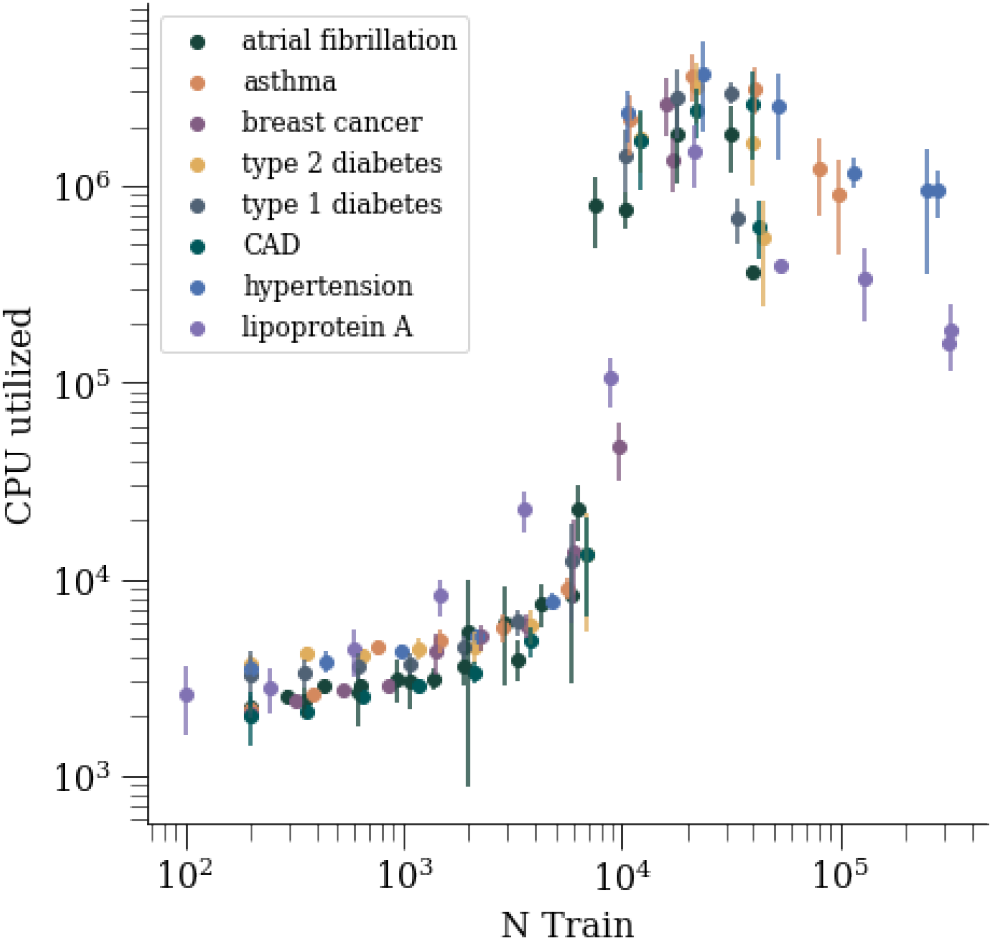
CPU utilization as a function of training size for LASSO training.

## References

1. 1000 Genomes Project Consortium. A map of human genome variation from population-scale sequencing. Nature 467, 1061 (2010) (cit. on pp. 1, 3).

2. TOPMed https://www.nhlbiwgs.org/ (cit. on p. 1).

3. UK Biobank Available online: http://www.ukbiobank.ac.uk/ (accessed: 21-03-2021). http://www.ukbiobank.ac.uk/ (cit. on pp. 1, 2).

4. Taiwan Precision Medicine Initiative https://tpmi.ibms.sinica.edu.tw/www/en/. Accessed: 2023-02-01 (cit. on p. 2).

5. All of Us Research Program Investigators. The “All of Us” research program. New England Journal of Medicine 381, 668–676 (2019) (cit. on p. 2).

6. Martin, A. R., et al. Current clinical use of polygenic scores will risk exacerbating health disparities. bioRxiv. eprint: https://www.biorxiv.org/content/early/2019/02/01/441261.full.pdf. https://www.biorxiv.org/content/early/2019/02/01/441261 (2019) (cit. on p. 2).

7. Duncan, L. et al. Analysis of polygenic risk score usage and performance in diverse human populations. Nature Communications 10, 3328. https://doi.org/10.1038/s41467-019-11112-0 (2019) (cit. on p. 2).

8. Wang, Y., et al. Theoretical and empirical quantification of the accuracy of polygenic scores in ancestry divergent populations. bioRxiv. eprint: https://www.biorxiv.org/content/early/2020/01/15/2020.01.14.905927.full.pdf. https://www.biorxiv.org/content/early/2020/01/15/2020.01.14.905927 (2020) (cit. on p. 2).

9. Martin, A. R. et al. Clinical use of current polygenic risk scores may exacerbate health disparities. Nature Genetics 51, 584–591 (2019) (cit. on p. 2).

10. Lello, L., Raben, T. G., Yong, S. Y., Tellier, L. C. & Hsu, S. D. H. Genomic prediction of 16 complex disease risks including heart attack, diabetes, breast and prostate cancer. Sci Rep 9. [PMC6814833], 1–16 (2019) (cit. on pp. 2, 3, 6, 17).

11. Widen, E., Raben, T. G., Lello, L. & Hsu, S. D. H. Machine Learning Prediction of Biomarkers from SNPs and of Disease Risk from Biomarkers in the UK Biobank. Genes 12. issn: 2073-4425. https://www.mdpi.com/2073-4425/12/7/991 (2021) (cit. on pp. 2, 3).

12. Rosenberg, N. A. et al. Genome-wide association studies in diverse populations. Nature Reviews Genetics 11, 356–366 (2010) (cit. on p. 2).

13. Huang, H., et al. Improving Polygenic Prediction in Ancestrally Diverse Populations (2021) (cit. on p. 2).

14. Cavazos, T. B. & Witte, J. S. Inclusion of Variants Discovered from Diverse Populations Improves Polygenic Risk Score Transferability. bioRxiv. eprint: https://www.biorxiv.org/content/early/2020/05/24/2020.05.21.108845.full.pdf. https://www.biorxiv.org/content/early/2020/05/24/2020.05.21.108845 (2020) (cit. on p. 2).

15. Martin, A. R. et al. Human demographic history impacts genetic risk prediction across diverse populations. The American Journal of Human Genetics 100, 635–649 (2017) (cit. on p. 2).

16. Lewis, C. M. & Vassos, E. Polygenic risk scores: from research tools to clinical instruments. Genome medicine 12, 1–11 (2020) (cit. on p. 2).

17. Lewis, A. C. & Green, R. C. Polygenic risk scores in the clinic: new perspectives needed on familiar ethical issues. Genome Medicine 13, 1–10 (2021) (cit. on p. 2).

18. Bitarello, B. D. & Mathieson, I. Polygenic scores for height in admixed populations. G3: Genes, Genomes, Genetics 10, 4027–4036 (2020) (cit. on p. 2).

19. Atkinson, E. G., et al. Tractor: A framework allowing for improved inclusion of admixed individuals in large-scale association studies. bioRxiv. eprint: https://www.biorxiv.org/content/early/2020/05/19/2020.05.17.100727.full.pdf. https://www.biorxiv.org/content/early/2020/05/19/2020.05.17.100727 (2020) (cit. on p. 2).

20. Cai, M. et al. A unified framework for cross-population trait prediction by leveraging the genetic correlation of polygenic traits. American Journal of Human Genetics 108, 632–655. issn: 15376605. https://doi.org/10.1016/j.ajhg.2021.03.002 (2021) (cit. on p. 2).

21. Weissbrod, O. et al. Functionally informed fine-mapping and polygenic localization of complex trait heritability. Nature Genetics 52. issn: 15461718. http://dx.doi.org/10.1038/s41588-020-00735-5 (2020) (cit. on p. 2).

22. Weissbrod, O. et al. Leveraging fine-mapping and multipopulation training data to improve cross-population polygenic risk scores. Nature Genetics 54, 450–458 (2022) (cit. on p. 2).

23. Veenstra, D. L., Roth, J. A., Garrison Jr, L. P., Ramsey, S. D. & Burke, W. A formal risk-benefit framework for genomic tests: facilitating the appropriate translation of genomics into clinical practice. Genetics in Medicine 12, 686 (2010) (cit. on p. 2).

24. Jacob, H. J. et al. Genomics in clinical practice: lessons from the front lines. Science translational medicine 5, 194cm5–194cm5 (2013) (cit. on p. 2).

25. Euesden, J., Lewis, C. M. & O’reilly, P. F. PRSice: polygenic risk score software. Bioinformatics 31, 1466– 1468 (2014) (cit. on p. 2).

26. Priest, J. R. & Ashley, E. A. Genomics in clinical practice. BMJ Heart 100, 1569–1570 (2014) (cit. on p. 2).

27. Abraham, G. & Inouye, M. Genomic risk prediction of complex human disease and its clinical application. Current Opinion in Genetics & Development 33, 10–16 (2015) (cit. on p. 2).

28. Chatterjee, N., Shi, J. & García-Closas, M. Developing and evaluating polygenic risk prediction models for stratified disease prevention. Nature Reviews Genetics 17, 392 (2016) (cit. on p. 2).

29. Khera, A. V. et al. Genome-wide polygenic scores for common diseases identify individuals with risk equivalent to monogenic mutations. Nature Genetics 50, 1219 (2018) (cit. on pp. 2, 28).

30. Torkamani, A., Wineinger, N. E. & Topol, E. J. The personal and clinical utility of polygenic risk scores. Nature Reviews Genetics 19, 581 (2018) (cit. on p. 2).

31. Liu, L. & Kiryluk, K. Genome-wide polygenic risk predictors for kidney disease. Nature Reviews Nephrology 14, 723–724 (2018) (cit. on p. 2).

32. Khera, A. V. et al. Polygenic prediction of weight and obesity trajectories from birth to adulthood. Cell 177. [PMC6661115], 587–596 (2019) (cit. on p. 2).

33. Nelson, H. D., Pappas, M., Cantor, A., Haney, E. & Holmes, R. Risk assessment, genetic counseling, and genetic testing for BRCA-related cancer in women: updated evidence report and systematic review for the US Preventive Services Task Force. Jama 322, 666–685 (2019) (cit. on p. 2).

34. Amir, E., Freedman, O. C., Seruga, B. & Evans, D. G. Assessing women at high risk of breast cancer: a review of risk assessment models. JNCI: Journal of the National Cancer Institute 102. Oxford University Press, 680–691 (2010) (cit. on p. 2).

35. Choi, S. W., Mak, T. S.-H. & O’Reilly, P. F. Tutorial: a guide to performing polygenic risk score analyses. Nature Protocols 15, 2759–2772 (2020) (cit. on p. 2).

36. Shieh, Y. et al. Breast cancer risk prediction using a clinical risk model and polygenic risk score. Breast Cancer Research and Treatment 159, 513–525 (2016) (cit. on p. 2).

37. Lewis, C. M. & Vassos, E. Prospects for using risk scores in polygenic medicine. Genome Medicine 9, 96. issn: 1756994X (2017) (cit. on p. 2).

38. Bowdin, S. et al. Recommendations for the integration of genomics into clinical practice. Genetics in Medicine 18, 1075 (2016) (cit. on p. 2).

39. Lambert, S. A., Abraham, G. & Inouye, M. Towards clinical utility of polygenic risk scores. Human Molecular Genetics 28, R133–R142. issn: 14602083 (Nov. 2019) (cit. on p. 2).

40. Kuchenbaecker, K. et al. Evaluation of Polygenic Risk Scores for Breast and Ovarian Cancer Risk Prediction in BRCA1 and BRCA2 Mutation Carriers. JNCI: Journal of the National Cancer Institute 109, 7 (2017) (cit. on p. 2).

41. Mavaddat, N. et al. Polygenic Risk Scores for Prediction of Breast Cancer and Breast Cancer Subtypes. The American Journal of Human Genetics 104, 21–34. https://doi.org/10.1016%2Fj.ajhg.2018.11.002 (Jan. 2019) (cit. on p. 2).

42. Hughes, E. et al. Development and Validation of a Clinical Polygenic Risk Score to Predict Breast Cancer Risk. JCO Precision Oncology, 585–592. https://doi.org/10.1200/PO.19.00360 (Aug. 6, 2020) (cit. on p. 2).

43. Fahed, A. C. et al. Polygenic background modifies penetrance of monogenic variants for tier 1 genomic conditions. Nature Communications 11, 1–9 (2020) (cit. on p. 2).

44. Llewellyn, C. H., Trzaskowski, M, Plomin, R & Wardle, J. Finding the missing heritability in pediatric obesity: the contribution of genome-wide complex trait analysis. International Journal of Obesity 37, 1506–1509. https://doi.org/10.1038/ijo.2013.30 (Mar. 2013) (cit. on p. 2).

45. Maher, B. Personal genomes: The case of the missing heritability. Nature 456, 18–21. https://doi.org/10.1038/456018a (Nov. 2008) (cit. on p. 2).

46. Makowsky, R. et al. Beyond Missing Heritability: Prediction of Complex Traits. PLoS Genetics 7 (ed Gibson, G.) e1002051. https://doi.org/10.1371/journal.pgen.1002051 (Apr. 2011) (cit. on p. 2).

47. Vattikuti, S., Guo, J. & Chow, C. C. Heritability and Genetic Correlations Explained by Common SNPs for Metabolic Syndrome Traits. PLoS Genetics 8 (ed Visscher, P. M.) e1002637. https://doi.org/10.1371/journal.pgen.1002637 (Mar. 2012) (cit. on p. 2).

48. De los Campos, G., Sorensen, D. & Gianola, D. Genomic Heritability: What Is It? PLOS Genetics 11 (ed Barsh, G. S.) e1005048. https://doi.org/10.1371/journal.pgen.1005048 (May 2015) (cit. on p. 2).

49. Kim, H., Grueneberg, A., Vazquez, A. I., Hsu, S. & de los Campos, G. Will Big Data Close the Missing Heritability Gap? Genetics 207. [PMC5676235], 1135–1145. https://doi.org/10.1534/genetics.117.300271 (Sept. 2017) (cit. on p. 2).

50. Lee, J. J. & Chow, C. C. Conditions for the validity of SNP-based heritability estimation. Human Genetics 133, 1011–1022 (2014) (cit. on p. 2).

51. Lello, L. et al. Accurate genomic prediction of human height. Genetics 210. [PMC6216598], 477–497 (2018) (cit. on pp. 2, 17).

52. Privé, F. et al. Portability of 245 polygenic scores when derived from the UK Biobank and applied to 9 ancestry groups from the same cohort. American Journal of Human Genetics 109, 12–23. issn: 15376605 (2022) (cit. on pp. 2, 4).

53. Privé, F., Arbel, J. & Vilhjálmsson, B. J. LDpred2: better, faster, stronger. Bioinformatics 36, 5424–5431 (2020) (cit. on p. 2).

54. Wand, H. et al. Improving reporting standards for polygenic scores in risk prediction studies. Nature 591, 211–219. https://doi.org/10.1038/s41586-021-03243-6 (2021) (cit. on p. 3).

55. Martin, A. R. et al. Clinical use of current polygenic risk scores may exacerbate health disparities. Nature genetics 51. PMC6563838, 584 (2019) (cit. on pp. 3, 4, 14).

56. Lello, L., Raben, T. G. & Hsu, S. D. H. Sibling validation of polygenic risk scores and complex trait prediction. Scientific Reports 10. [PMC7411027], 13190. https://doi.org/10.1038/s41598-020-69927-7 (2020) (cit. on pp. 3, 5, 17).

57. Lello, L., Hsu, M., Widen, E. & Raben, T. G. Sibling Variation in Phenotype and Genotype: Polygenic Trait Distributions and DNA Recombination Mapping with UK Biobank and IVF Family Data. medRxiv (2022) (cit. on pp. 3, 5, 16).

58. Kong, A. et al. The nature of nurture: Effects of parental genotypes. Science 359, 424–428 (2018) (cit. on p. 5).

59. Vattikuti, S., Lee, J. J., Chang, C. C., Hsu, S. D. H. & Chow, C. C. Applying compressed sensing to genome-wide association studies. GigaScience 3. [PMC4078394], 10. issn: 2047-217X. http://dx.doi.org/10.1186/2047-217X-3-10 (2014) (cit. on pp. 7, 9, 17).

60. Park, L. Population-specific long-range linkage disequilibrium in the human genome and its influence on identifying common disease variants. Scientific Reports 9, 1–13 (2019) (cit. on p. 8).

61. Ober, C. & Nicolae, D. L. Meta-analysis of genome-wide association studies of asthma in ethnically diverse North American populations. Nature genetics 43, 887–892 (2011) (cit. on p. 8).

62. Moffatt, M. F. et al. A large-scale, consortium-based genomewide association study of asthma. New England Journal of Medicine 363, 1211–1221 (2010) (cit. on p. 8).

63. Ferreira, M. A. et al. Genome-wide association analysis identifies 11 risk variants associated with the asthma with hay fever phenotype. Journal of Allergy and Clinical Immunology 133, 1564–1571 (2014) (cit. on p. 8).

64. Smith, D. et al. A rare IL33 loss-of-function mutation reduces blood eosinophil counts and protects from asthma. PLoS genetics 13, e1006659 (2017) (cit. on p. 8).

65. Pividori, M., Schoettler, N., Nicolae, D. L., Ober, C. & Im, H. K. Shared and distinct genetic risk factors for childhood-onset and adult-onset asthma: genome-wide and transcriptome-wide studies. The Lancet Respiratory Medicine 7, 509–522 (2019) (cit. on p. 8).

66. Feghaly, J., Zakka, P., London, B., MacRae, C. A. & Refaat, M. M. Genetics of atrial fibrillation. Journal of the American Heart Association 7, e009884 (2018) (cit. on p. 8).

67. Thorolfsdottir, R. B. et al. Coding variants in RPL3L and MYZAP increase risk of atrial fibrillation. Communications biology 1, 1–9 (2018) (cit. on p. 8).

68. Ling, T.-Y. et al. F-box protein-32 down-regulates small-conductance calcium-activated potassium channel 2 in diabetic mouse atria. Journal of Biological Chemistry 294, 4160–4168 (2019) (cit. on p. 8).

69. Roselli, C., Rienstra, M. & Ellinor, P. T. Genetics of atrial fibrillation in 2020: GWAS, genome sequencing, polygenic risk, and beyond. Circulation research 127, 21–33 (2020) (cit. on p. 8).

70. Shiovitz, S. & Korde, L. A. Genetics of breast cancer: a topic in evolution. Annals of Oncology 26, 1291–1299 (2015) (cit. on p. 8).

71. Mambiya, M. et al. The play of genes and non-genetic factors on type 2 diabetes. Frontiers in public health 7, 349 (2019) (cit. on p. 8).

72. Nyaga, D. M., Vickers, M. H., Jefferies, C., Perry, J. K. & O’Sullivan, J. M. Type 1 diabetes mellitus-associated genetic variants contribute to overlapping immune regulatory networks. Frontiers in genetics 9, 535 (2018) (cit. on p. 8).

73. McPherson, R. & Tybjaerg-Hansen, A. Genetics of coronary artery disease. Circulation research 118, 564–578 (2016) (cit. on p. 8).

74. Khera, A. V. & Kathiresan, S. Genetics of coronary artery disease: discovery, biology and clinical translation. Nature Reviews Genetics 18, 331–344 (2017) (cit. on p. 8).

75. Ehret, G. B. & Caulfield, M. J. Genes for blood pressure: an opportunity to understand hypertension. European heart journal 34, 951–961 (2013) (cit. on p. 8).

76. Lin, J.-P., Vitek, L. & Schwertner, H. A. Serum bilirubin and genes controlling bilirubin concentrations as biomarkers for cardiovascular disease. Clinical chemistry 56, 1535–1543 (2010) (cit. on p. 8).

77. Chiddarwar, A. S., D’Silva, S. Z., Colah, R. B., Ghosh, K. & Mukherjee, M. B. Genetic variations in bilirubin metabolism genes and their association with unconjugated hyperbilirubinemia in adults. Annals of Human Genetics 81, 11–19 (2017) (cit. on p. 8).

78. Choquet, H. & Meyre, D. Genetics of obesity: what have we learned? Current genomics 12, 169–179 (2011) (cit. on p. 8).

79. Lui, J. C. et al. Synthesizing genome-wide association studies and expression microarray reveals novel genes that act in the human growth plate to modulate height. Human molecular genetics 21, 5193–5201 (2012) (cit. on p. 8).

80. Yengo, L., et al. A Saturated Map of Common Genetic Variants Associated with Human Height from 5.4 Million Individuals of Diverse Ancestries. bioRxiv (2022) (cit. on p. 8).

81. Ronald, J. et al. Genetic variation in LPAL2, LPA, and PLG predicts plasma lipoprotein (a) level and carotid artery disease risk. Stroke 42, 2–9 (2011) (cit. on p. 8).

82. Paquette, M., Bernard, S. & Baass, A. SLC22A3 is associated with lipoprotein (a) concentration and cardiovascular disease in familial hypercholesterolemia. Clinical Biochemistry 66, 44–48 (2019) (cit. on p. 8).

83. Wang, L. et al. Functional variant in the SLC22A3-LPAL2-LPA gene cluster contributes to the severity of coronary artery disease. Arteriosclerosis, Thrombosis, and Vascular Biology 36, 1989–1996 (2016) (cit. on p. 8).

84. Yang, J., Lee, S. H., Goddard, M. E. & Visscher, P. M. GCTA: A Tool for Genome-wide Complex Trait Analysis. The American Journal of Human Genetics 88, 76–82. https://doi.org/10.1016/j.ajhg.2010.11.011 (Jan. 2011) (cit. on pp. 9, 17).

85. Weng, L.-C. et al. Heritability of atrial fibrillation. Circulation: Cardiovascular Genetics 10, e001838 (2017) (cit. on p. 13).

86. Möller, S. et al. The Heritability of Breast Cancer among Women in the Nordic Twin Study of CancerThe Heritability of Breast Cancer in NorTwinCan. Cancer Epidemiology, Biomarkers & Prevention 25, 145–150 (2016) (cit. on p. 13).

87. Drobni, Z. D. et al. Heritability of coronary artery disease: Insights from a classical twin study. Circulation: Cardiovascular Imaging 15, e013348 (2022) (cit. on p. 13).

88. Li, A.-l., Fang, X., Zhang, Y.-y., Peng, Q. & Yin, X.-h. Familial aggregation and heritability of hypertension in Han population in Shanghai China: a case-control study. Clinical hypertension 25, 1–7 (2019) (cit. on p. 13).

89. Ullemar, V. et al. Heritability and confirmation of genetic association studies for childhood asthma in twins. Allergy 71, 230–238 (2016) (cit. on p. 13).

90. Pociot, F. Type 1 diabetes genome-wide association studies: not to be lost in translation. Clinical & translational immunology 6, e162 (2017) (cit. on p. 13).

91. Willemsen, G. et al. The concordance and heritability of type 2 diabetes in 34,166 twin pairs from international twin registers: the discordant twin (DISCOTWIN) consortium. Twin Research and Human Genetics 18, 762–771 (2015) (cit. on p. 13).

92. Pedregosa, F. et al. Scikit-learn: Machine Learning in Python. Journal of Machine Learning Research 12, 2825–2830 (2011) (cit. on p. 15).

93. PRScs GitHub repository https://github.com/getian107/PRScs. Accessed: 2022-Feb-22 (cit. on p. 15).

94. Lello, L., Hsu, M., Widen, E. & Raben, T. G. Sibling variation in polygenic traits and DNA recombination mapping with UK Biobank and IVF family data. Scientific Reports 13, 376. https://doi.org/10.1038/s41598-023-27561-z (2023) (cit. on p. 16).

95. Bulik-Sullivan, B. K. et al. LD Score regression distinguishes confounding from polygenicity in genome-wide association studies. Nature genetics 47, 291–295 (2015) (cit. on p. 17).

96. *Heritability of >4,000 traits & disorders in UK Biobank* https://nealelab.github.io/UKBB_ldsc/index.html. Accessed: 2023-03-01 (cit. on p. 17).

97. *GCTA a tool for Genome-wide Complex Trait Analysis* https://yanglab.westlake.edu.cn/software/gcta/#Overview. Accessed: 2023-03-01 (cit. on p. 17).

98. Bellot, P., de los Campos, G. & Pérez-Enciso, M. Can deep learning improve genomic prediction of complex human traits? Genetics 210, 809–819 (2018) (cit. on p. 18).

99. Azodi, C. B. et al. Benchmarking Parametric and Machine Learning Models for Genomic Prediction of Complex Traits. G3: Genes, Genomes, Genetics 9, 3691–3702 (2019) (cit. on p. 18).

100. Abraham, G., Kowalczyk, A., Zobel, J. & Inouye, M. Performance and Robustness of Penalized and Unpenalized Methods for Genetic Prediction of Complex Human Disease. Genetic Epidemiology 37, 184–195. eprint: https://onlinelibrary.wiley.com/doi/pdf/10.1002/gepi.21698. https://onlinelibrary.wiley.com/doi/abs/10.1002/gepi.21698 (2013) (cit. on p. 18).

101. Alexander, D. H., Novembre, J. & Lange, K. Fast model-based estimation of ancestry in unrelated individuals. Genome research 19, 1655–1664 (2009) (cit. on p. 27).

102. Bycroft, C., Freeman, C. & Petkova, D. The UK Biobank resource with deep phenotyping and genomic data. Nature 562, 203–209 (cit. on p. 27).

103. *Current Asthma Demographics* https://www.lung.org/research/trends-in-lung-disease/asthma-trends-brief/current-demographics. Accessed: 2022-08-23 (cit. on p. 28).

104. Asthma Prevalence, Health Care Use and Mortality: United States, 2003-05 https://www.cdc.gov/nchs/data/hestat/asthma03-05/asthma03-05.htm. Accessed: 2022-08-23 (cit. on p. 28).

105. Huang, J.-L. Asthma severity and genetics in Taiwan. Journal of Microbiology, Immunology, and Infection= Wei Mian yu gan ran za zhi 38, 158–163 (2005) (cit. on p. 28).

106. Jan, I, Chou, W.-H., Wang, J.-D., Kuo, S.-H., et al. Prevalence of and major risk factors for adult bronchial asthma in Taipei City. Journal of the Formosan Medical Association 103, 259–263 (2004) (cit. on p. 28).

107. Rosser, F. J., Forno, E., Cooper, P. J. & Celedón, J. C. Asthma in Hispanics. An 8-year update. American journal of respiratory and critical care medicine 189, 1316–1327 (2014) (cit. on p. 28).

108. Alonso, A. et al. Incidence of atrial fibrillation in whites and African-Americans: the Atherosclerosis Risk in Communities (ARIC) study. American heart journal 158, 111–117 (2009) (cit. on p. 28).

109. Chiang, C.-E. et al. 2016 Guidelines of the Taiwan Heart Rhythm Society and the Taiwan Society of Cardiology for the management of atrial fibrillation. Journal of the Formosan Medical Association 115, 893–952 (2016) (cit. on p. 28).

110. Linares, J. D. et al. Prevalence of atrial fibrillation and association with clinical, sociocultural, and ancestral correlates among Hispanic/Latinos: The Hispanic Community Health Study/Study of Latinos. Heart rhythm 16, 686–693 (2019) (cit. on p. 28).

111. Hunt, B. R. Breast cancer prevalence and mortality among Hispanic subgroups in the United States, 2009– 2013. Journal of Cancer Epidemiology 2016 (2016) (cit. on p. 28).

112. American Cancer Society. Cancer Facts & Figures for African Americans 2019-2021. Atlanta: American Cancer Society, 2019. https://www.cancer.org/content/dam/cancer-org/research/cancer-facts-and-statistics/cancer-facts-and-figures-for-african-americans/cancer-facts-and-figures-for-african-americans-2019-2021.pdf. Accessed: 2022-08-23 (cit. on p. 28).

113. Liu, F.-C. et al. Epidemiology and survival outcome of breast cancer in a nationwide study. Oncotarget 8, 16939 (2017) (cit. on p. 28).

114. Lee, Y.-T. et al. Chin-Shan Community Cardiovascular Cohort in Taiwan–baseline data and five-year followup morbidity and mortality. Journal of clinical epidemiology 53, 838–846 (2000) (cit. on p. 28).

115. CDC 2021. Summary Health Statistics: National Health Interview Survey: 2018. Table A-1a. https://www.cdc.gov/nchs/nhis/shs/tables.htm and https://ftp.cdc.gov/pub/Health_Statistics/NCHS/NHIS/SHS/2018_SHS_Table_A-1.pdf. Accessed: 2022-08-23 (cit. on p. 28).

116. Lackland, D. T. Racial differences in hypertension: implications for high blood pressure management. The American journal of the medical sciences 348, 135–138 (2014) (cit. on p. 28).

117. Pan, H.-Y., Lin, H.-J., Chen, W.-J. & Wang, T.-D. Prevalence, treatment, control and monitoring of hypertension: a nationwide community-based survey in Taiwan, 2017. Acta Cardiologica Sinica 36, 375 (2020) (cit. on p. 29).

118. Lora, C. M. et al. Prevalence, awareness, and treatment of hypertension in hispanics/latinos with CKD in the Hispanic Community Health Study/Study of Latinos. Kidney medicine 2, 332–340 (2020) (cit. on p. 29).

119. Borchers, A. T., Uibo, R. & Gershwin, M. E. The geoepidemiology of type 1 diabetes. Autoimmunity reviews 9, A355–A365 (2010) (cit. on p. 29).

120. Kinney, G. L. et al. The prevalence of type 1 diabetes in Hispanic/Latino populations in the United States: findings from the Hispanic Community Health Study/Study of Latinos. Epidemiology 31, e7–e8 (2020) (cit. on p. 29).

121. Jiang, Y.-D., Chang, C.-H., Tai, T.-Y., Chen, J.-F. & Chuang, L.-M. Incidence and prevalence rates of diabetes mellitus in Taiwan: analysis of the 2000–2009 Nationwide Health Insurance database. Journal of the Formosan Medical Association 111, 599–604 (2012) (cit. on p. 29).

122. Lu, C.-L., Shen, H.-N., Chen, H.-F. & Li, C.-Y. Epidemiology of childhood Type 1 diabetes in Taiwan, 2003 to 2008. Diabetic medicine 31, 666–673 (2014) (cit. on p. 29).

123. Gheith, O., Farouk, N., Nampoory, N., Halim, M. A. & Al-Otaibi, T. Diabetic kidney disease: world wide difference of prevalence and risk factors. Journal of nephropharmacology 5, 49 (2016) (cit. on p. 29).

124. Lin, C.-C. et al. Time trend analysis of the prevalence and incidence of diagnosed type 2 diabetes among adults in Taiwan from 2000 to 2007: a population-based study. BMC public health 13, 1–10 (2013) (cit. on p. 29).

